# ENTAgents: AI Agents for Complex Knowledge Otolaryngology

**DOI:** 10.1101/2025.01.01.25319863

**Authors:** Tsz Kin Chan, Ngoc-Duy Dinh

## Abstract

Various healthcare applications based on large language models (LLMs) have emerged as LLMs show improved efficiency and error reduction. Recently, retrieval augmented generation (RAG) has been adopted frequently for LLM applications to solve the problem of hallucinations. Despite the success of RAG, it has its drawbacks, including incomplete semantic meanings, and large-scale dataset requirements. AI Agents have shown great potential in medicine and healthcare applications by leveraging their rich background knowledge and reasoning capabilities. In this paper, we introduce ENTAgents, a framework that utilizes both the RAG and multi-agent systems. To achieve better decision-making and enhanced explainability, we use the verbal reinforcement learning framework, Reflexion, as a reference for the task planning of ENTAgents. With the verbal feedback received from the last agent node, the agentic system can decide the best agent to utilize to improve the response of ENTAgents. We tested ENTAgents with various types of questions, including short questions, essay questions, and multiple-choice questions. ENTAgents improve accuracy by over 11.3% in handling short questions, with 2.78 folds in the length of the text and explaining clearly. ENTAgents shows higher accuracy and is more comprehensive in its answer compared to other LLM models. ENTAgents also demonstrates its ability to refine its answer with additional information to make it more thorough and educational. It also presents the capability to change its response to the correct answer in multiple-choice questions according to its self-reflection. Overall, ENTAgents is easy to use, can self-correct, and can provide detailed information in complex knowledge to users. We look forward to integrating ENTAgents into various healthcare scenarios, such as medical education and clinical decision support, particularly otolaryngology.

## 1. Introduction

Large language models (LLMs) have been in the spotlight in different industries since the release of ChatGPT, with the healthcare sector being one of the potential industries that can unlock the power of LLMs and new opportunities in medical applications. Numerous utilizations of LLMs have been proposed in the healthcare system, including health record management [1–2], medical image analysis [3–4], and medical research [5–6]. These applications have made significant impacts on the healthcare systems by minimizing human errors, enhancing efficiency, and providing medical research support [6–7]. Nevertheless, there are still concerns in LLM-based medical applications. Since LLMs often generate hallucinations, which is a response that is either factually incorrect or nonsensical. The hallucination rate of top LLM models is from 1.3% to 4.3% [8]. Although these values are not significant, concerns could be raised when an inaccurate response is generated by the LLM application, especially when it comes to clinical decision support, as reliability and trustfulness of a system are the most important principles in the healthcare sector.

To eliminate hallucinations in LLM, several techniques have been proposed in which the retrieval augmented generation (RAG) is one of the easiest and most cost-effective methods to deal with hallucinations. RAG is a technique that provides related accurate data from the external knowledge base to the LLMs. With the advancements of RAG techniques, RAG has shown promising results in alleviating hallucinations and a wide range of applications in healthcare [9–14]. Long et al introduced ChatENT, which is an AI framework specialized in otolaryngology, with the use of ChatGPT 4.0 and Retrieval-Augmented Language Modeling (RALM) [14], demonstrated a good capability of answering otolaryngology questions. Despite the success of the RAG systems, information loss, the need of large-scale dataset and time-consuming are the issues when building a RAG system. Often RAG fails to deliver the complete significant information to the LLM as the context is separated by the chunk size but not the contextual meaning, and a large volume of manipulated dataset is required to cover all the knowledge in one specialty [14,15]. It has evolved very fast in the medical field, it is not feasible to update the knowledge database frequently for RAG, especially for comprehensive healthcare applications. Therefore, there is a need to produce an LLM system that can be reliable and handle different tasks in the healthcare industry. Agentic systems have emerged as a popular approach in different applications due to their scalability and robustness, and these advantages are crucial to address the dynamic healthcare environments. Several agentic systems have been proposed to the healthcare industry, offering medical knowledge, diagnosis, clinical record management and more [16–20]. Nevertheless, the existing agentic systems use information from their database or repository to handle clinical diagnosis and treatment. They lack the ability to generate insights into specialized medications and unseen diseases [17,21–23]. In addition, the current agentic system is not capable of examining their answer or understanding the question, including irrelevant contexts as a result [24].

Here, we introduce an innovative LLM agentic framework, ENTAgents, which is a system addressing the medical specialty in otolaryngology and utilizing RAG and multiple agents to optimize its response. We take Reflexion, the verbal reinforcement learning framework, as a reference for the task planning of our agents [25]. ENTAgents demonstrate the ability to self-correction their answer based on the question, with the aid of information found by the agents. We show that ENTAgents can utilize different agents successfully and perform excellently in different types of questions. It is a cost-efficient, ease-of-use, time-saving approach to handling medical specialty questions without any need for fine-tuning and has great potential for medical education and clinical support for all medical specialties.

## 2. Methods

### 2.1 Overview

Fig. 1 shows the pipeline of our proposed ENTAgents framework. The first node, “Drafter”, generates an initial answer based on the question by LLM. After that, the LLM model examines its answer and generates critiques in our designated format for reflection, followed by the search queries according to the reflection critiques. Subsequently, RAG is performed to extract relevant information from our knowledge base to search queries in the second agent node. For the third node, we call a “Revisor,” which revises the drafted answer based on the reflection critiques and extracted information. Then, the LLM will provide new reflection critiques and search queries if they decide the revised answer is not the best. The “Supervisor” node will examine the question and the revised answer to choose the best agent to acquire new information or finish the execution. The agent searches for information or data with their tool and summarizes it. They are then passed back to the Revisor, and hence, a loop is created until the Supervisor decides that the answer does not need to be adjusted. Llama 3.1 70B model from Meta is used for the Drafter and Revisor, and Llama-3-Groq-70b-Tool-Use from Groq is used for the Supervisor and the agents, built for tool use and function calling. ENTAgent is built on the LangGraph platform.

**Figure 1:**
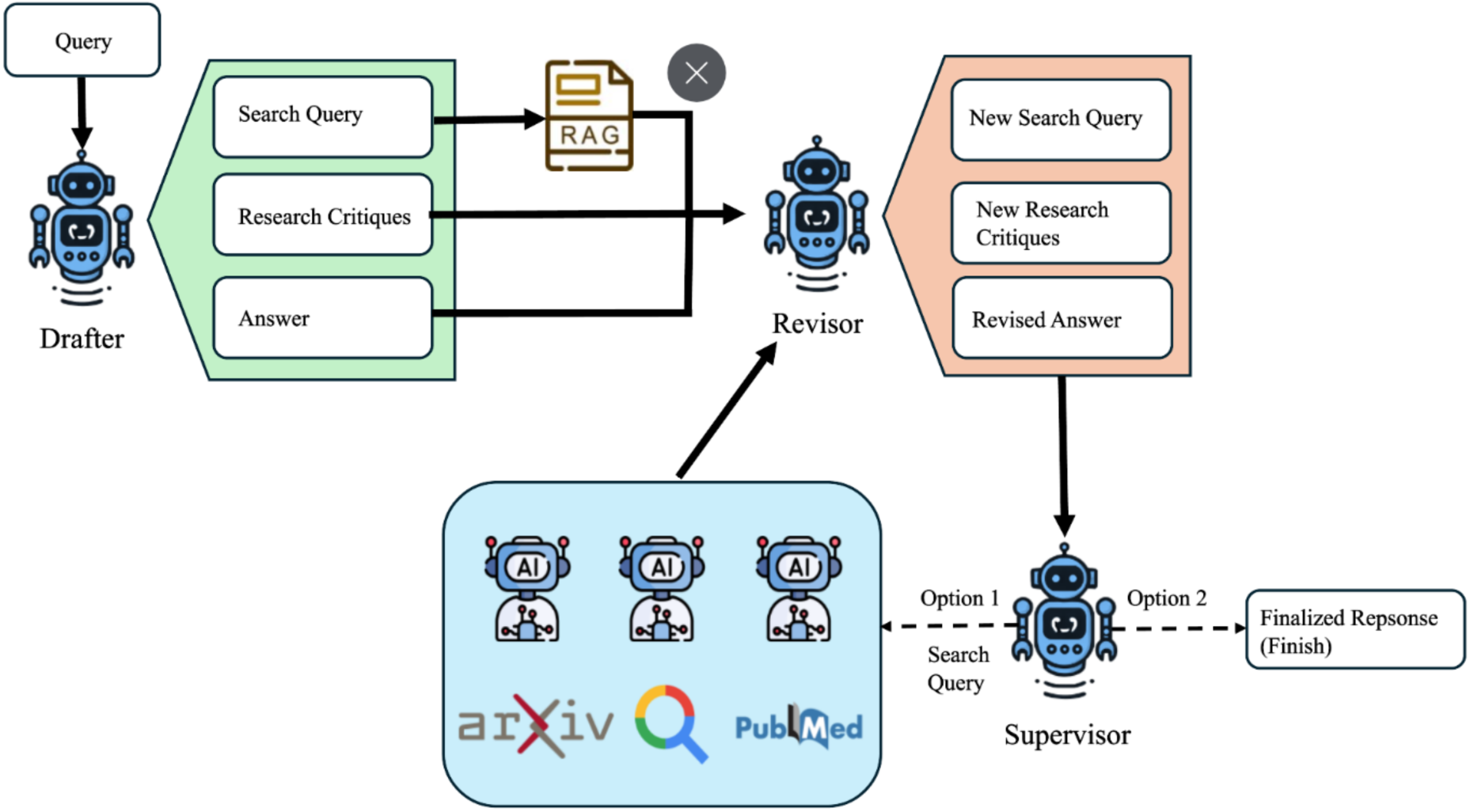
ENTAgents flow diagram from question input to answer generation.

### 2.2 RAG

To perform RAG, a knowledge base is first built with a variety of sources, including clinical guidelines, textbooks, and published articles, being chunked with the size of 1024 and chunk overlap of 200, including the whole knowledge base as a part of query extends the latency enormously and reaches the context window of Llama model easily. We used Voyage AI’s voyage-large-2-instruct to embed the chunked documents as vectors that represent their semantic meanings, and they were stored in a vector store initiated by the Facebook AI Similarity Search (FAISS) library. Having received the search queries, the embedding model also converts them into embedding vectors. For each of the queries, six chunks from the knowledge base are selected with the highest similarity between their embedding vectors and search queries’ embedding vectors.

### 2.3 Task Planning: Reflexion

We adopted the Reflexion framework proposed by the reinforcement learning proposed by Shinn et al [25] as the reinforcement learning of our ENTAgents. By leveraging Reflexion, AgenticENT can reflect on their answer and provide verbal feedback, which is stored in the episodic memory and added to the message passed to the next episode. The critiques of what is missing and what is superfluous in the answer were used in the self-reflection for short questions and essay questions, while critiques of inaccuracy and unnecessary were used for multiple choice questions. The Revisor reduces the redundant information in the last answer based on the critique of unnecessary and adds additional information provided by RAG or agents or changes the chosen answer according to the critique of missing and inaccuracy, respectively.

### 2.4 Multi-Agent System

We have three agents that use different tools to search for information from three tools: Google Search, ArXiv, and Pubmed. ArXiv tool searches for relevant articles from its database, which contains preprint and postprint scholarly articles, while the Pubmed tool finds published biomedical literature from the Pubmed database. Google Search tool uses the search engine to find the most relevant information from the website to the query. When the Supervisor decides the best agent to act on next, the search query is passed to that agent. The agent then executes the search query with its tool and concludes what it has found and how it can address the search query in a summary. Finally, the summary is sent to the revisor for revision again.

## 3. Results

The effectiveness and the capability of ENTAgents were evaluated using various types of questions, which are short answer questions, essay questions, and multiple-choice questions.

### 3.1 Short Answer Questions

19 short answer questions were used to assess the response of ENTAgents in otolaryngology, which includes 11 sample questions from the Royal College of Physicians and Surgeons of Canada (RCPSC). The accuracy of ENTAgents was evaluated with these 11 questions. ENTAgents based on Llama 3.1 scored 30.5 out of 37.5, a correct percentage of 81.3%, which is significantly higher than Llama 3.1 without Agentic, which is 70% (Figure 2). We can see the correct percentage of ENTAgents based on Llama 3.1 is a little higher than the paid model ChatGPT 4.0, which is 79.3%, but there are significant differences between ChatGPT 4.0 (ChatENT based on ChatGPT 4.0) and ENTAgents. The responses from ChatENT and ENTAgents were compared, as shown in Table 1. As ChatENT is not an open-source model, the responses from ChatENT were extracted from the supplementary information of the original paper. The average length of ENTAgents’s responses was approximately 258.3 and that of ChatENT’s responses was 92.5. The average latency of ENTAgents was 118.7 seconds. From Table 1, it can be observed that ENTAgents provided a more thorough answer than ChatENT did. The ChatENT failed to identify the number of sub-questions in this question, and it was only able to answer the second part of the question, while our ENTAgents provided complete answers for two parts of the question. For the first part of the question, both correct answers and explanations were provided by ENTAgents, making the answer understandable to the user. Both ENTAgents and ChatENT were able to generate three techniques required from the question and their functions were also provided. However, ENTAgents generated additional information related to the question and its answer. For instance, ENTAgents gave an example of intraoperative imaging and its functions during surgery so that the third technique could be explained clearly. Extra information related to preventing complications and the advantages of osteoplastic flap procedures was also provided.

**Figure 2:**
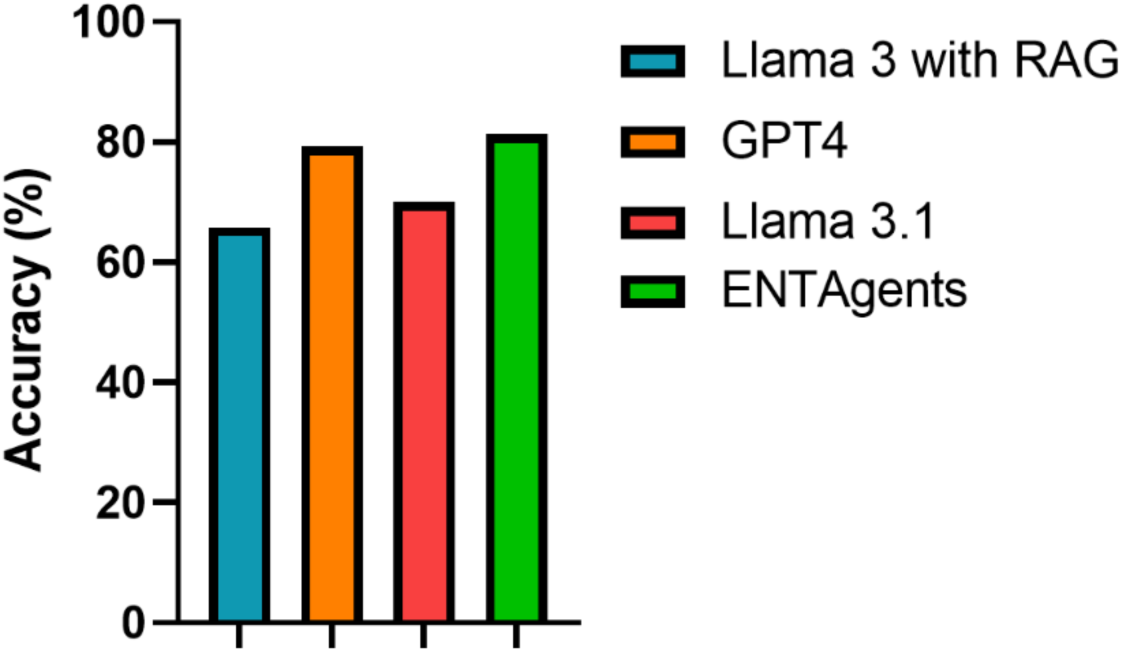
Accuracy rate of various LLM models for the short questions from the Royal College of Physicians and Surgeons of Canada (RCPSC).

**Table 1:** Comparison of the responses from ChatENT and ENTAgents to a short answer question. The italics paragraphs are the relevant additional information generated by the ENTAgents besides answering the question.

| Question | ChatENT | ENTAgents |
| --- | --- | --- |
| You are performing an osteoplastic flap procedure using a bi-coronal approach for drainage of mucocoeles.<br>a. In which soft tissue planes will you elevate the bicoronal flap?<br>b. Once the frontal bone is exposed, list THREE techniques to identify the margins of the frontal sinus prior to making the bony cuts for the osteoplastic flap. | Once the frontal bone is exposed, three techniques to identify the margins of the frontal sinus prior to making the bony cuts for the osteoplastic flap include:<br>- Caldwell view X-ray obtained from a distance of 6 feet: This radiographic view helps visualize the frontal sinus and its margins.<br>- Navigation system: Image- | To answer the question, I will break it down into two parts: a) the soft tissue planes for elevating the bicoronal flap, and b) techniques for identifying the margins of the frontal sinus prior to making the bony cuts for the osteoplastic flap.<br>a) The bicoronal flap is elevated in the subgaleal plane, taking care to avoid injury to the pericranium. |
|  | guided navigation equipment can be used to accurately identify the position of the frontal sinus and guide the surgeon during the procedure.<br>- Shining an endoscope transnasally: By inserting an endoscope through the nasal cavity, the surgeon can visualize the frontal sinus and its margins directly. | This plane is chosen to minimize bleeding and maximize exposure of the frontal bone.<br>b) Once the frontal bone is exposed, there are several techniques that can be used to identify the margins of the frontal sinus prior to making the bony cuts for the osteoplastic flap. These techniques include:<br>1) Using a 6-foot Caldwell radiograph to outline the frontal sinus on the bone prior to making the bony cuts. This technique allows for precise localization of the frontal sinus and helps to avoid injury to the surrounding bone.<br>2) Using a combination of anatomical landmarks and palpation to identify the margins of the frontal sinus. This technique relies on the surgeon's knowledge of the anatomy of the frontal bone and the location of the frontal sinus.<br>3) Using a navigation system to identify the margins of the frontal sinus in real-time. This technique uses intraoperative imaging to provide precise localization of the frontal sinus and helps to minimize the risk of complications. *Intraoperative imaging, such as MRI and CT* |
|  |  | scans, can be used to provide real-time feedback during the surgery, helping to identify potential complications and adjust the surgical plan accordingly.<br>Additionally, the osteoplastic flap procedure is effective in managing complications such as frontal outflow stenosis, mucocoele, infection, and paresthesia. Careful preoperative evaluation, precise surgical techniques, and postoperative care are recommended to manage these complications. Endoscopic sinus surgery and the osteoplastic flap procedure with frontal sinus obliteration are also effective methods for dealing with complications. Obtaining patient consent for possible complications and discussing management strategies preoperatively is also important. Furthermore, the osteoplastic flap can help improve outcomes and manage hydrocephalus. The osteoplastic flap procedure with frontal sinus obliteration is a viable option for managing frontal sinus pathology and preventing complications.<br>Overall, a comprehensive approach that includes careful preoperative evaluation, precise surgical techniques, and postoperative care is recommended to manage |
|  |  | *complications and improve outcomes in osteoplastic flap procedures.* |

### 3.2 Essay Questions

The ability to understand and generate subject matter content was also examined with 24 essay questions. The result shows that ENTAgents generated a significant number of relevant words compared to other LLM models (Figure 3). The ENTAgents generated answers with an average of 425.08 words, and the average latency ENTAgents required was 118.67 seconds. An average of 2.67 agents was invoked for ENTAgents to handle an essay question.

**Figure 3:**
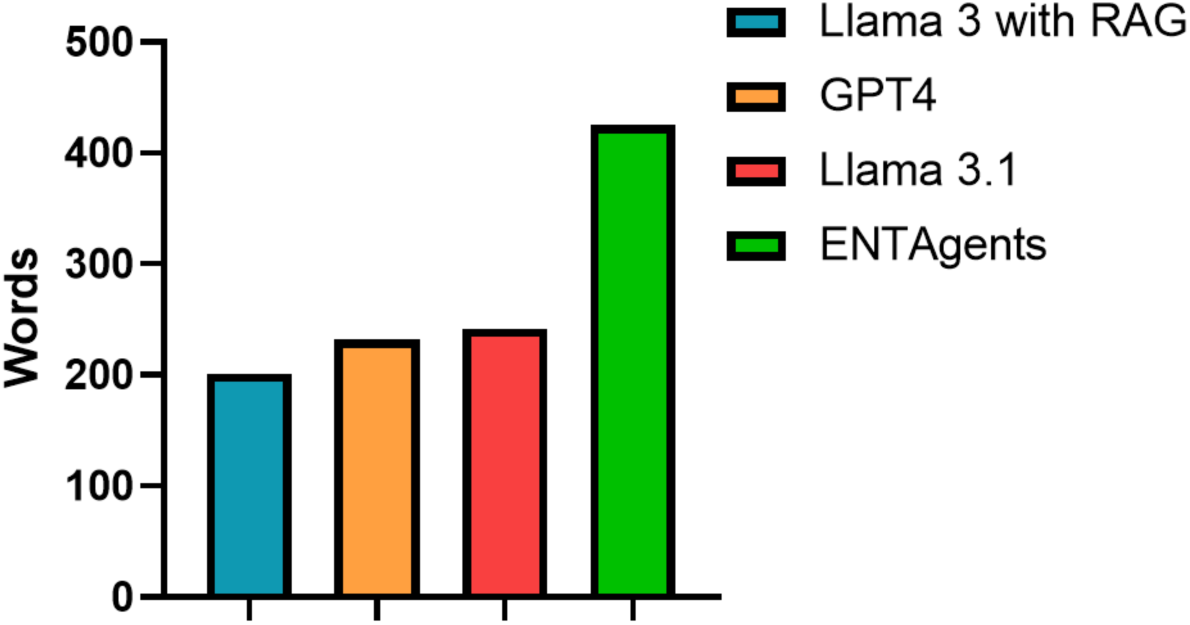
Length of responses from different LLMs to the essay questions.

The workflow of the ENTAgents is demonstrated in Table 2. In the initial response, ENTAgents reflected on its answer and stated that the information on aetiopathogenesis was insufficient and that the answer should be more concise and remove repetitive language. The search query was also generated based on the critiques of what is missing. After relevant information was found with the search query, the Revisor edited the previous answer based on the reflection critiques with the aid of the information. The Revisor reviewed its answer again and provided new critiques if it decided the answer required improvements. The supervisor received all the fields from the Revisor and decided that the Google agent should be used. Having revised the answer with the information provided by the agent, the Supervisor saw the answer being the best and finished the execution.

**Table 2:** Process of ENTAgents handling a question. The bold texts are the revised content in the latest answer compared to the previous answer.

|  |  |
| --- | --- |
| Example: Discuss the aetiopathogenesis, clinical features and management of tubotympanictype of chronic suppurative otitis media. |  |
| Step 1: LLM Drafter | Answer: Tubotympanic chronic suppurative otitis media (CSOM) is a type of infection that affects the middle ear. [...] This may include the treatment of allergies, the use of nasal decongestants, and the avoidance of upper respiratory tract infections.<br>Reflection:<br>1. Missing: One potential critique of this answer is that it lacks specific information on the microbiology of tubotympanic CSOM, including the most common pathogens involved and the antibiotic resistance patterns. Additionally, the answer could benefit from a more detailed discussion of the role of biofilms in the pathogenesis of CSOM.<br>2. Superfluous: The answer could be improved by removing some of the repetitive language and condensing the information into more concise paragraphs.<br>Search query: What are the most common pathogens involved in tubotympanic chronic suppurative otitis media and what are the current antibiotic resistance patterns? |
| Step 2: RAG | [...] These pathogens have been mentioned above and include *P. aeruginosa*, in a moist mastoid cavity, although patients may become sensitized to sulfamethoxazole or amphotericin. Systemic antibiotics should be used in refractory cases when specific pathogens are found on culture. Several quinolones such as ciprofloxacin, ofloxacin, and norfloxacin may be useful in these patients. Ideally, an ear with a tympanic membrane perforation should be free from infection for 3 months before tympanoplasty. [...] |
| Step 3: Revisor | Revised Answer: [...] *The most common pathogens involved in tubotympanic CSOM include Pseudomonas aeruginosa, followed by Proteus mirabilis and Staphylococcus aureus. [...] Furthermore, it is crucial to consider the role of biofilms in the pathogenesis of CSOM, as well as the potential for antibiotic resistance among the causative pathogens.* In conclusion, tubotympanic CSOM is a complex condition that requires a comprehensive management approach. By understanding the aetiopathogenesis, clinical features, and management options, healthcare providers can provide effective treatment and improve patient outcomes.<br>Reflection:<br>1. Missing: One potential critique of this revised answer is that it could |
|  | benefit from a more detailed discussion of the role of biofilms in the pathogenesis of CSOM, as well as the potential for antibiotic resistance among the causative pathogens. Additionally, the answer could be improved by including more information on the prevention and treatment of complications associated with tubotympanic CSOM.<br>2. Superfluous: The answer could be improved by removing some of the repetitive language and condensing the information into more concise paragraphs."<br>Search query: What are the current treatment options for biofilm-related infections in tubotympanic chronic suppurative otitis media? |
| Step 4: Supervisor | Next: Google |
| Step 5: Google Agent | [...] The management of CSOM is complex and can involve medical and/or surgical approaches. Medical management for a dry ear is essential before surgical treatment, as biofilms are most prevalent in CSOM. [...]<br>Globally, CSOM occurs in 4.76% of the population, with hearing impairment present in 30 children per 10,000. |
| Step 6: Revisor | Revised Answer: [...] *Globally, CSOM affects 4.76% of the population, with hearing impairment present in 30 children per 10,000. The tubotympanic type of CSOM is characterized by a chronic infection of the middle ear, with a perforated tympanic membrane and a discharge.* [...]<br>Reflection:<br>1. Missing: One potential critique of this revised answer is that it could benefit from a more detailed discussion of the role of biofilms in the pathogenesis of CSOM, as well as the potential for antibiotic resistance among the causative pathogens. Additionally, the answer could be improved by including more information on the prevention and treatment of complications associated with tubotympanic CSOM<br>2. Superfluous: The answer could be improved by removing some of the repetitive language and condensing the information into more concise paragraphs.<br>Search query: What are the current treatment options for biofilm-related infections in tubotympanic chronic suppurative otitis media? |
| Step 7: Supervisor | Next: FINISH |

An example of the answers to an essay question from Llama 3.1 and ENTAgents was compared in Table 3. The ENTAgents provided a more comprehensive answer to the question than Llama 3.1. In the pathogenesis part, the answers of ENTAgents included detailed and specific pathological processes and consequences of cholesteatoma and covered the impact on surrounding structures. In addressing the etiology of cholesteatoma, Llama 3.1 provided only a list of factors associated with the condition, whereas ENTAgents offered a detailed explanation. ENTAgents not only detailed how these factors contribute to cholesteatoma development but also discussed the different types of cholesteatoma and their specific causes. Additionally, while Llama 3.1 produced a simple list of clinical features, ENTAgents elaborated on the underlying causes of these features. It then presented a thorough response, addressing complications seen in both typical and advanced cases. In terms of cholesteatoma management, ENTAgents described not only the primary approaches and surgical techniques based on the size and location of the cholesteatoma but also mentioned the use of imaging tools to aid in management. Overall, ENTAgents provided a more comprehensive and structured response that covered all aspects of the query with detailed explanations.

**Table 3:** Comparison of the responses from ENTAgents and Llama 3.1 to an essay question.

| Questions | What is cholesteatoma? Describe its pathogenesis, etiology, clinical features and management. |  |
| --- | --- | --- |
| Models | Llama 3.1-70b | ENTAgents |
| What is cholesteatoma? | A cholesteatoma is a type of skin cyst located in the middle ear. It is a sac filled with skin cells and other debris, which can cause destruction of the surrounding bone and tissue. | Cholesteatoma is a type of skin cyst located in the middle ear. It is characterized by the abnormal growth of skin cells within the middle ear, which can lead to the destruction of surrounding bone and tissue. |
| Pathogenesis | The pathogenesis of cholesteatoma involves the accumulation of skin cells in the middle ear, which can occur due to a blockage in the Eustachian tube, a perforation in the eardrum, or other factors. | The pathogenesis of cholesteatoma involves the formation of an inflamed ectopic mass of keratinizing squamous epithelium in the middle ear and mastoid bone. This can lead to the erosion of adjacent bony structures and is often associated with chronic suppurative otitis media. |
| Etiology | The etiology of cholesteatoma can be congenital or acquired, with the latter being more common.<br>Acquired cholesteatomas are often associated with chronic otitis media, Eustachian tube dysfunction, and other conditions. | The etiology of cholesteatoma is multifactorial and can be attributed to various factors such as chronic otitis media, eustachian tube dysfunction, and congenital abnormalities. Chronic otitis media can lead to the formation of a cholesteatoma by causing a perforation in the eardrum, which allows skin cells to enter the middle ear. Eustachian tube dysfunction can also contribute to the development of cholesteatoma by causing a buildup of mucus and debris in the middle ear, which can become infected and lead to the growth of skin cells.<br>There are three main types of cholesteatoma: congenital, primary acquired, and secondary acquired. Congenital cholesteatoma is a rare condition that is present at birth and is thought to be caused by a genetic mutation. Primary acquired cholesteatoma is the most common type and is caused by a perforation in the eardrum or by the migration of skin cells into the middle ear through the eustachian tube. Secondary acquired cholesteatoma is caused by the implantation of skin cells into the middle ear through trauma or surgery. |
| Clinical Features | Clinical features of cholesteatoma include hearing loss, ear fullness, discharge, and vertigo. | The clinical features of cholesteatoma can vary depending on the size and location of the cyst. Common symptoms include hearing loss, ear fullness, and discharge. In advanced cases, cholesteatoma can cause more serious complications such as facial paralysis, vertigo, and meningitis. |
| Management | Management of cholesteatoma typically involves surgical removal of the cyst, as well as treatment of any underlying conditions. In some cases, antibiotics or other medications may be prescribed to manage symptoms and prevent complications. | The management of cholesteatoma primarily involves surgical intervention, aiming to eliminate the disease, control infection, and preserve or restore hearing. Long-term monitoring and follow-up are crucial to prevent recurrence.<br>Different surgical approaches are used depending on the size and location of the cholesteatoma, including atticotomy, atticoantrostomy, or limited mastoidectomy. In cases where the cholesteatoma is present in an only hearing ear, a more conservative approach is often recommended to minimize the risk of developing a postoperative sensorineural hearing loss.<br>Imaging studies, such as CT scans, can be useful in the diagnosis and management of cholesteatoma. These studies can help to identify the size and |
|  |  | location of the cyst, as well as any damage to surrounding tissue and bone. |

### 3.3 Multiple Choice Question (MCQ)

Finally, we challenged our ENTAgents with 992 MCQs from BoardVitals, which are US board review practice questions that follow the exam content outline for US otolaryngology qualifying examinations. The MCQ question banks consist of 9 sections, each with three levels of difficulty, which are hard, moderate, and easy, and questions from subspecialties including Allergy, Endocrine, Head and Neck, Laryngology, Otology, Pediatrics, Pharmacology, Plastic and Reconstructive and Sleep. The distributions of questions in different subspecialties in the question banks are presented in Table 4.

**Table 4:**
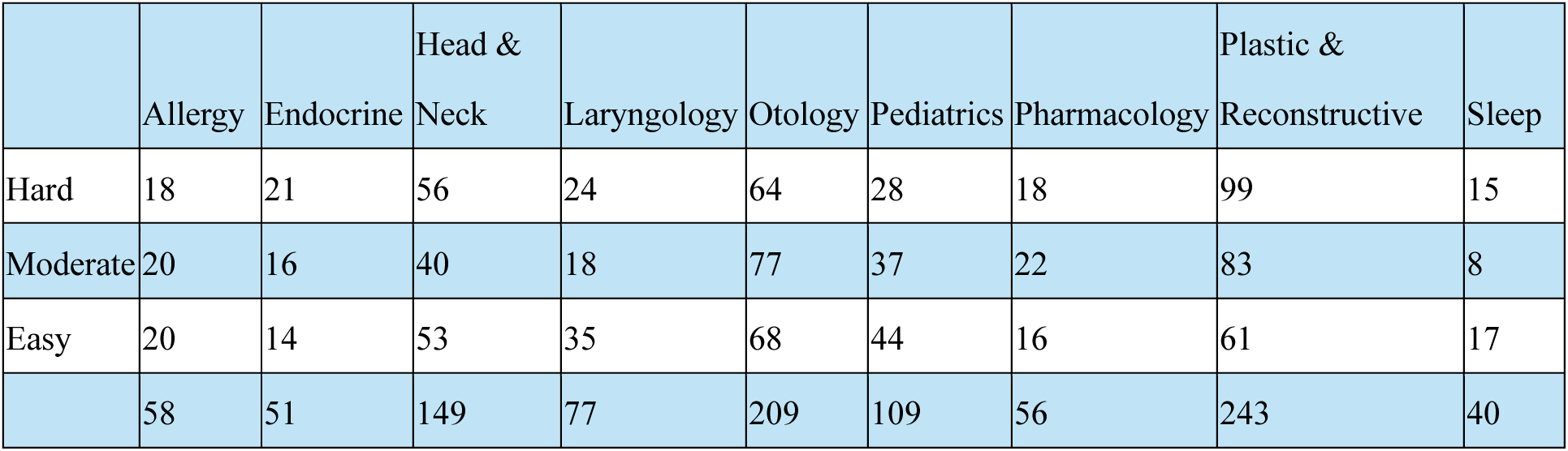
Distributions of the MCQs in 9 subspecialties of otolaryngology.

The performance of ENTAgents in answering questions from all specialties is summarized in Figure 4. The results were sorted by the score of ENTAgents from lowest to highest. ENTAgents obtained scores above 70% in all specialties, with the score in Plastic and Reconstructive being the lowest, 68.4%. ENTAgents’s highest score was in Allergy, which is 91.4%. The overall performance of ENTAgents was also calculated and compared to the average correct percentage of humans, obtained from the publicly available data from BoardVitals. ENTAgents had an overall performance of 75.5, 4.5% better than humans, scoring 71%.

**Figure 4:**
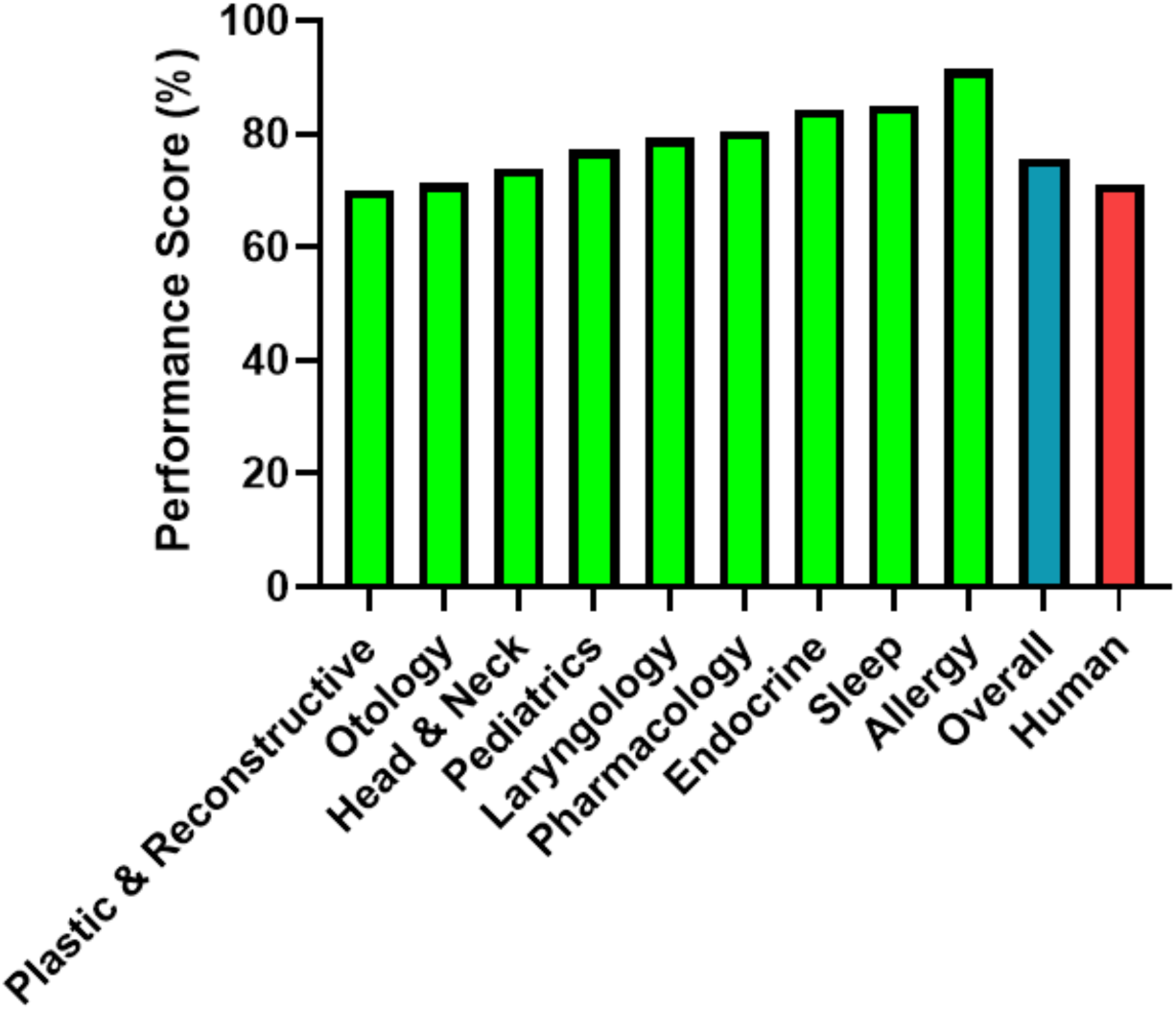
The scores of ENTAgent for the otolaryngology MCQ.

Table 5 shows an example of the response of ENTAgents to an MCQ. Without RAG and the agents, Llama 3.1 initially chose option B as its answer. Then, the answer was considered as not thorough by ENTAgents. With the retrieval from the knowledge base, it was discovered that the actual likelihood should be 45%. The revisor agent immediately changed its choice to C, which was the correct answer. ArXiv agent was invoked, and it summarized the scholar articles it searched for about ultrasound of the nodule to its response.

**Table 5:**
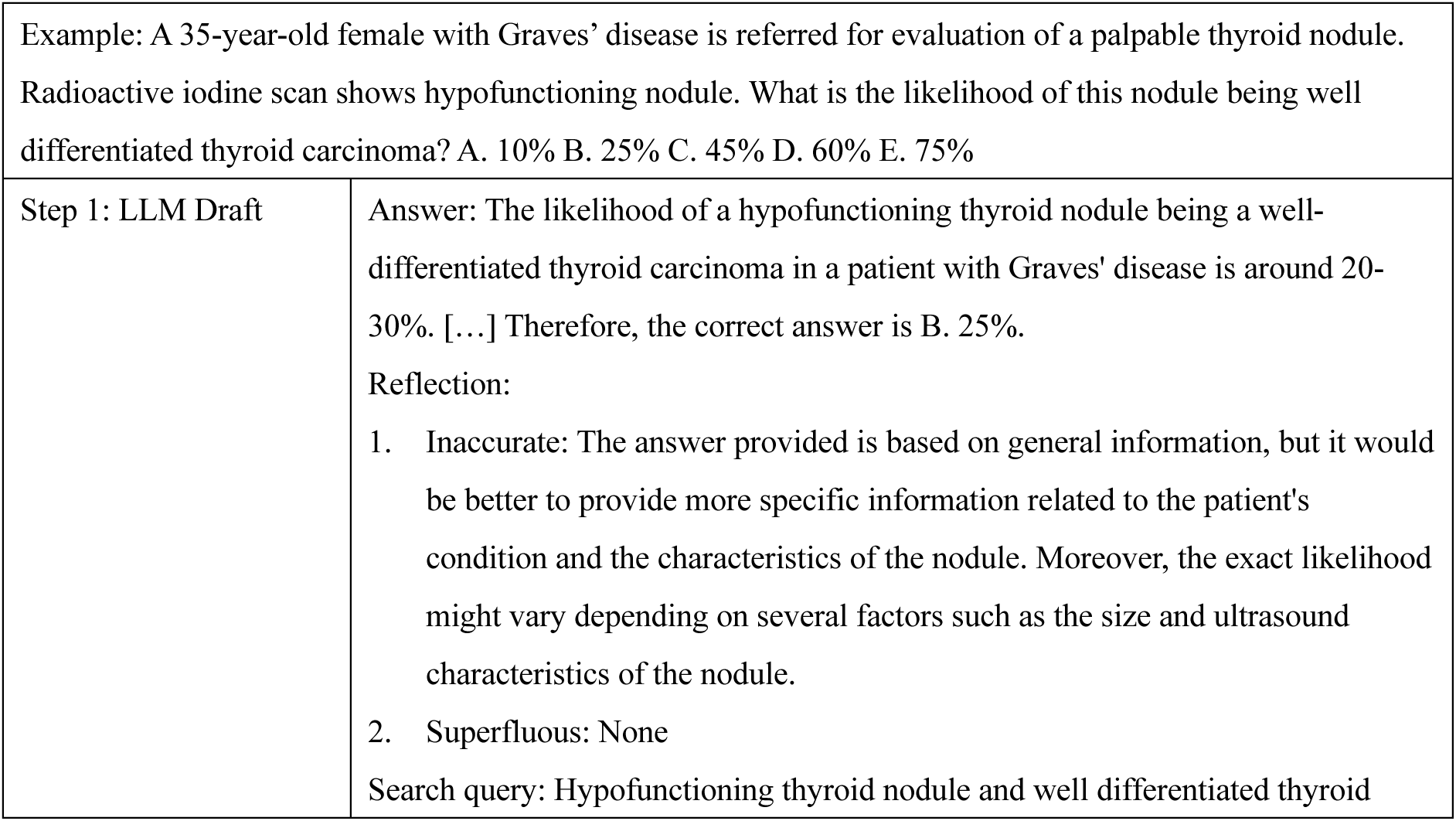
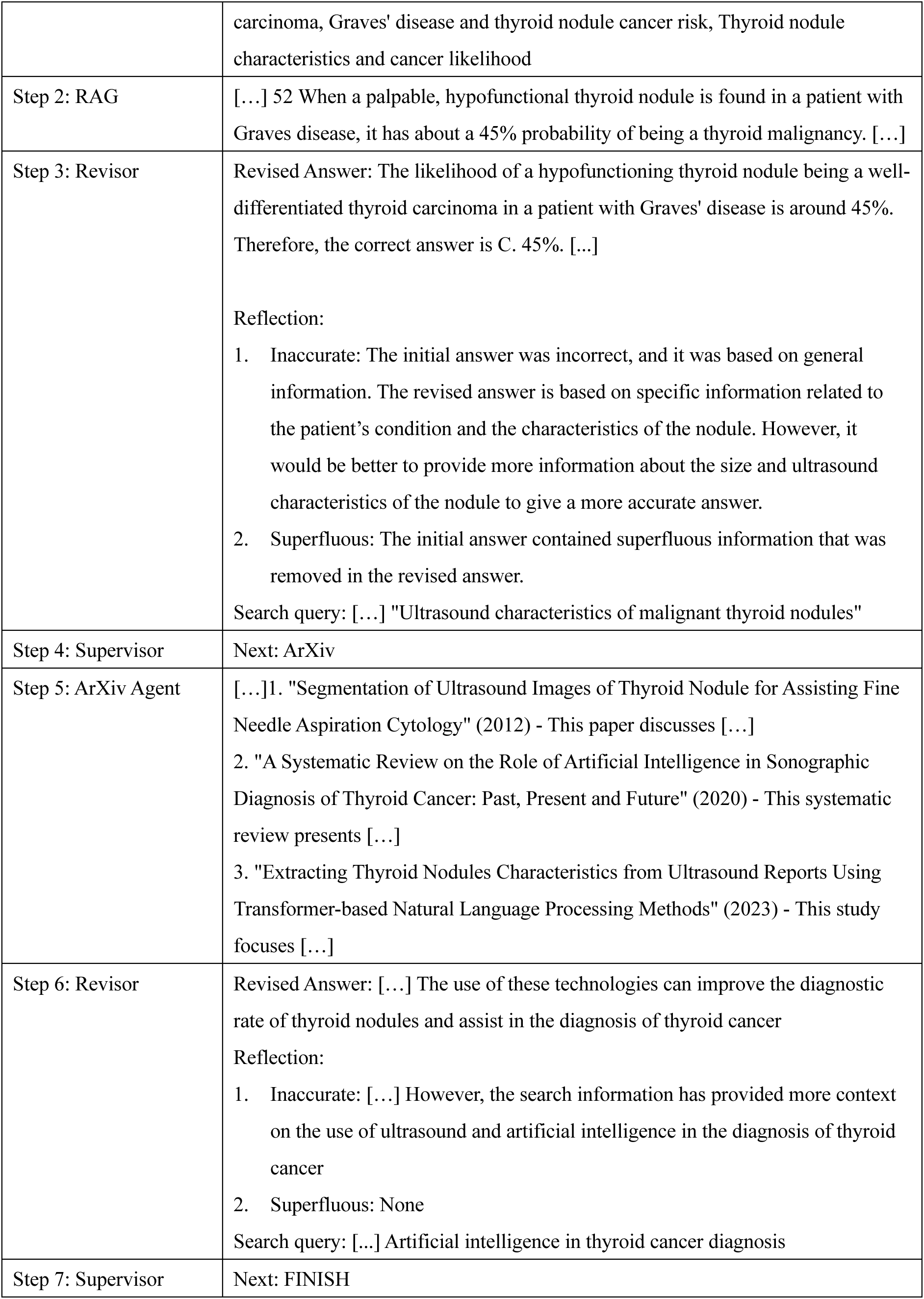
Example of the self-correction of ENTAgents to an MCQ.

## 4. Discussion

These results show that ENTAgents has the capability to provide comprehensive answers with the aid of reflection critiques. The reflection critiques enable ENTAgents to comment on its answers and suggest what can be improved. In answering short questions and essay questions, ENTAgents can provide the relevant additional information only and remove the abundant texts, making the answer more informative but also concise. ENTAgents provides more informative responses than other AI models, demonstrating enhanced accuracy. For the MCQ question, ENTAgents demonstrates the ability to change its option to the correct one, the Revisor examines these messages from RAG and agents to adjust its answer if needed. Utilizing agents and RAG can supplement the AI framework with up-to-date, highly specific, and detailed information, improving the accuracy and completeness of its responses. Since the RAG database contains knowledge of otolaryngology, ENTAgents is therefore able to draw from verified information and provides accurate answers for this field. The multi-agent system is critical for complex queries, which requires a diverse range of answers and the latest information. The agents can search websites and the literature database to find up-to-date developments related to the question. This approach can effectively address essay questions, allowing inquirers to gain insights into recent advancements and research developments in otolaryngology rather than relying solely on static, pre-existing information from RAG. Thus, our framework offers three distinct advantages over the other LLM agent applications. The first one is extensive search capabilities. For example, ENTAgents draws information from both the RAG knowledge base and external sources through the use of agents. Therefore, it does not require the continuous update of the pertinent sources from the field. This enhances the accuracy, completeness and timeliness of its responses, offering ENTAgents the ability to address a wide range of questions, from fundamental concepts to the latest advancements. The second is self-correction. This means that the reflection framework allows ENTAgents to evaluate their responses according to linguistic feedback. As the use of agents is iterative, the framework ensures that agents are leveraged only when flaws or inaccuracies are detected in the response. Furthermore, it removes superfluous information, ensuring that the final answer is both comprehensive and free of redundancies. Finally, ENTAgents is ease of use. Since the RAG knowledge base is built by external sources and ENTAgents’s ability to self-correct derived from its own reflective critiques, generated through prompting alone, fine-tuning is not required within our framework. The reflection critiques and search queries are provided along with the answers so that users can understand clearly the generation of the response of ENTAgents, enhancing its explainability. There are many opportunities for ENTAgents to be utilized across different fields or situations; adjustments to the knowledge base and the reflection prompts only allow ENTAgents to provide its response according to other criteria as needed. In addition, the model we used, Llama 3.1, is an open source LLM model, which offers full access and capability to develop advanced solutions, unlike other proprietary models, and hence we can modify ENTAgents easily for future applications.

Our work illustrates the potential applications of ENTAgents, including medical education, patient education, and clinical support. First, ENTAgents can be leveraged to provide medical education to students effectively. With the ability to retrieve solid knowledge from guidelines, books or academic papers in the knowledge base in a second, the learning process can be streamlined, and students can absorb concentrated information efficiently. ENTAgents can also be used in revision for medical students, given its ability to respond to various types of questions comprehensively. Healthcare professionals can also benefit from learning about the advancements in their field. The latest literature and academic symposiums can be found by the agents, and they will be summarized to the users. Professionals can then learn about cutting-edge technology and recent developments in the field. On top of that, the public can be educated by ENTAgents. The flexibility of ENTAgents to change its response makes it easy to explain difficult medical terms or knowledge to the layman, and it can be developed into the mobile application. The patients can thus easily access ENTAgents to have a better understanding of precautions and treatments of the diseases. Lastly, the accessibility of ENTAgents to various sources of scholarly articles provides insights to clinicians to handle rare cases and assist them in decision-making. Since the explainable concerns can be dampened with the reflection critiques provided, physicians can understand the workflow of ENTAgents and be assured to take the recommendations of ENTAgents.

Although our framework excels in generating comprehensive responses to users, the latency of ENTAgents needs to be improved. ENTAgents consumes a lot of time searching the information and revising the answers by the agents. Thus, it will not be a user-friendly application to wait such a long time for its response, further improvement in its speed is paramount. Moreover, the responses of ENTAgents deviated from the prescribed format several times. While the response structure is organized into distinct sections such as answer, reflective critique, and search query, the agent occasionally produces reflective content as an answer, resulting in wrong outputs and altering the workflow. Prompting techniques should be refined to ensure consistent adherence to the designated response format. Lastly, ENTAgents focuses on integrating verbal reinforcement learning within a multi-agent system to deliver comprehensive responses. Therefore, it lacks multimodal capabilities to store visual or audio information within its knowledge base and respond to queries beyond text-based inputs. Despite these limitations, the ENTAgents presents an advanced framework capable of generating accurate and thorough responses, demonstrating significant potential for diverse applications.

Our future research will first emphasize enhancing the multimodal capabilities of ENTAgents to enable the interpretation of visual data, particularly medical images sourced from both the RAG and the agents, and provide visual explanations to the users. As a result, ENTAgents will be better equipped to tackle complex medical cases across different modalities, offering clearer and more comprehensible responses for users. In addition, we aim to scale our knowledge database to encompass diverse entities, including diseases and conditions, imaging data, anatomical features, and more. This data will be structured well and interlinked to organize a healthcare knowledge graph for ENTAgents. With this approach, ENTAgents will deliver higher quality and more relevant answers effectively.

## Data Availability

All data produced in the present study are available upon reasonable request to the authors

## Acknowledgements

We gratefully acknowledge the funding provided by the Research Grant Council of Hong Kong, General Research Fund (Ref No. 14211223).

## Supplementary Material 1: Used documents for RAG knowledge base

**Table 1:**
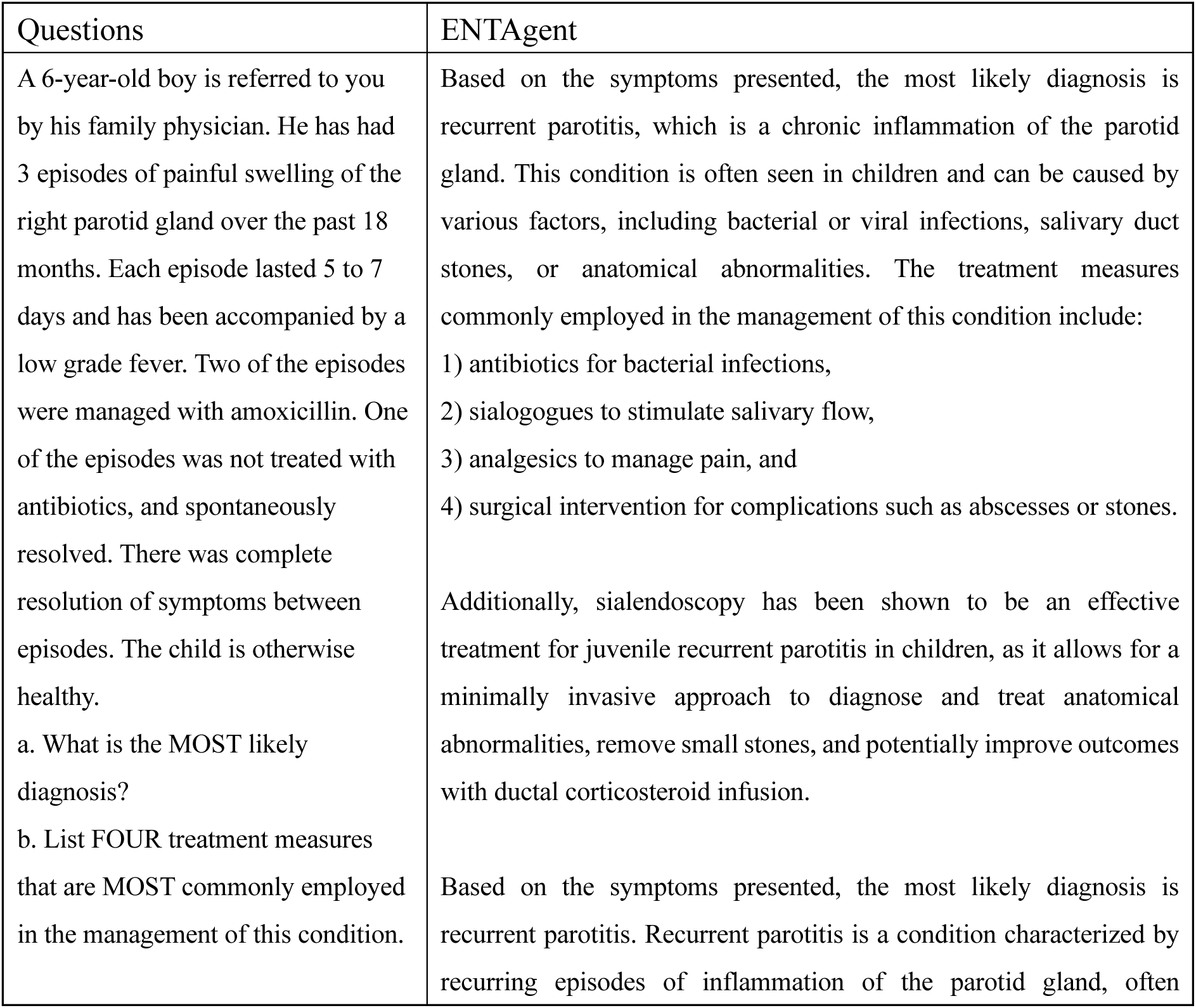
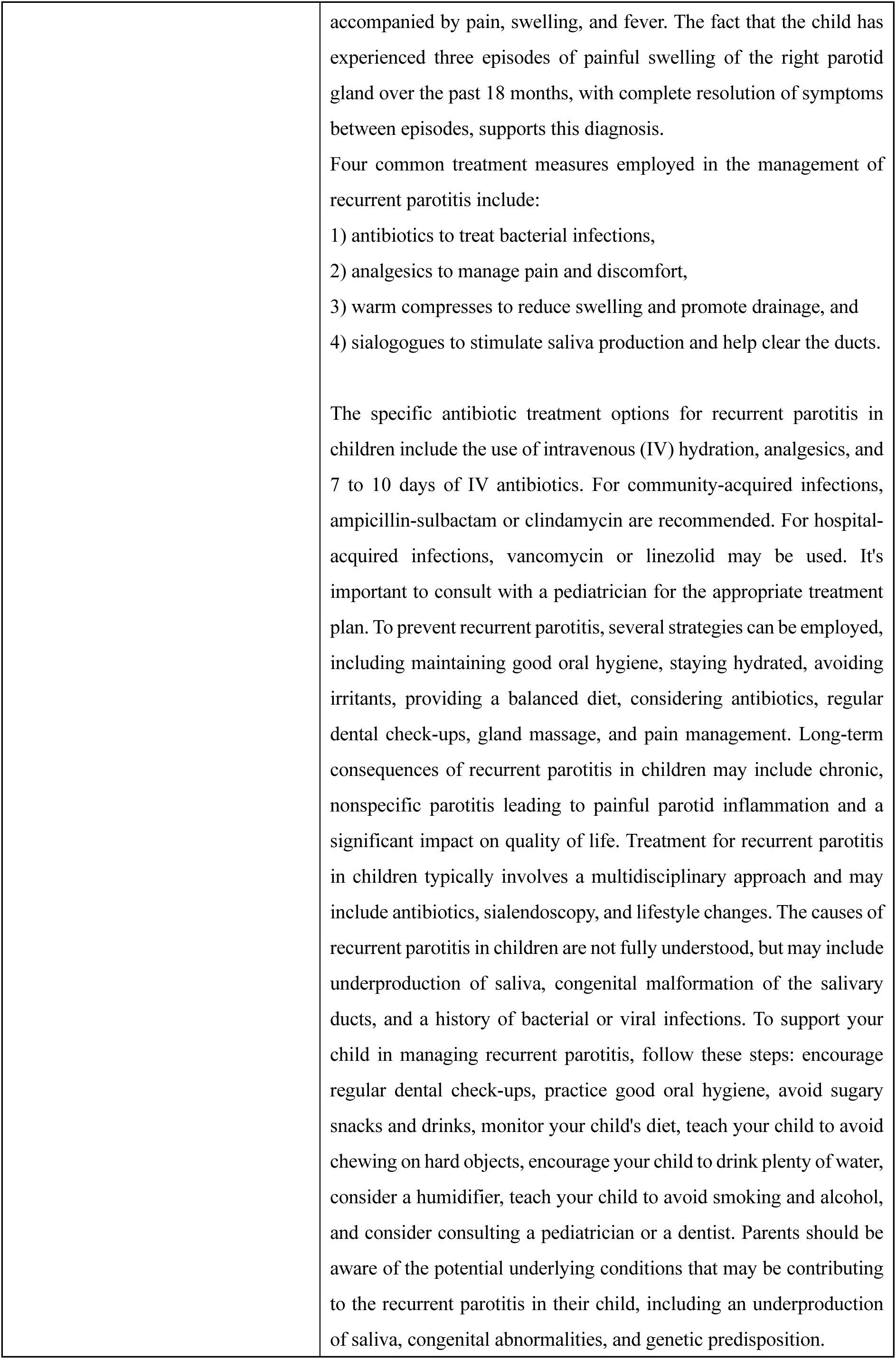
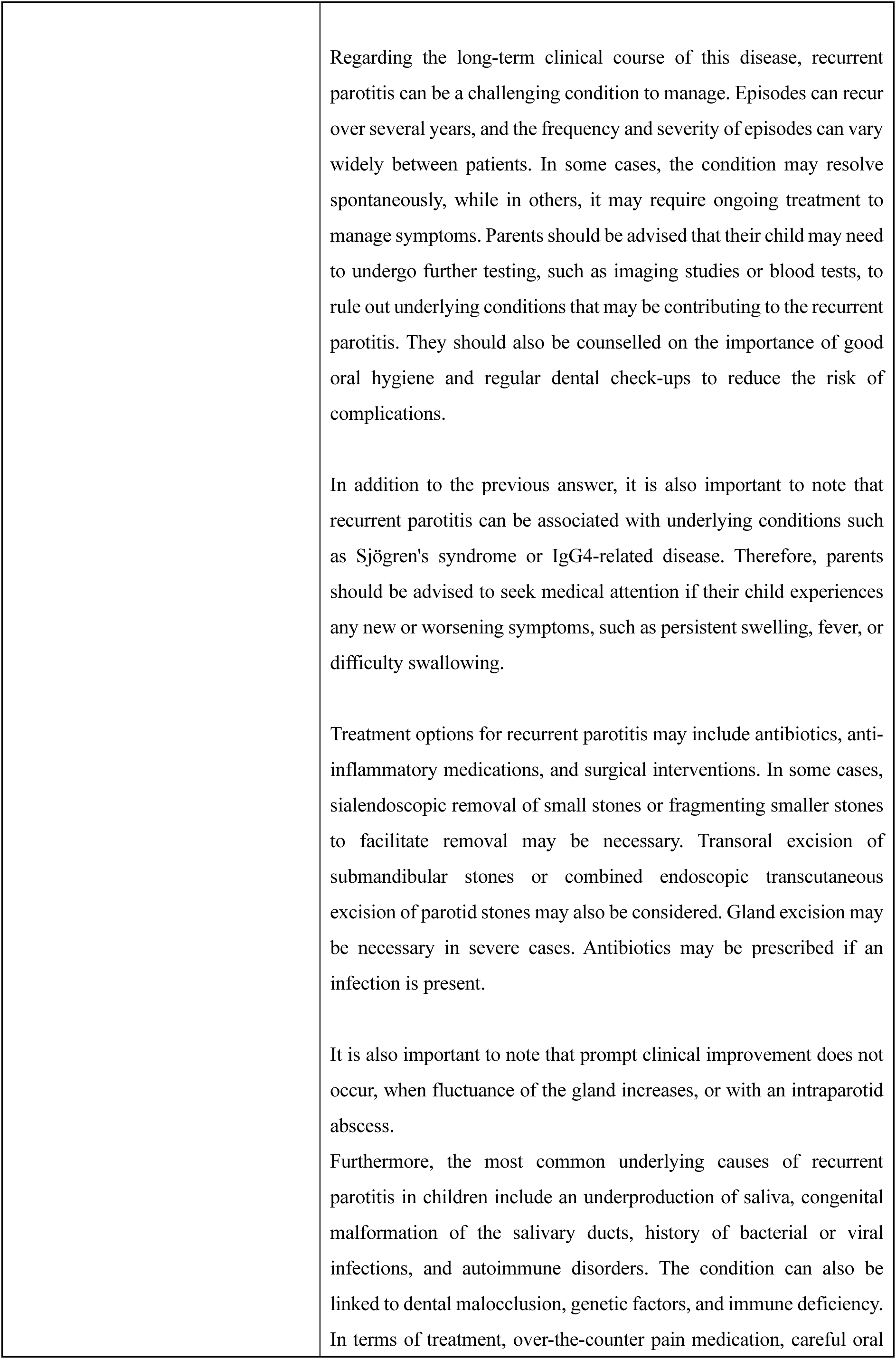
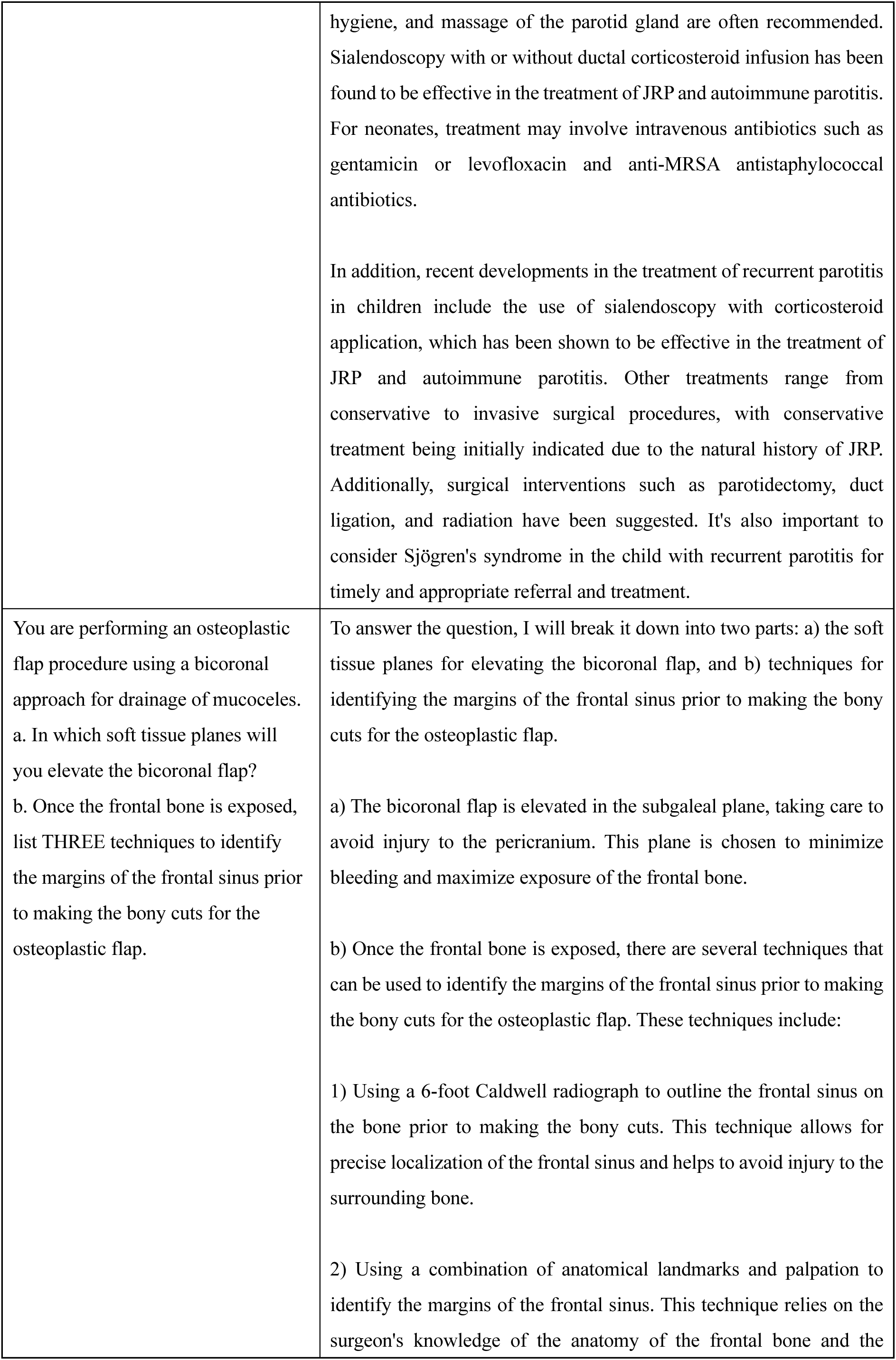
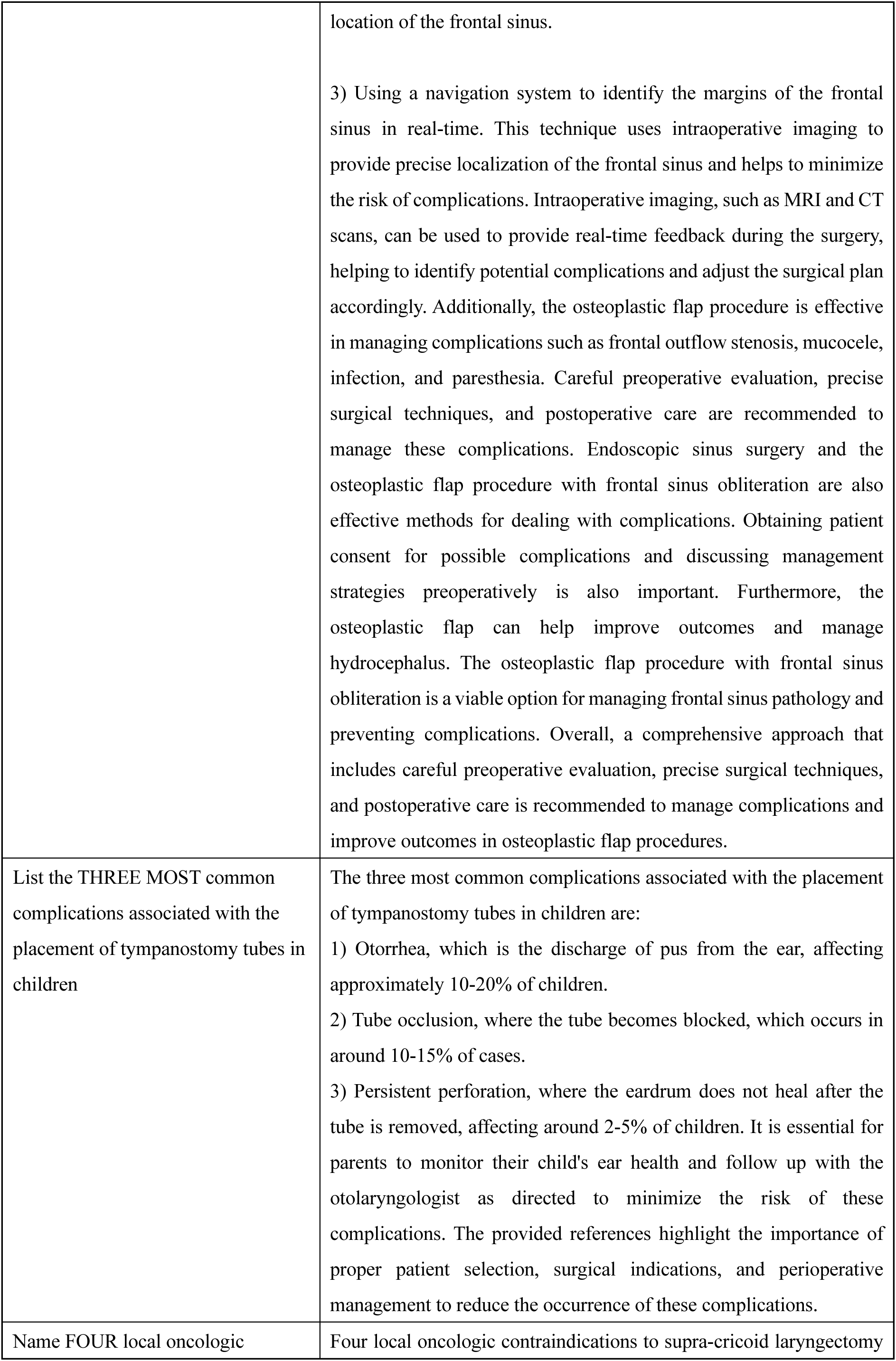
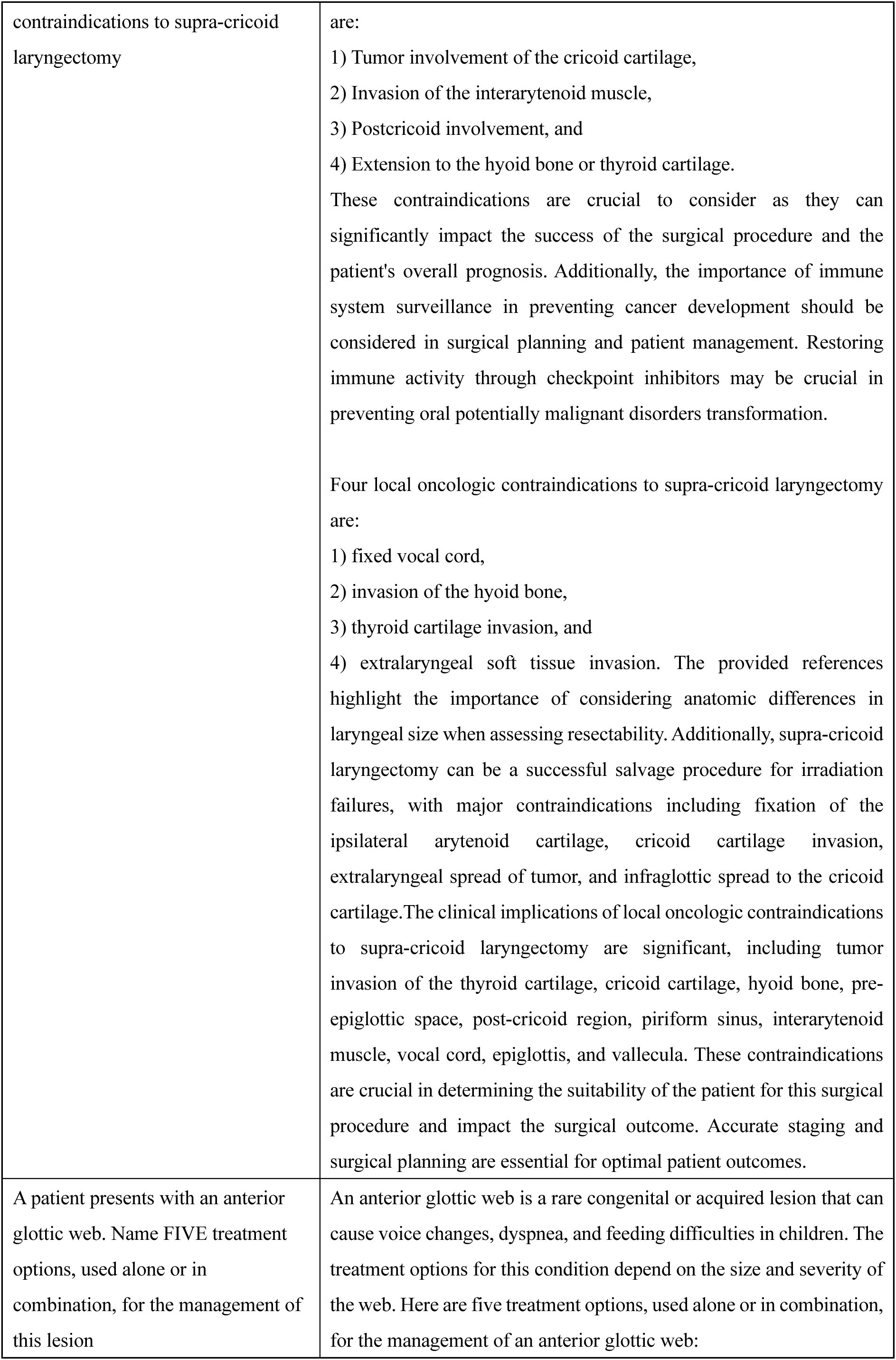
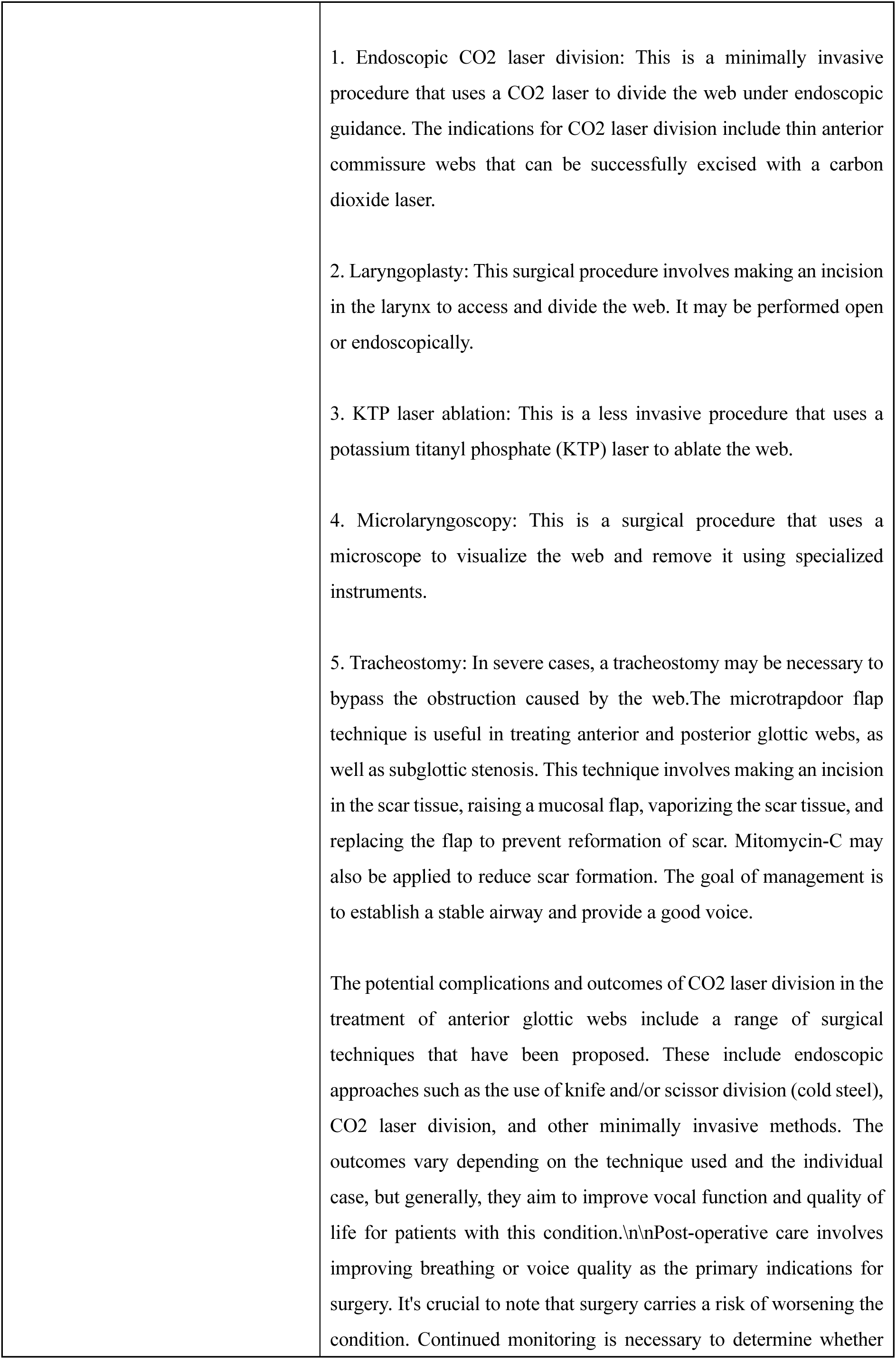
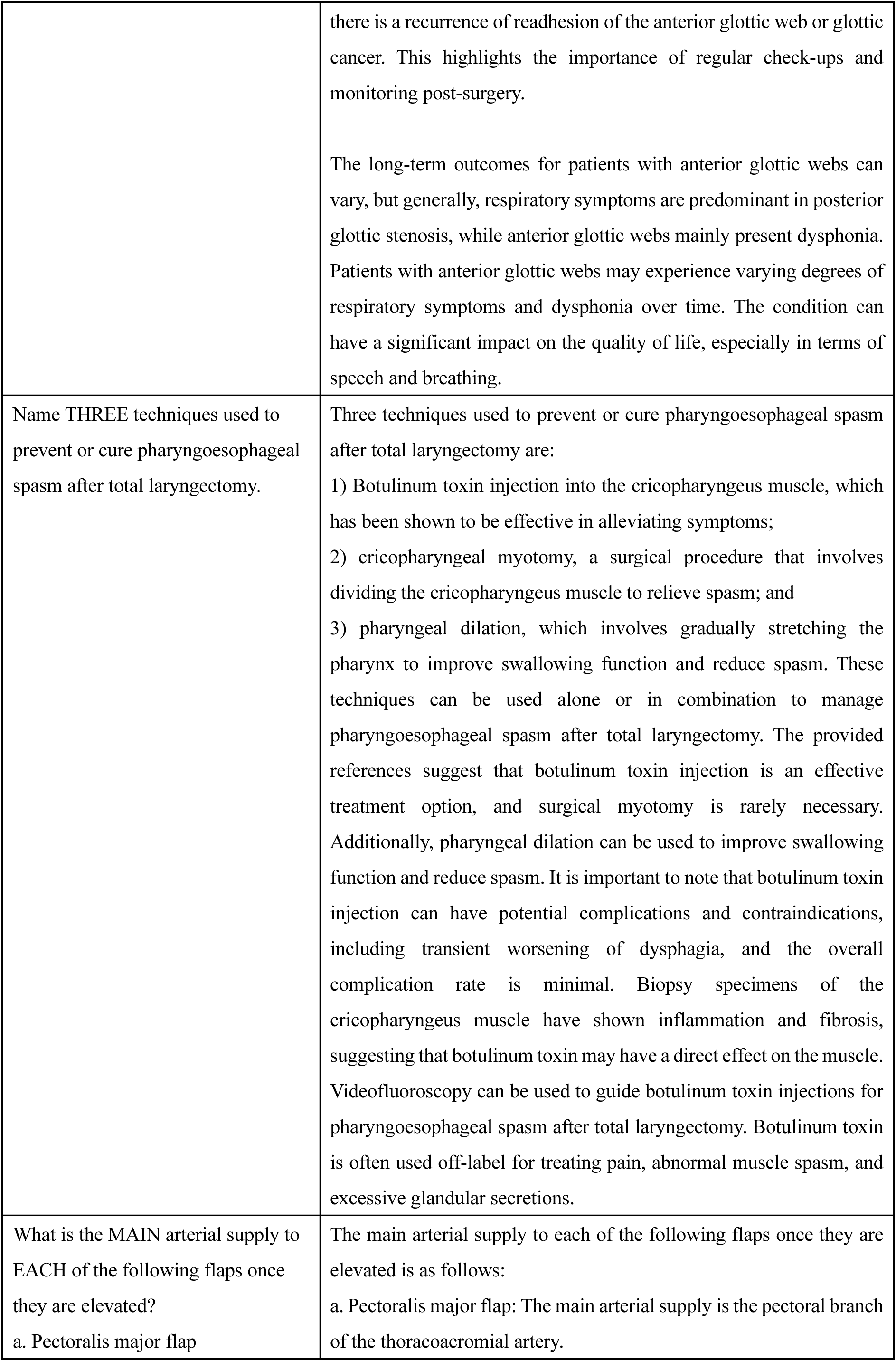
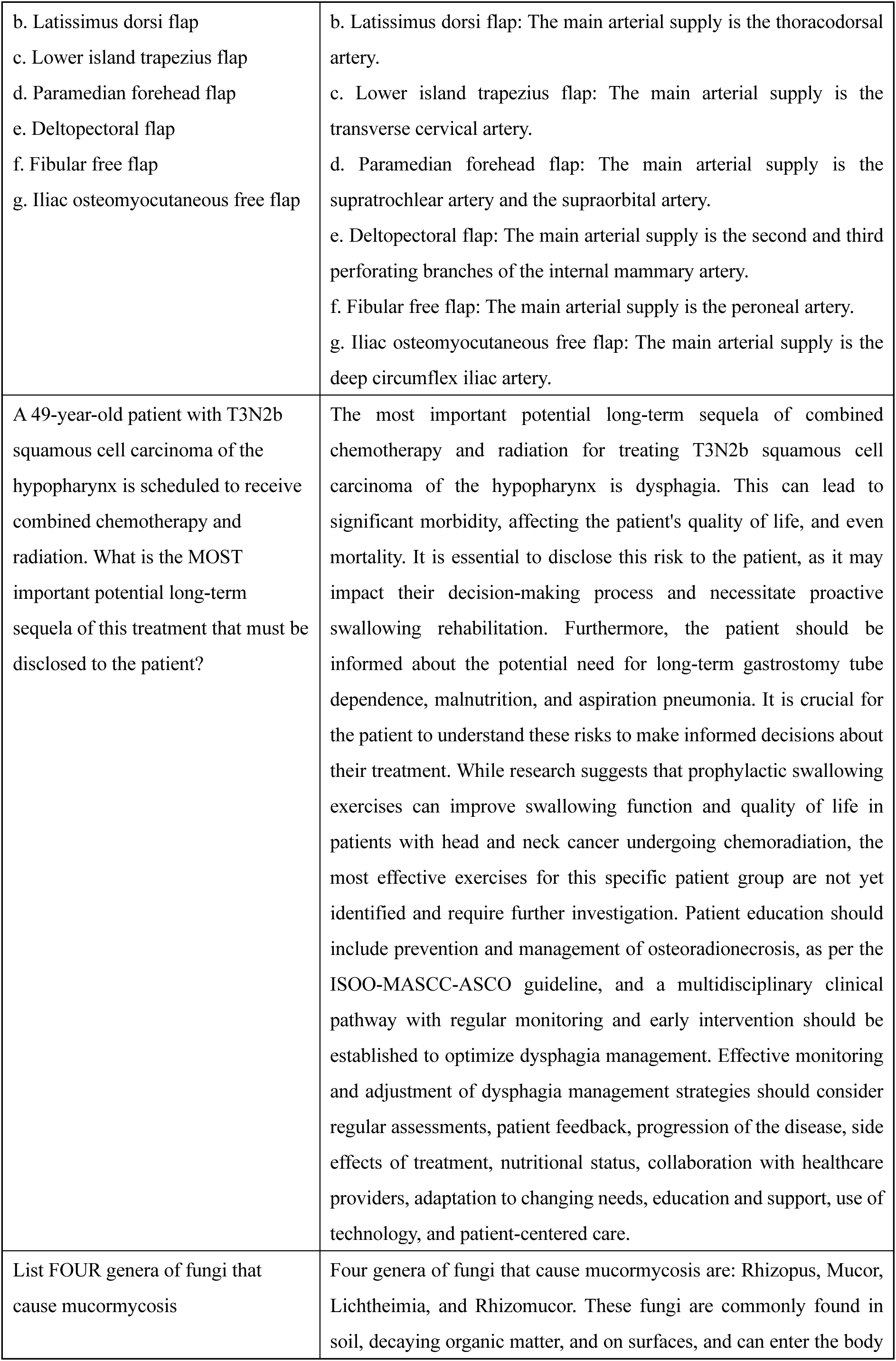
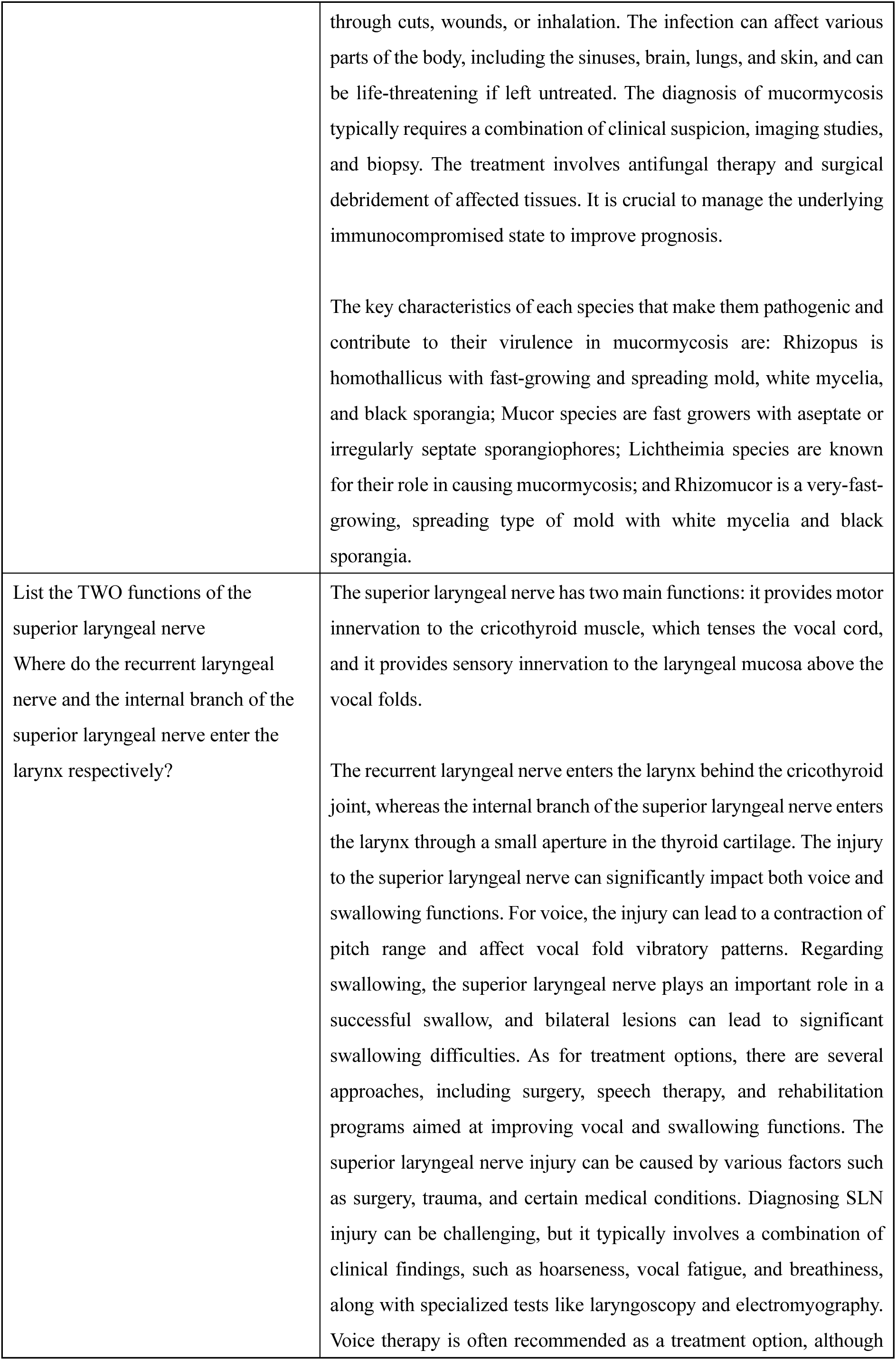
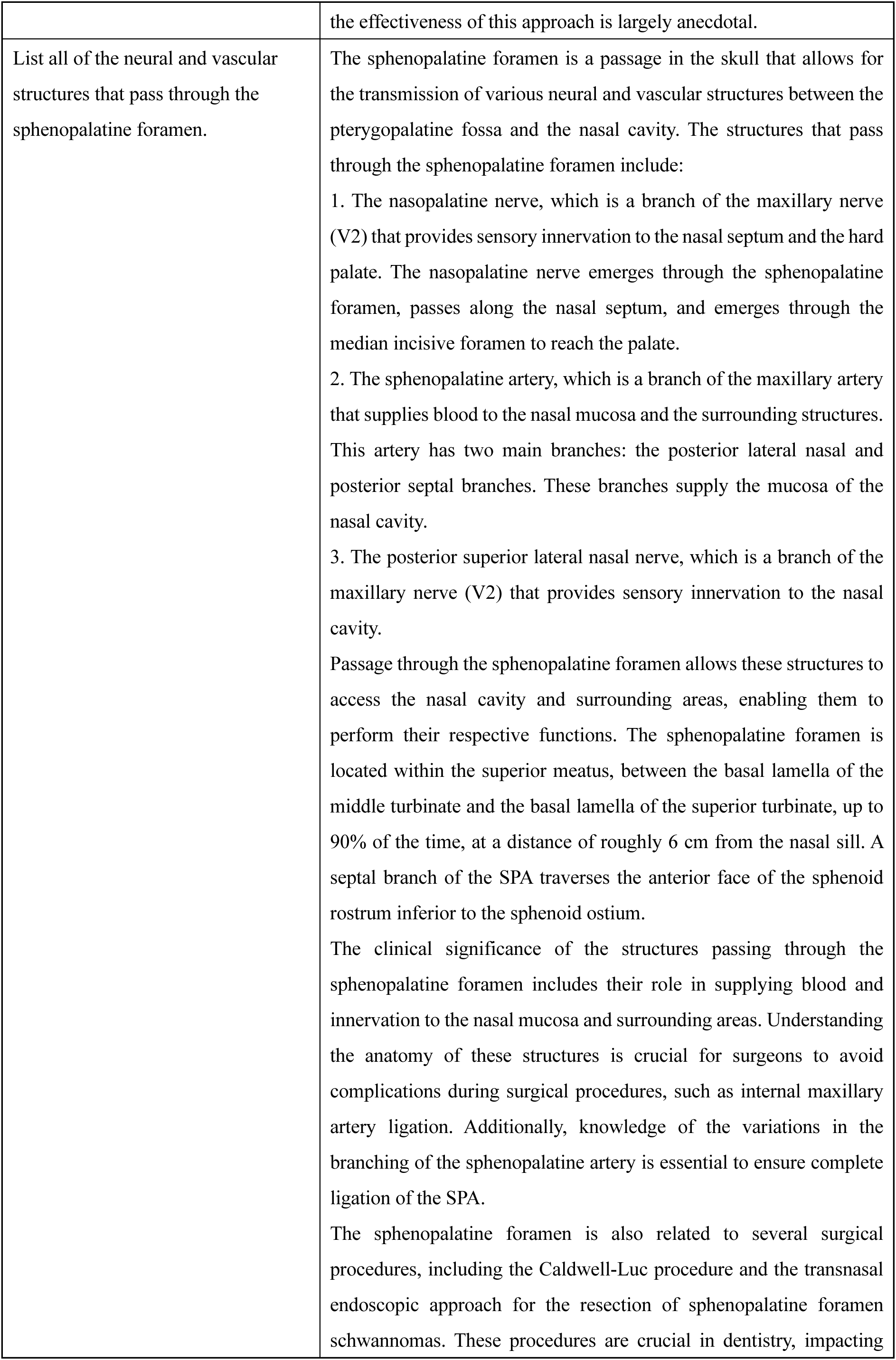
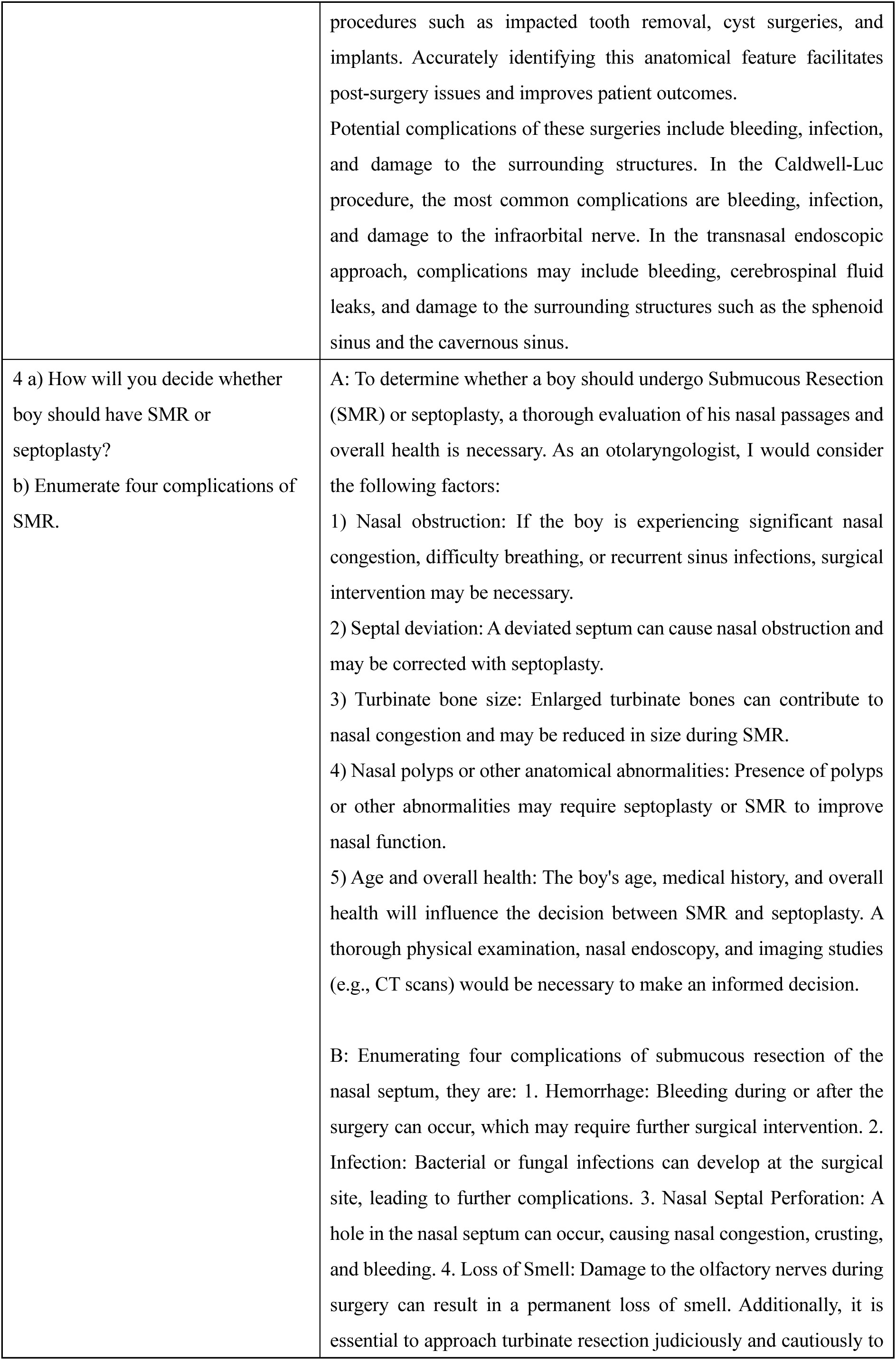
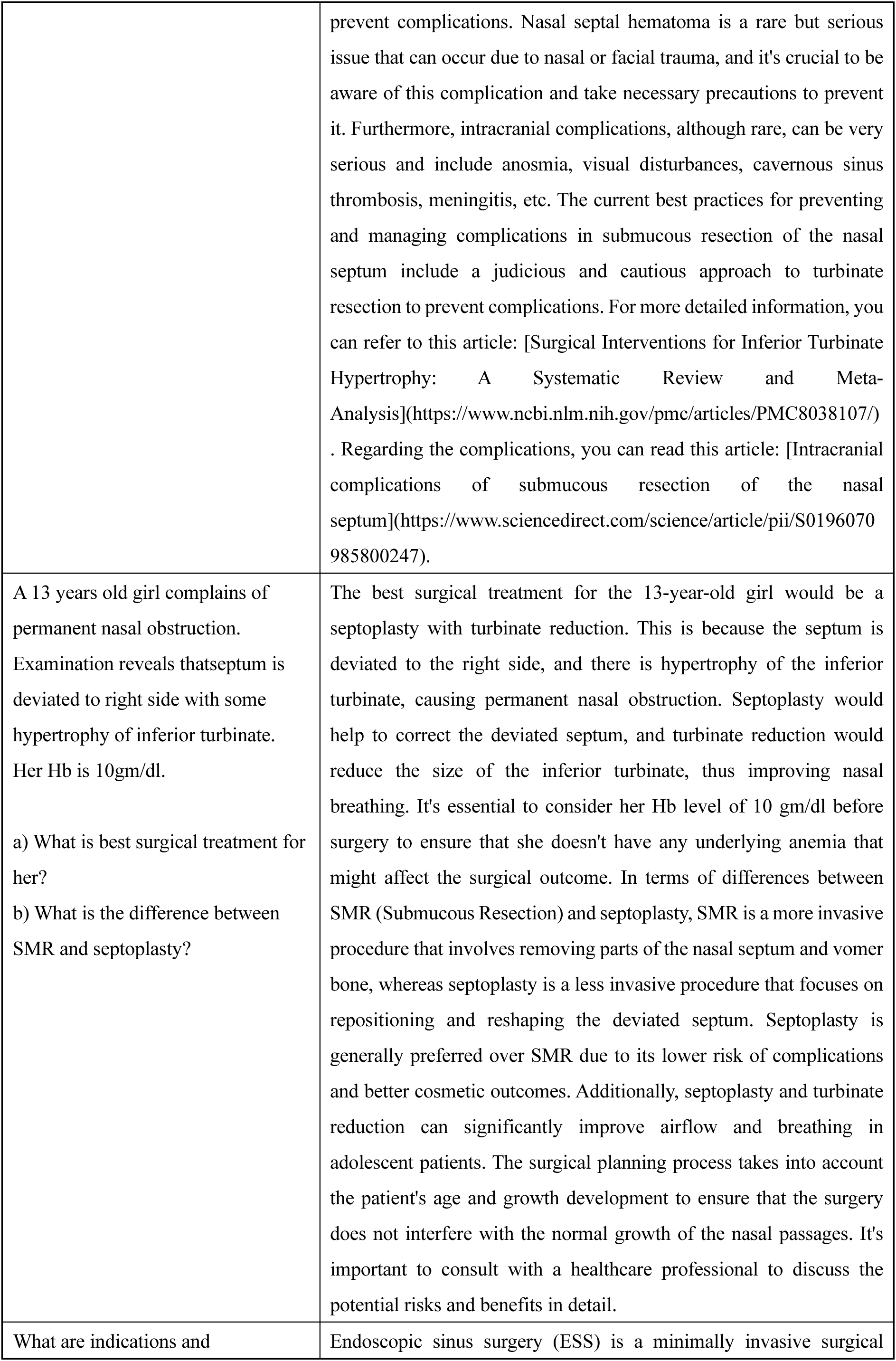
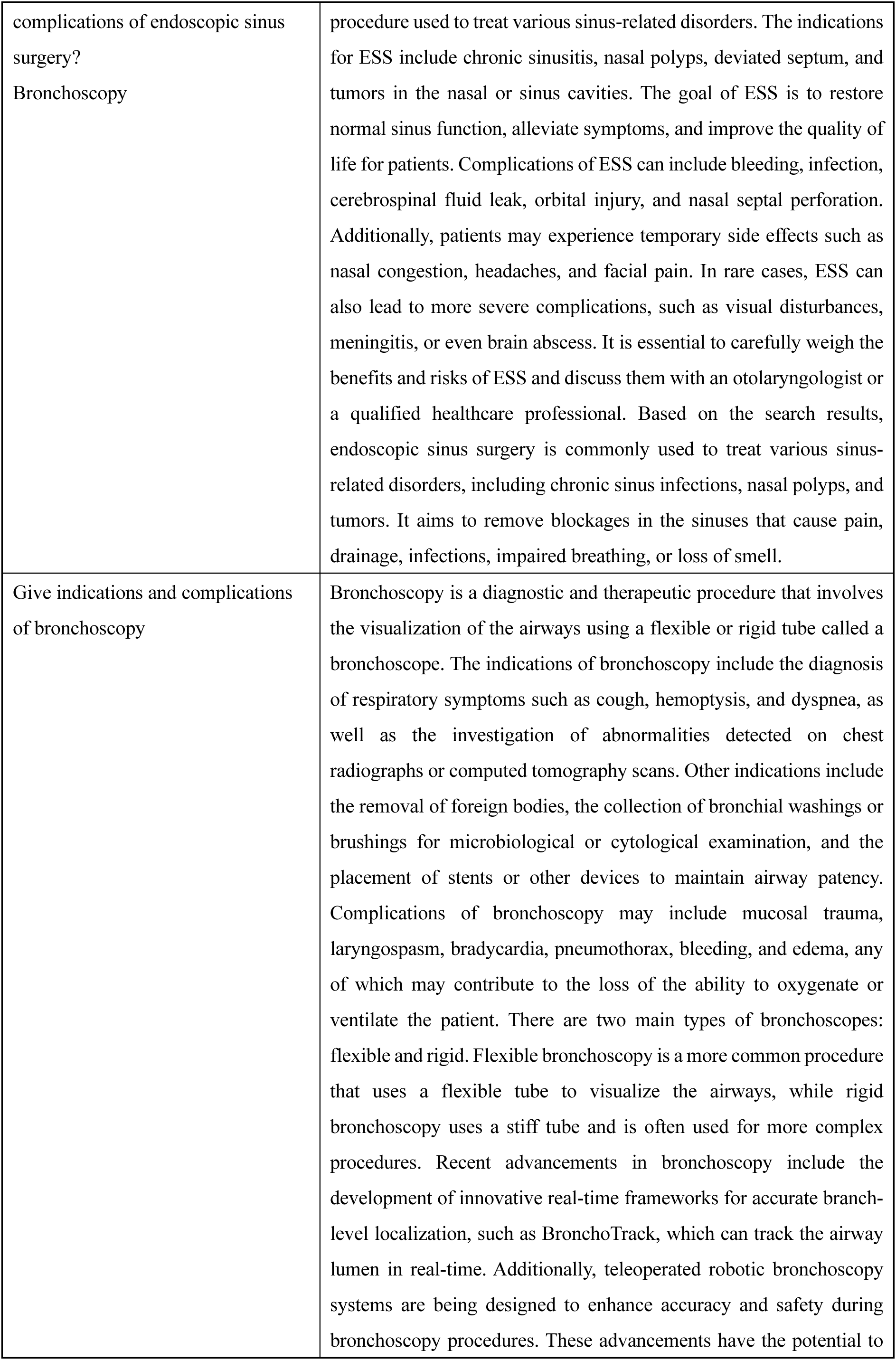
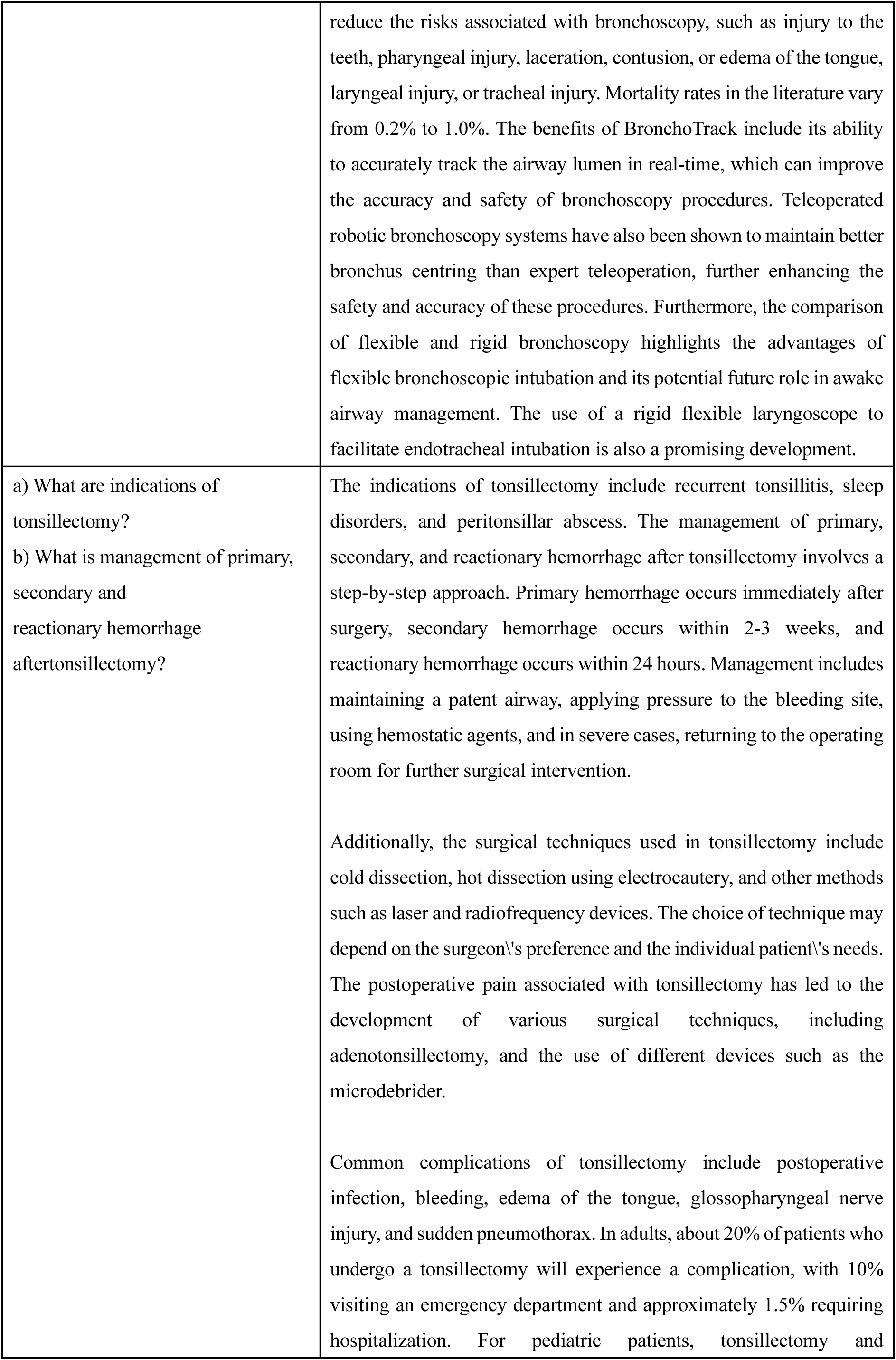
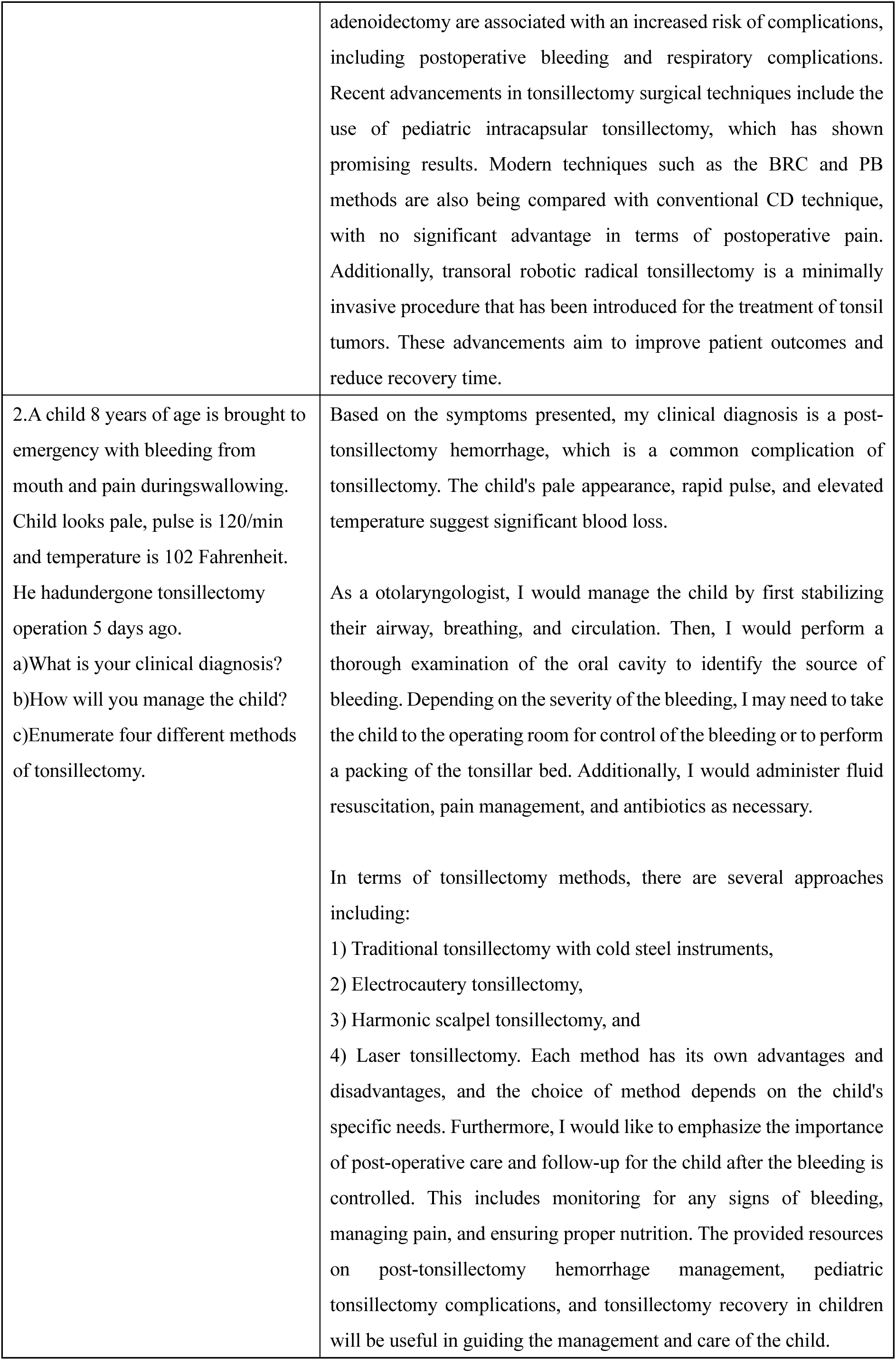
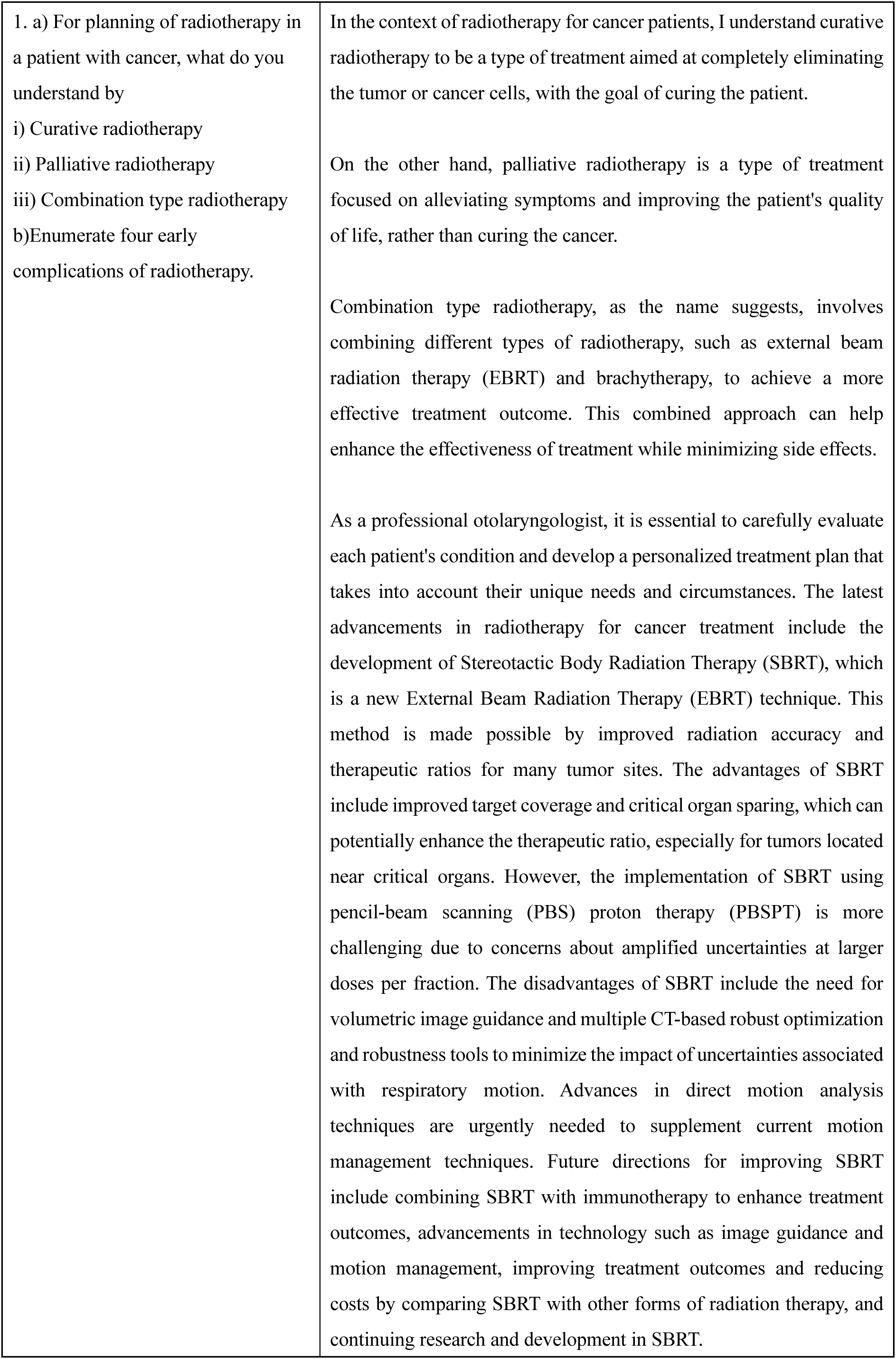
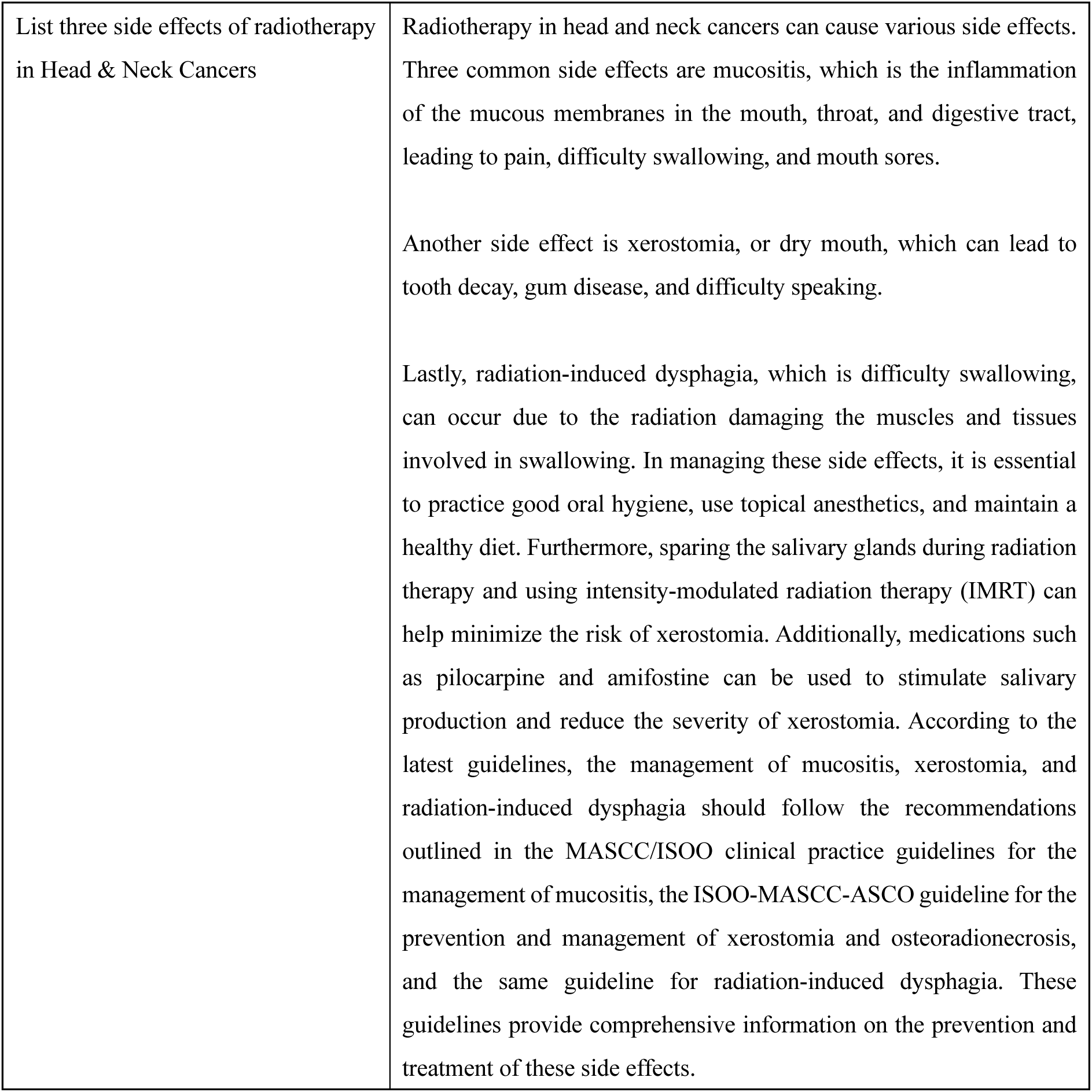
Short questions and answers from ENTAgent.

**Table 2:**
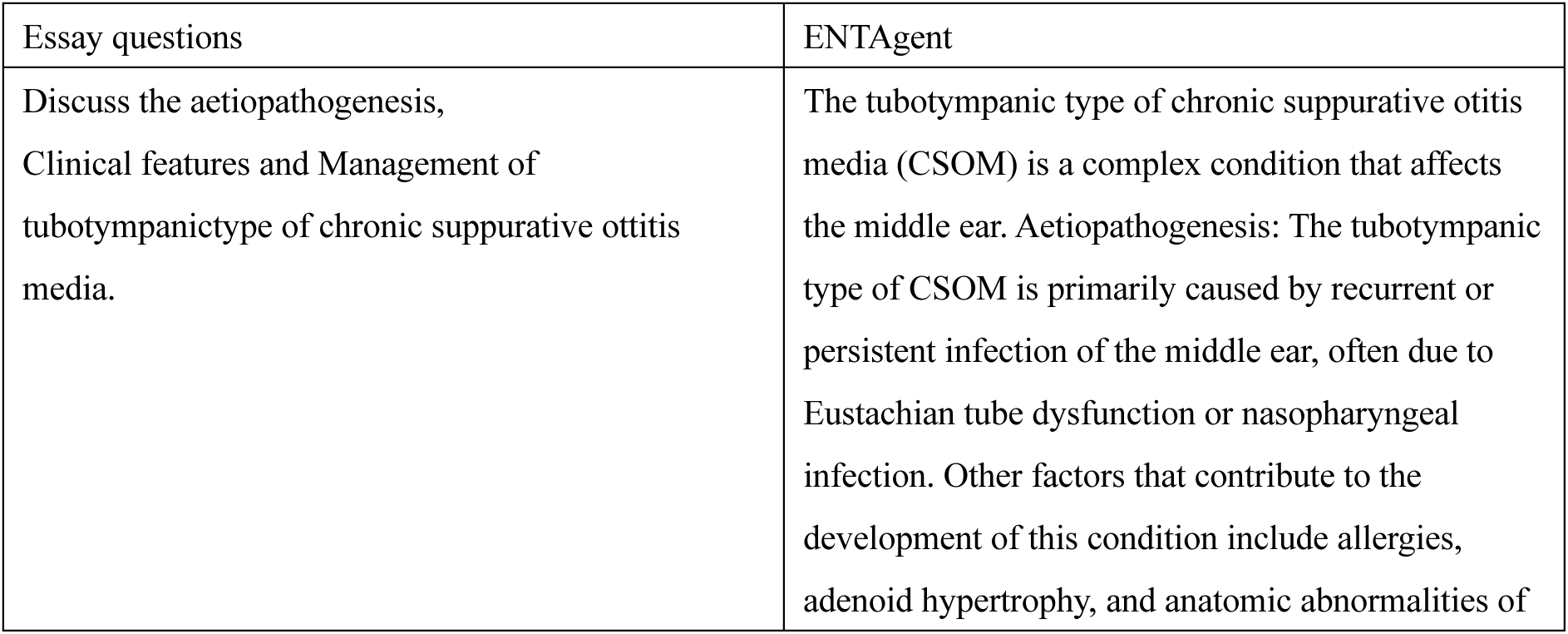
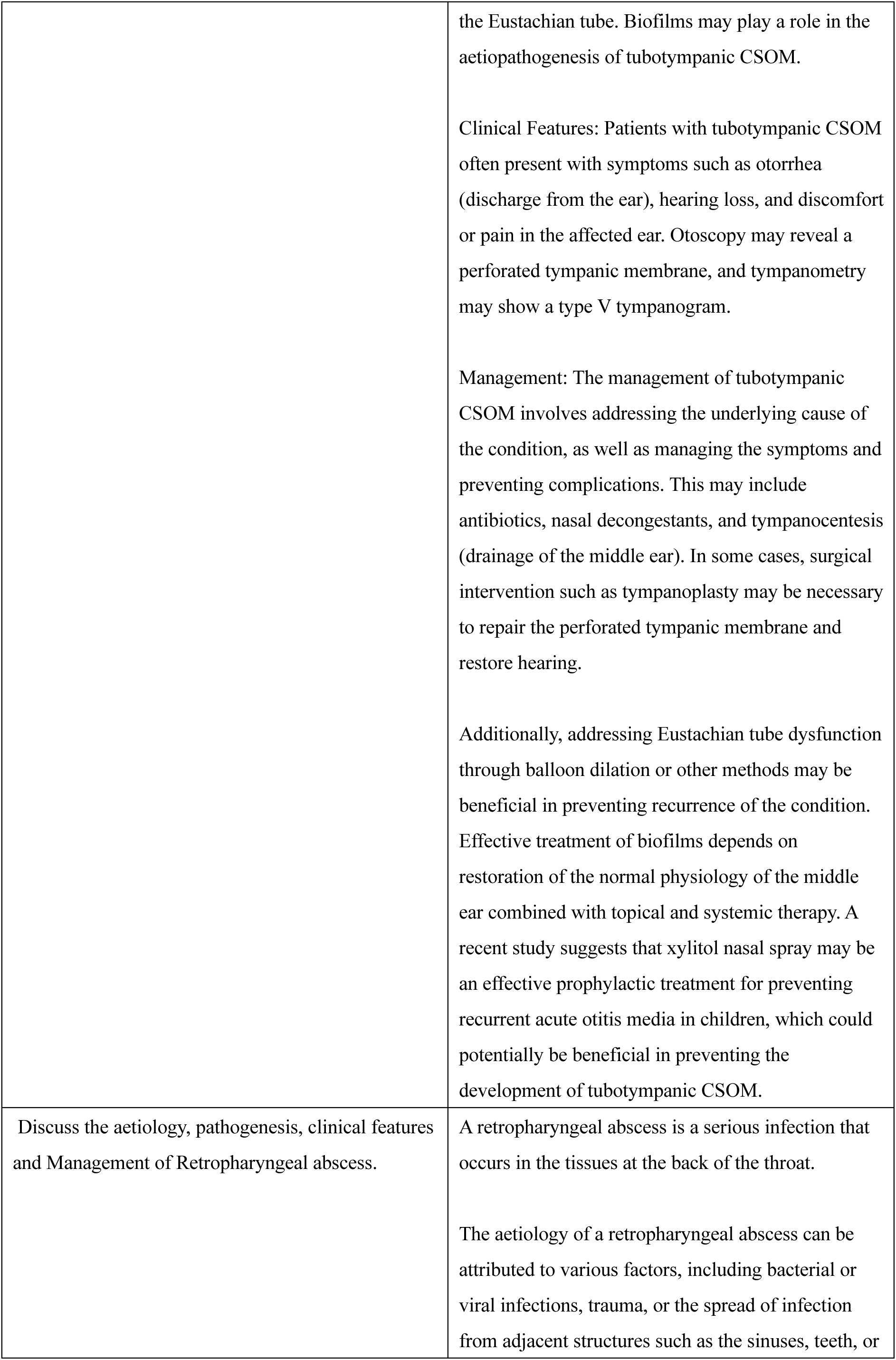
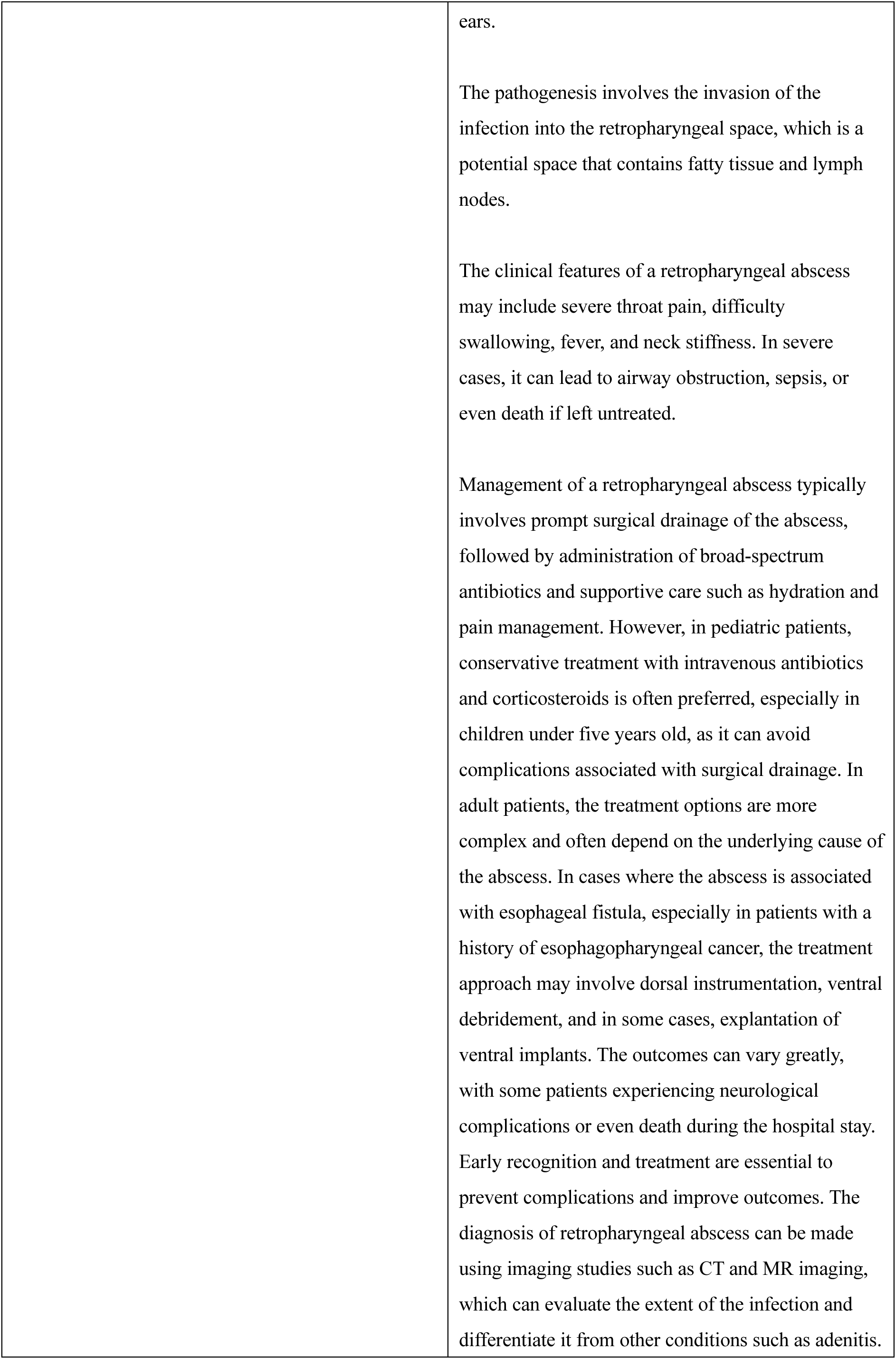
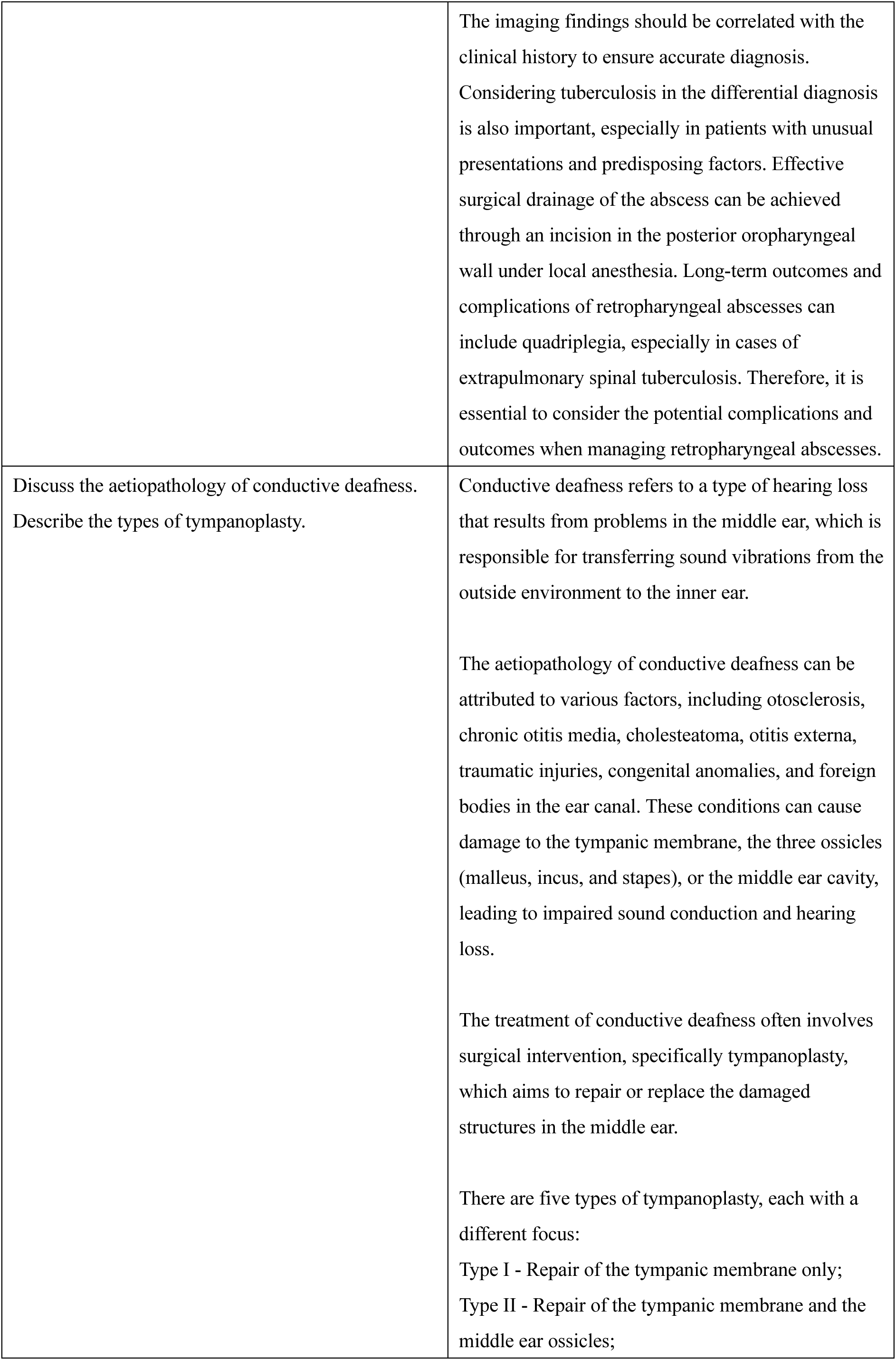
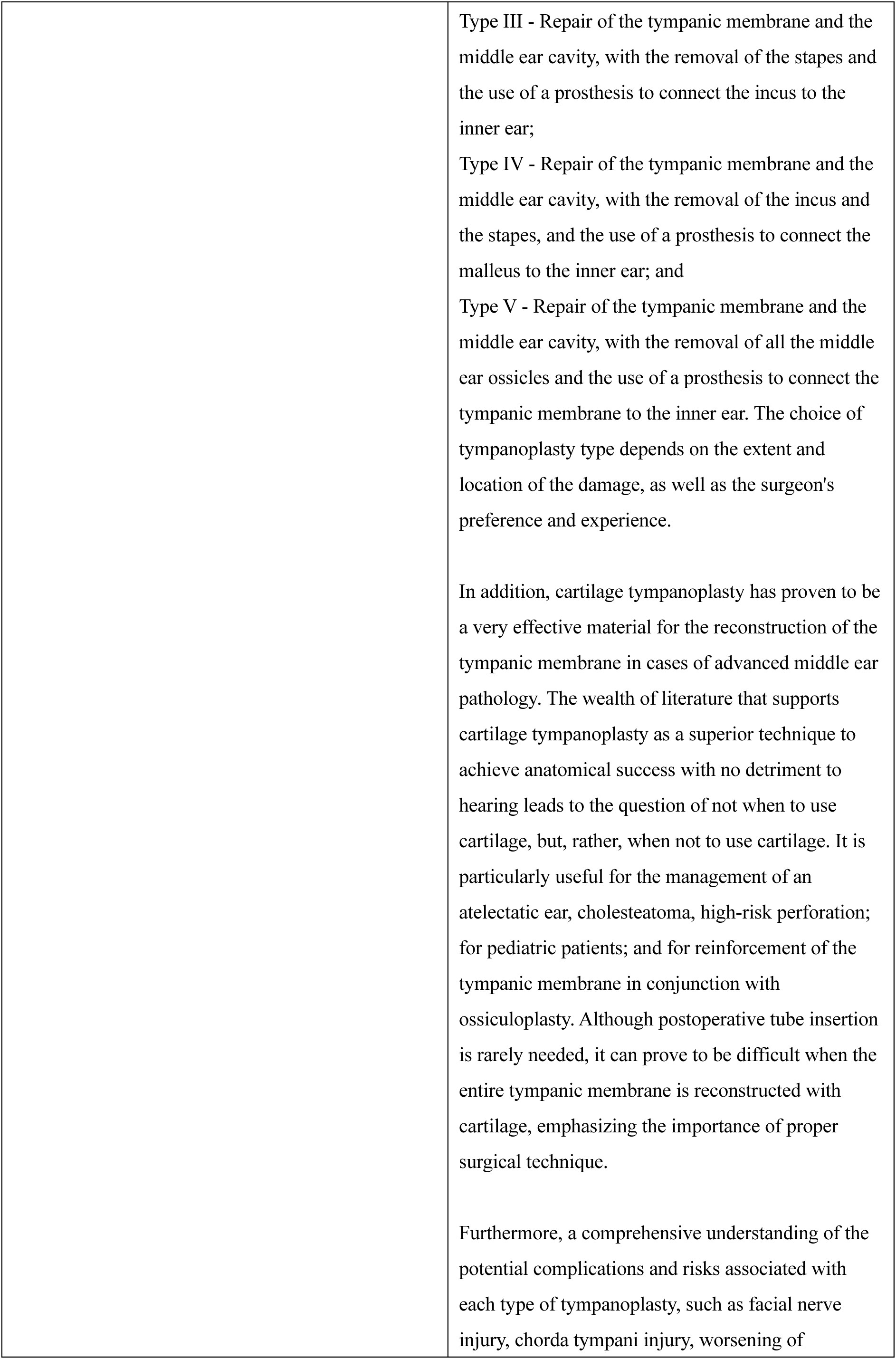
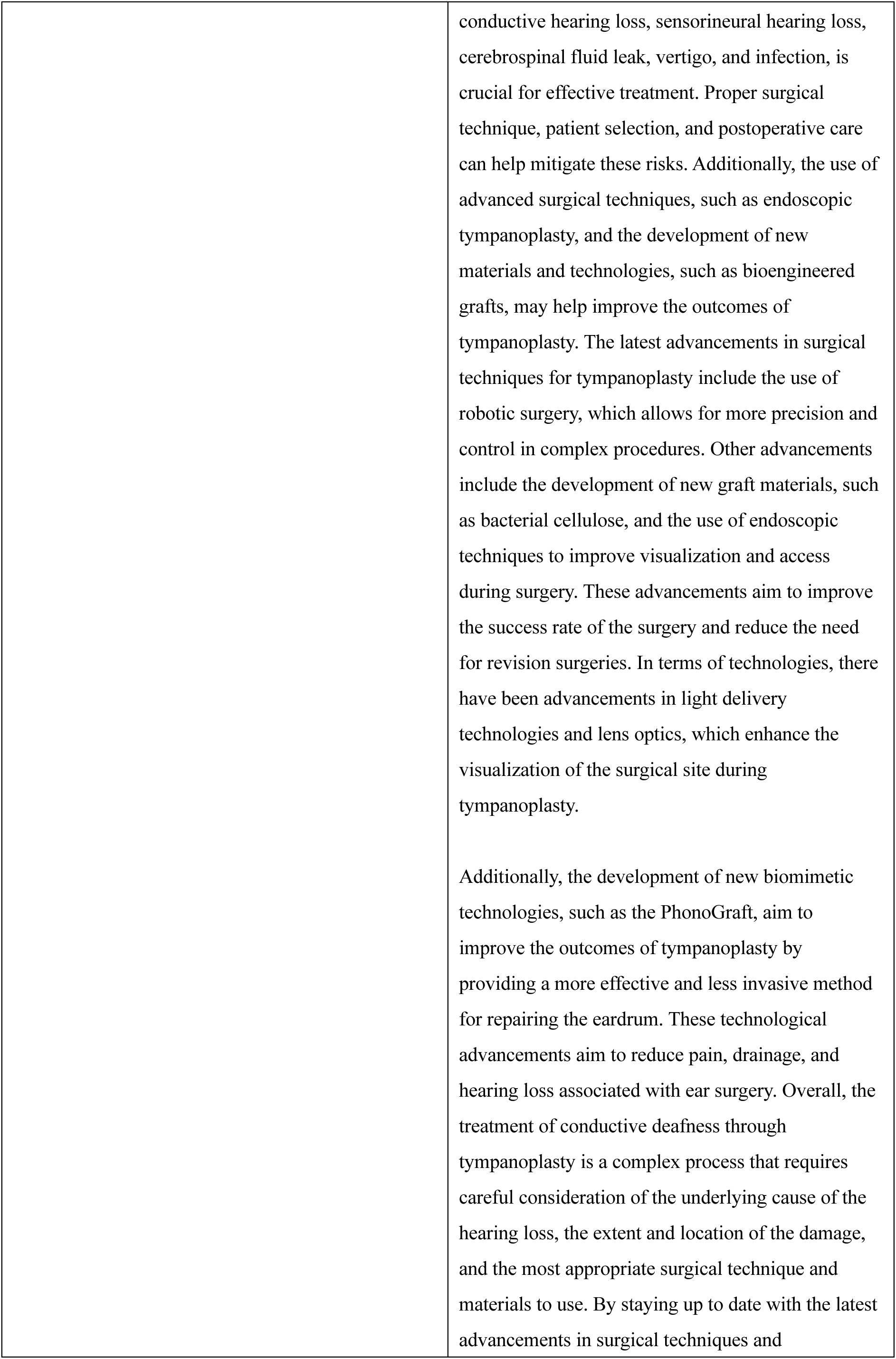
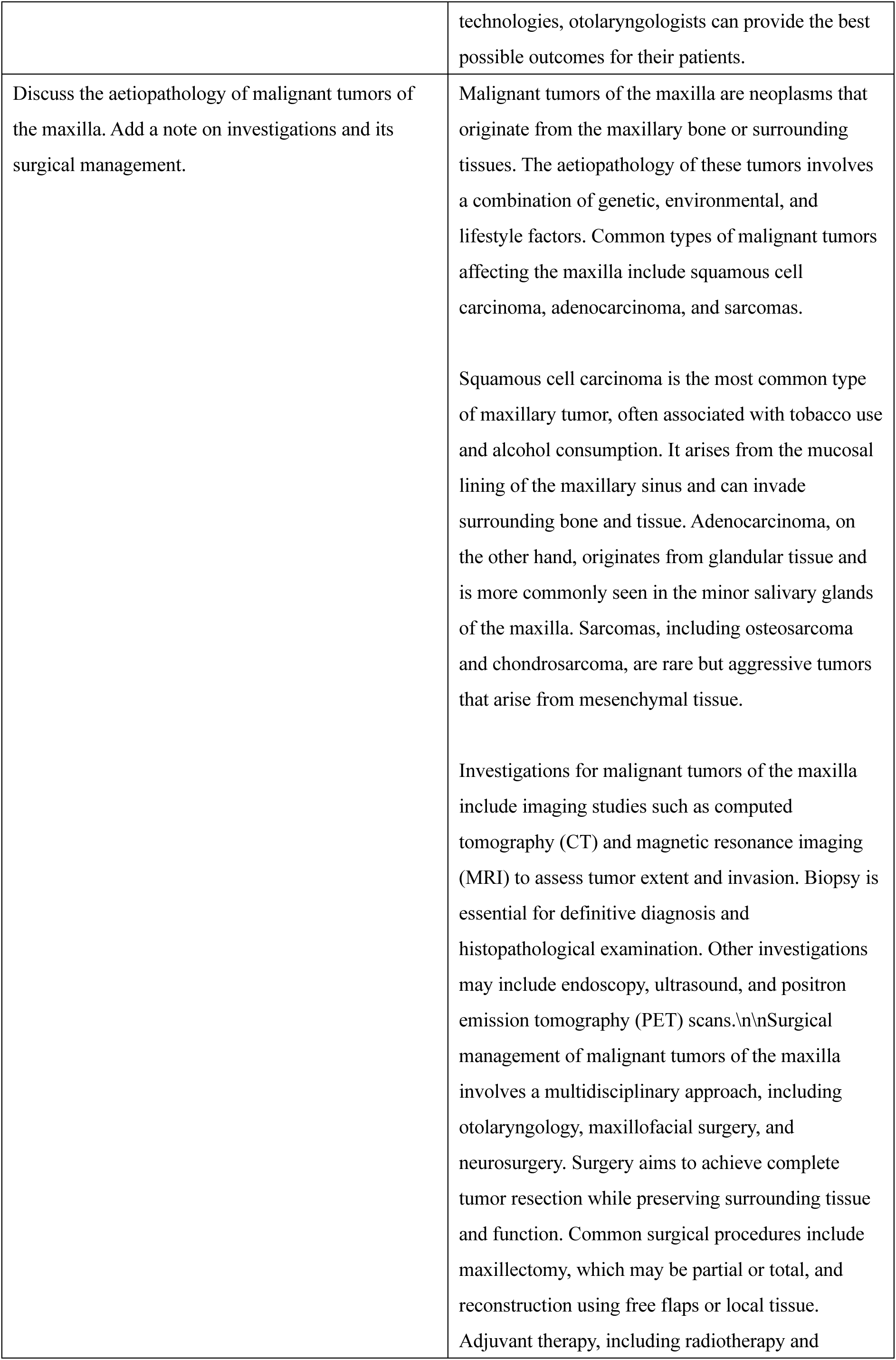
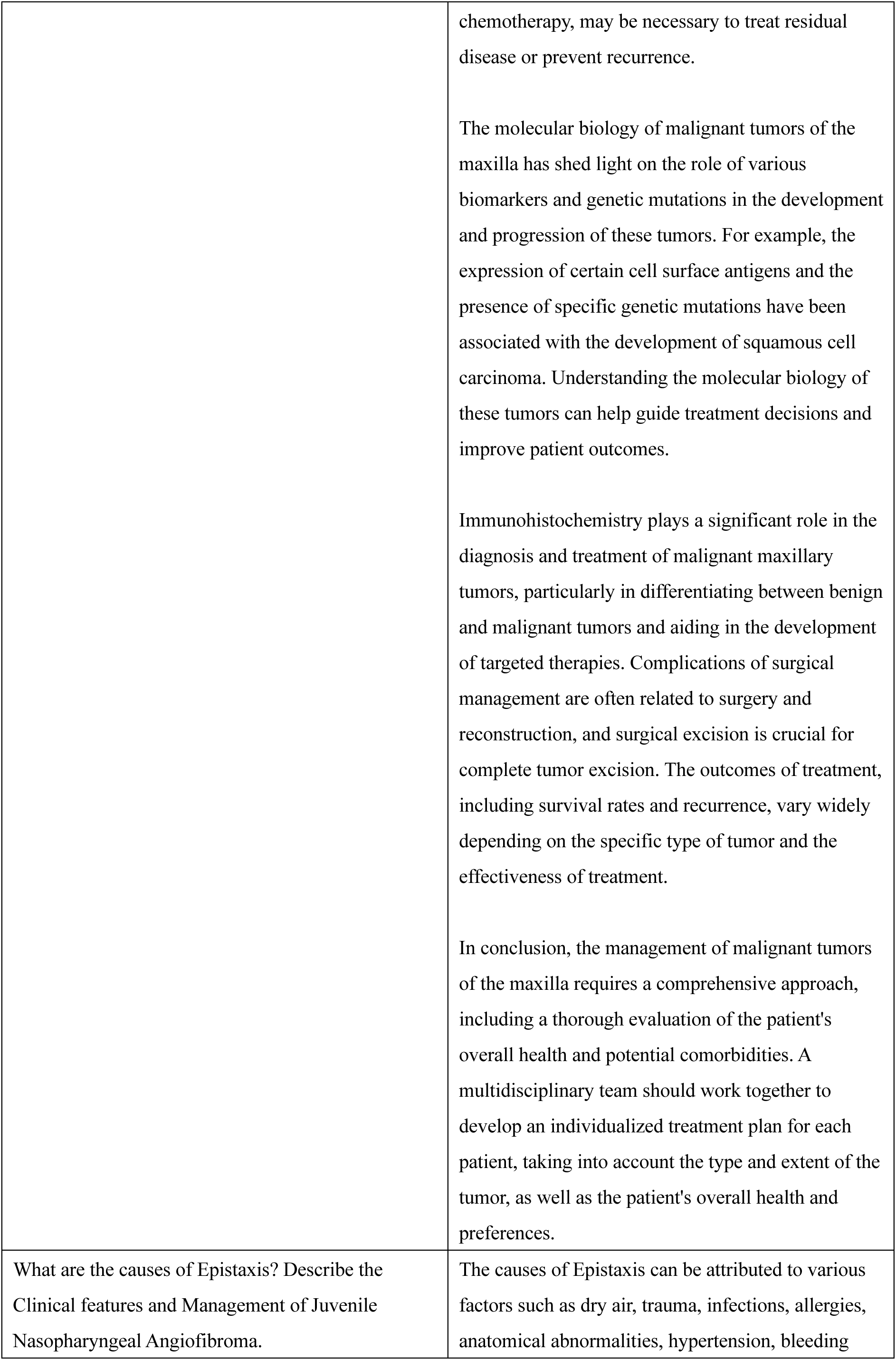
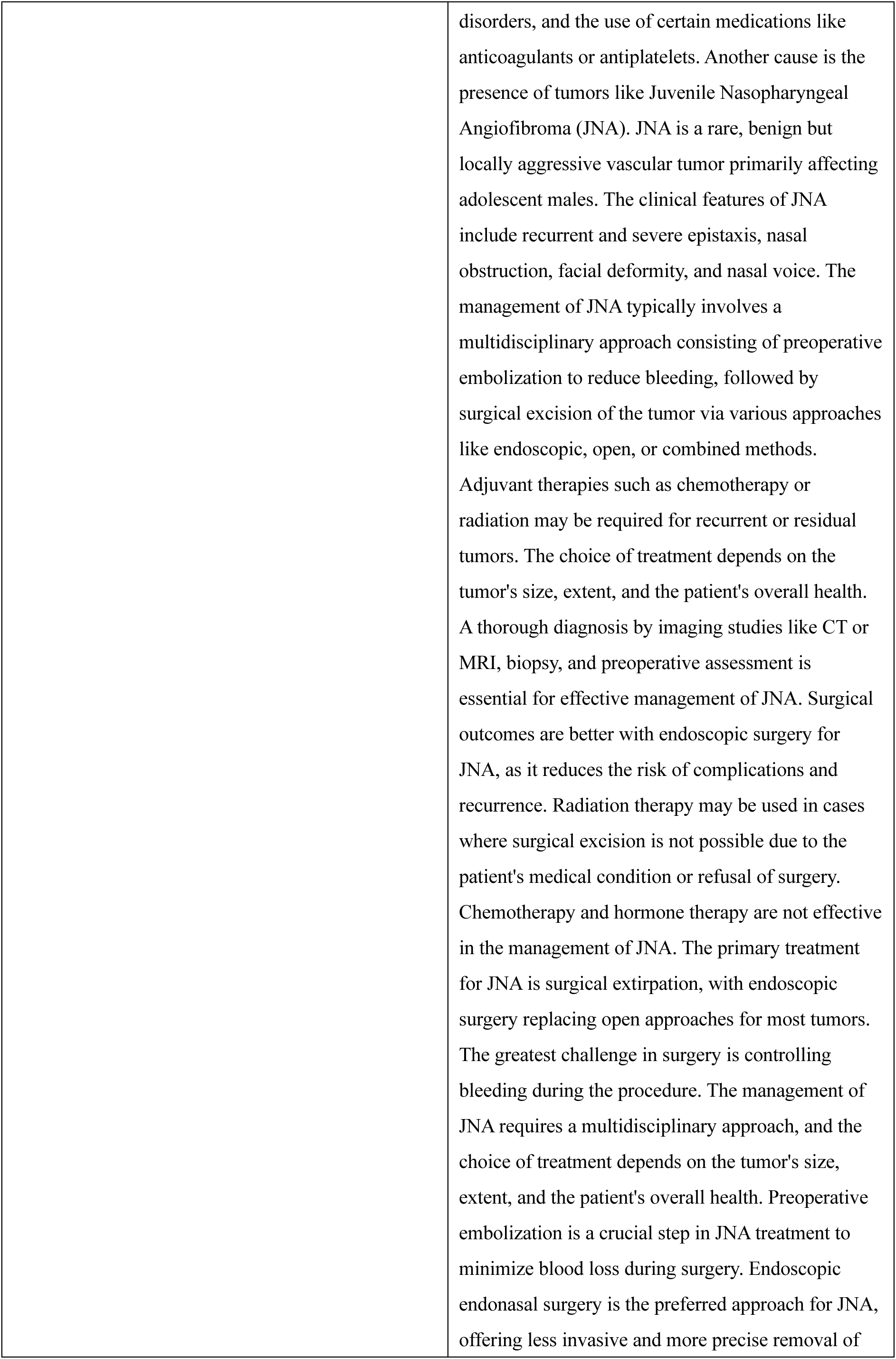
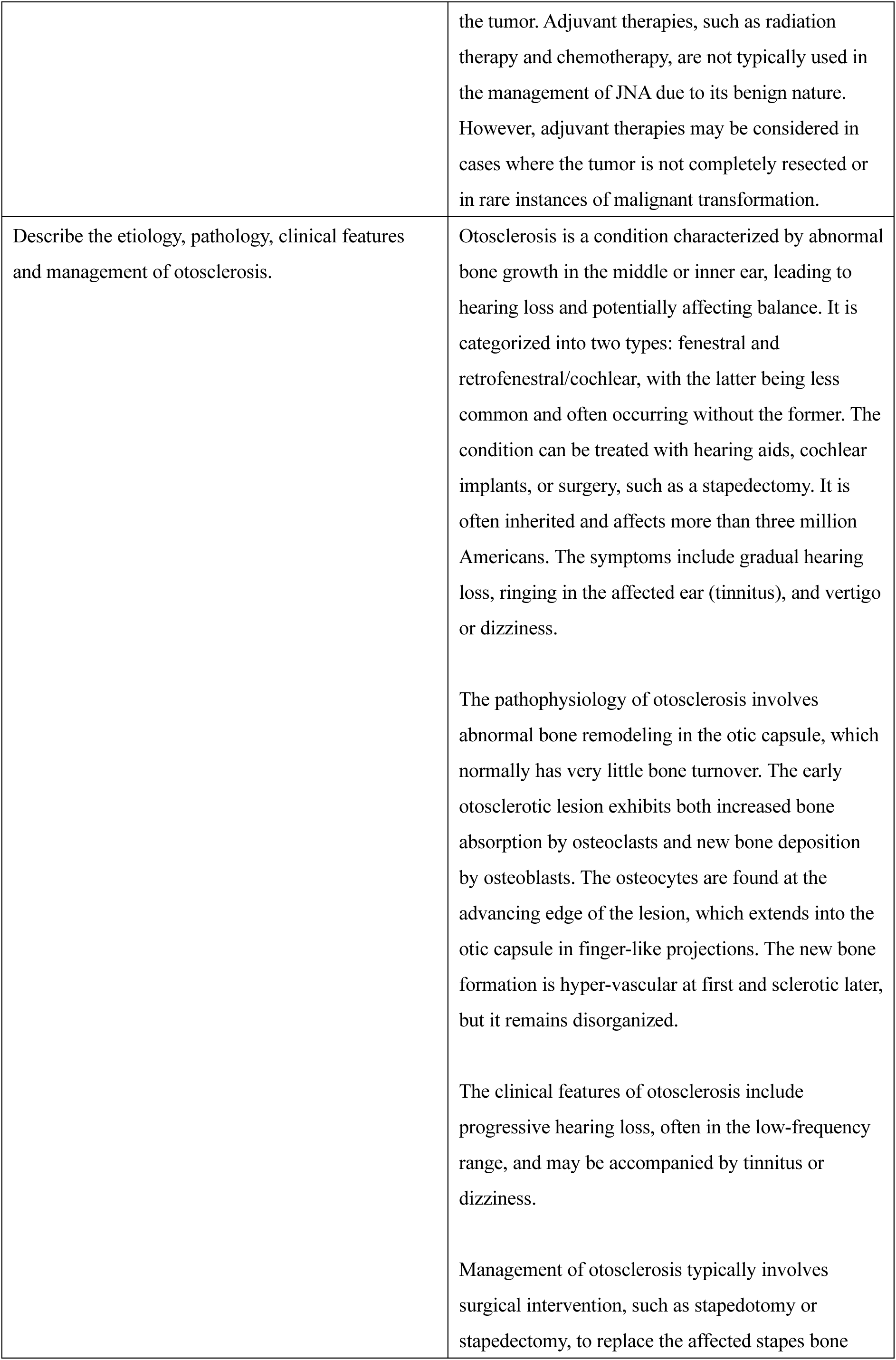
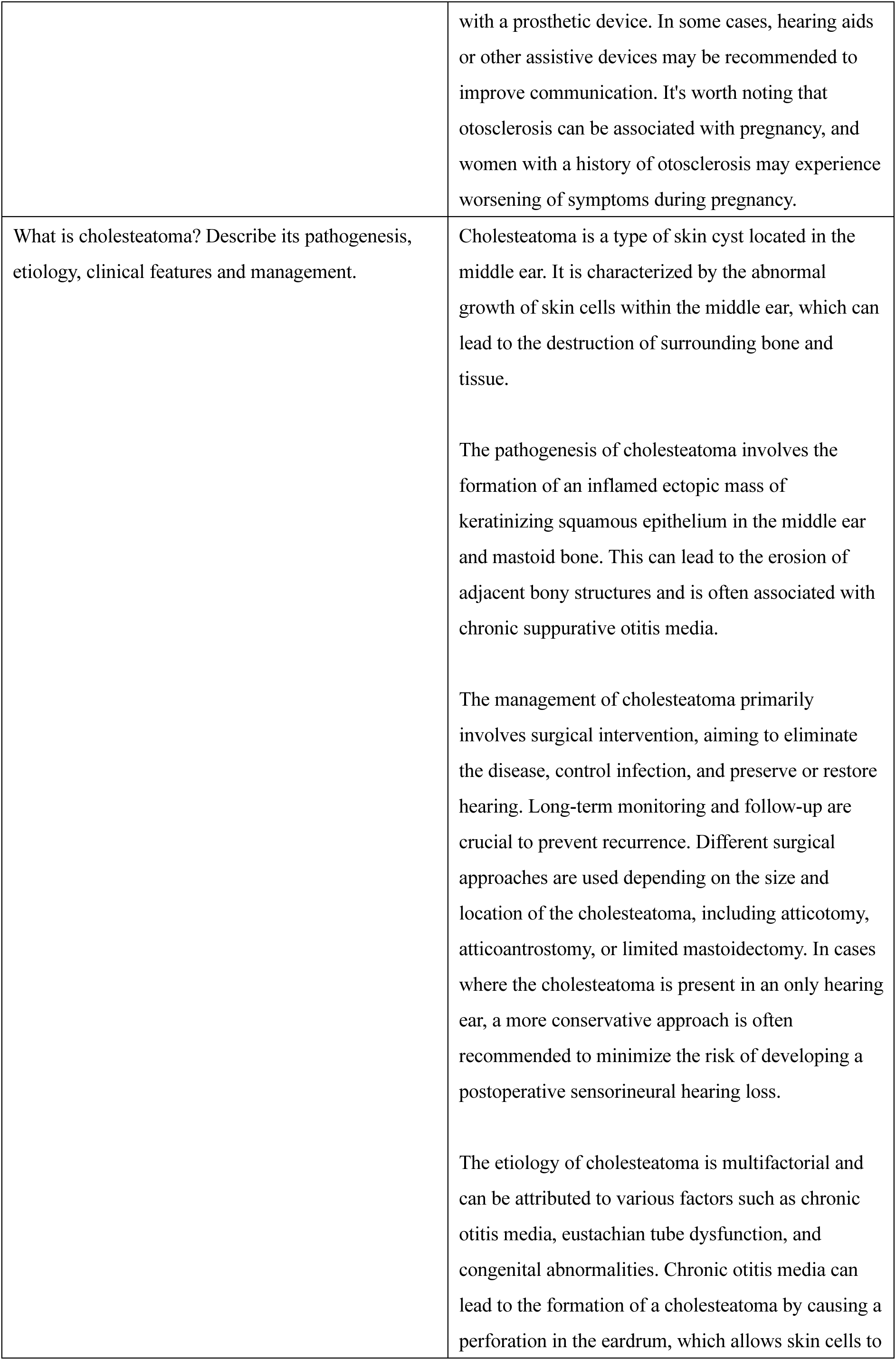
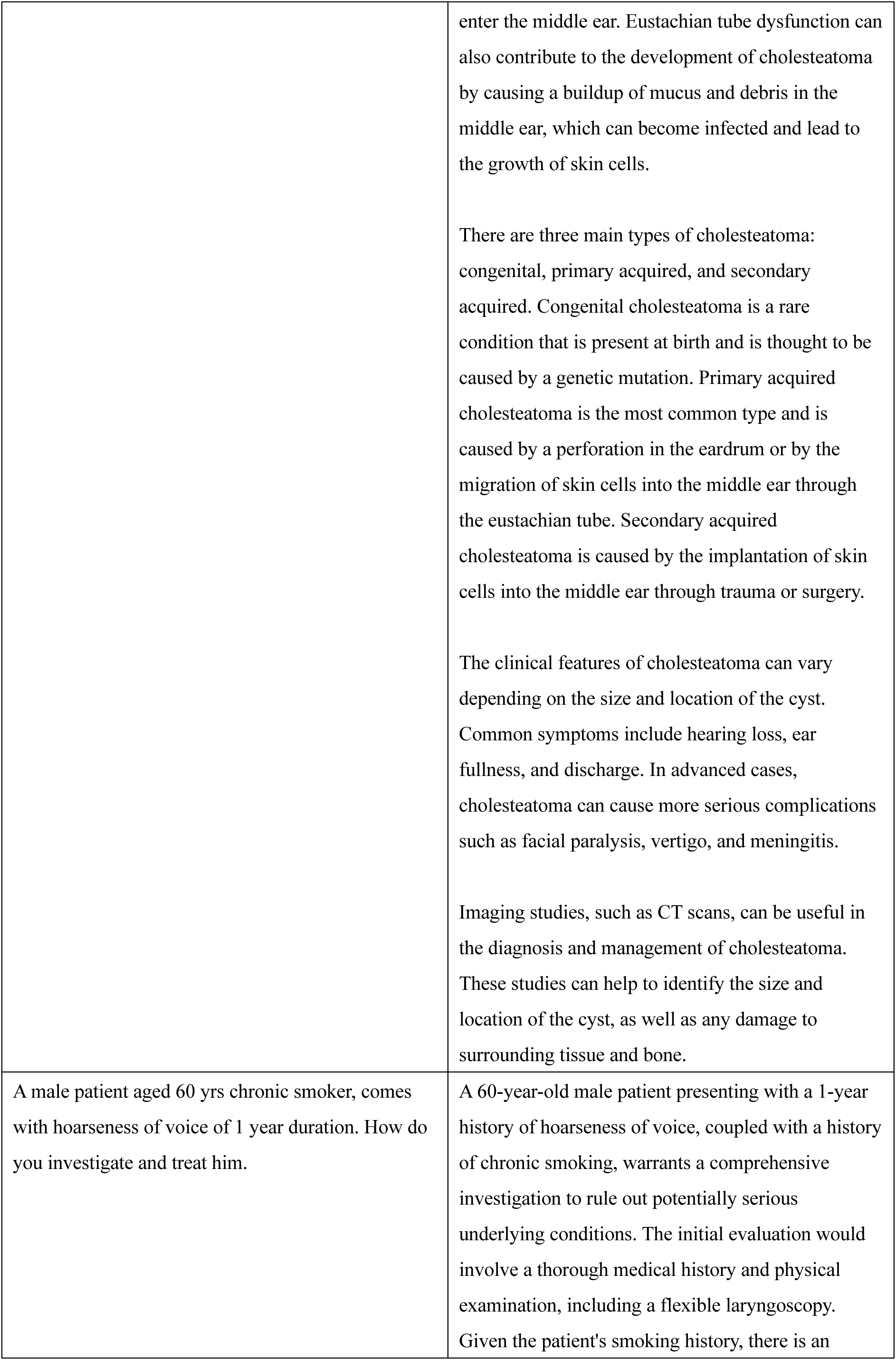
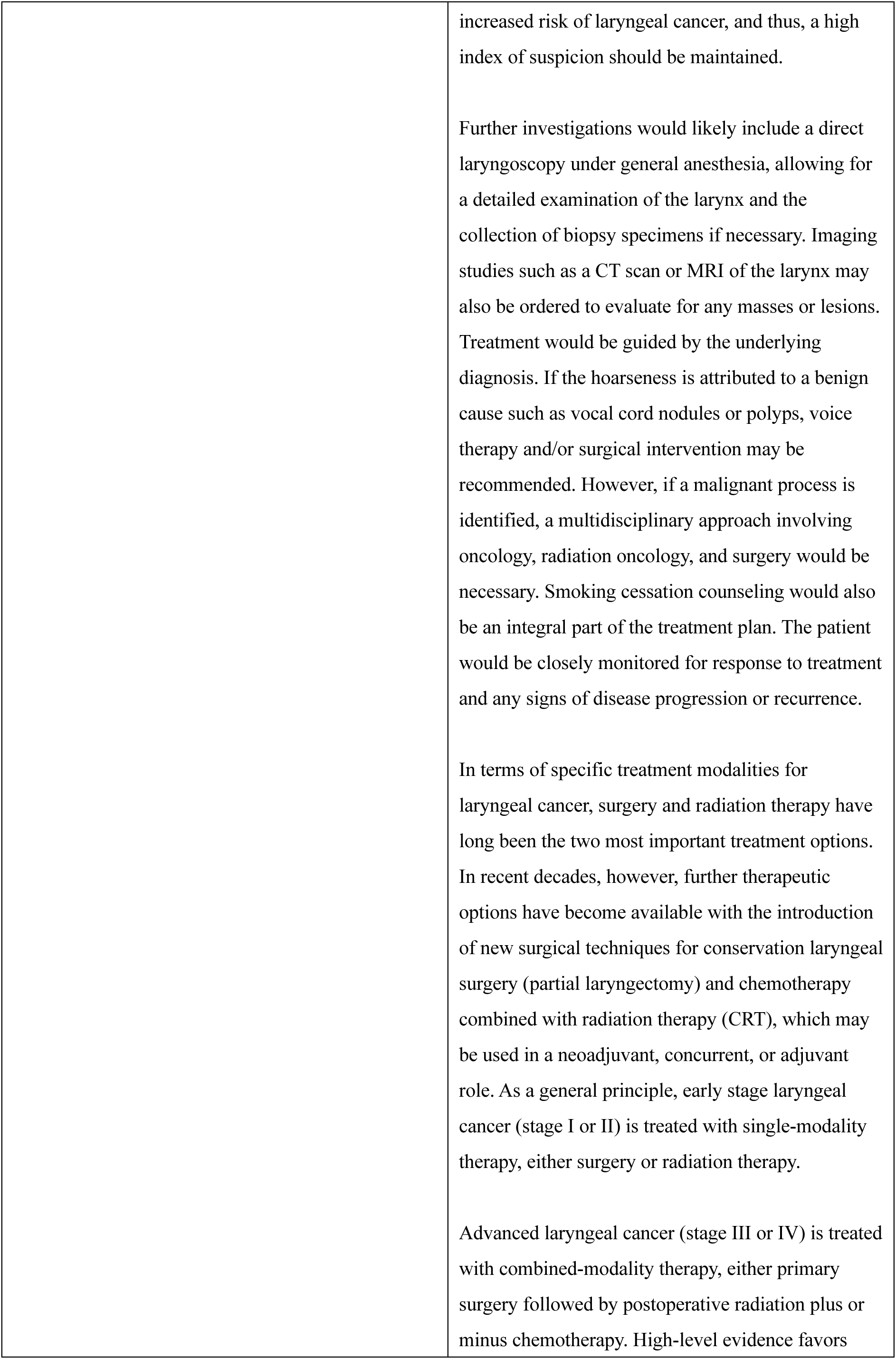
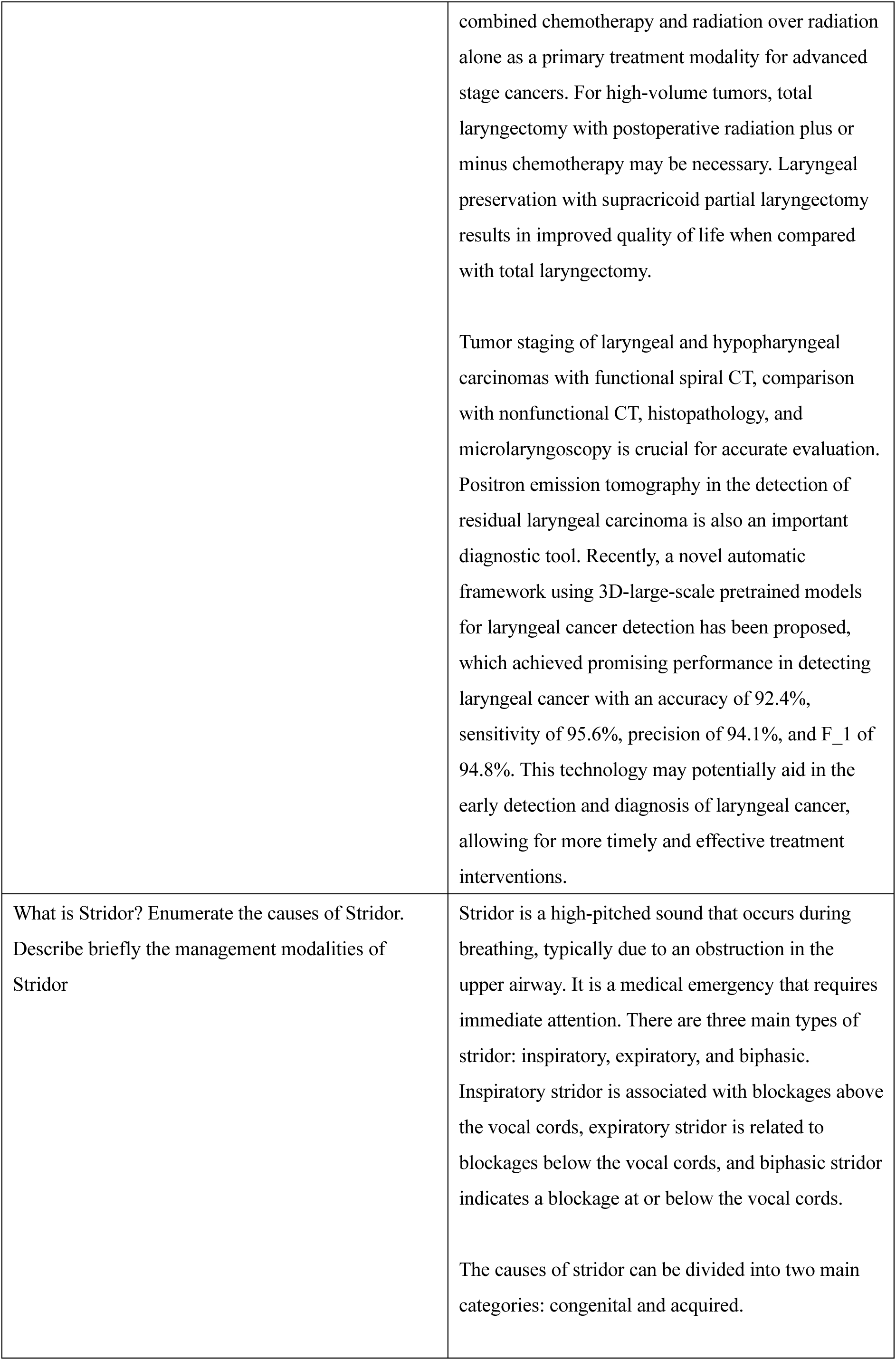
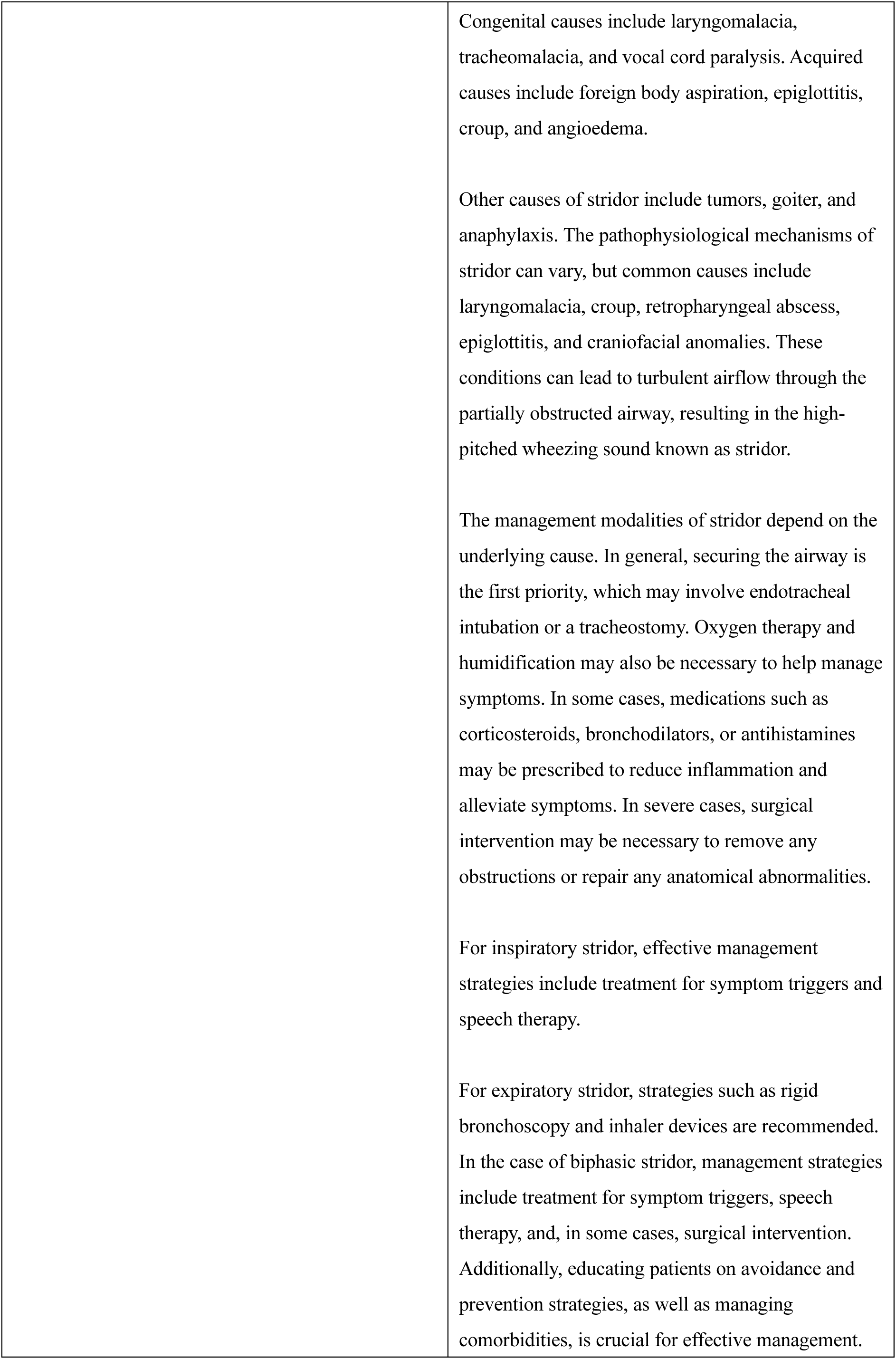
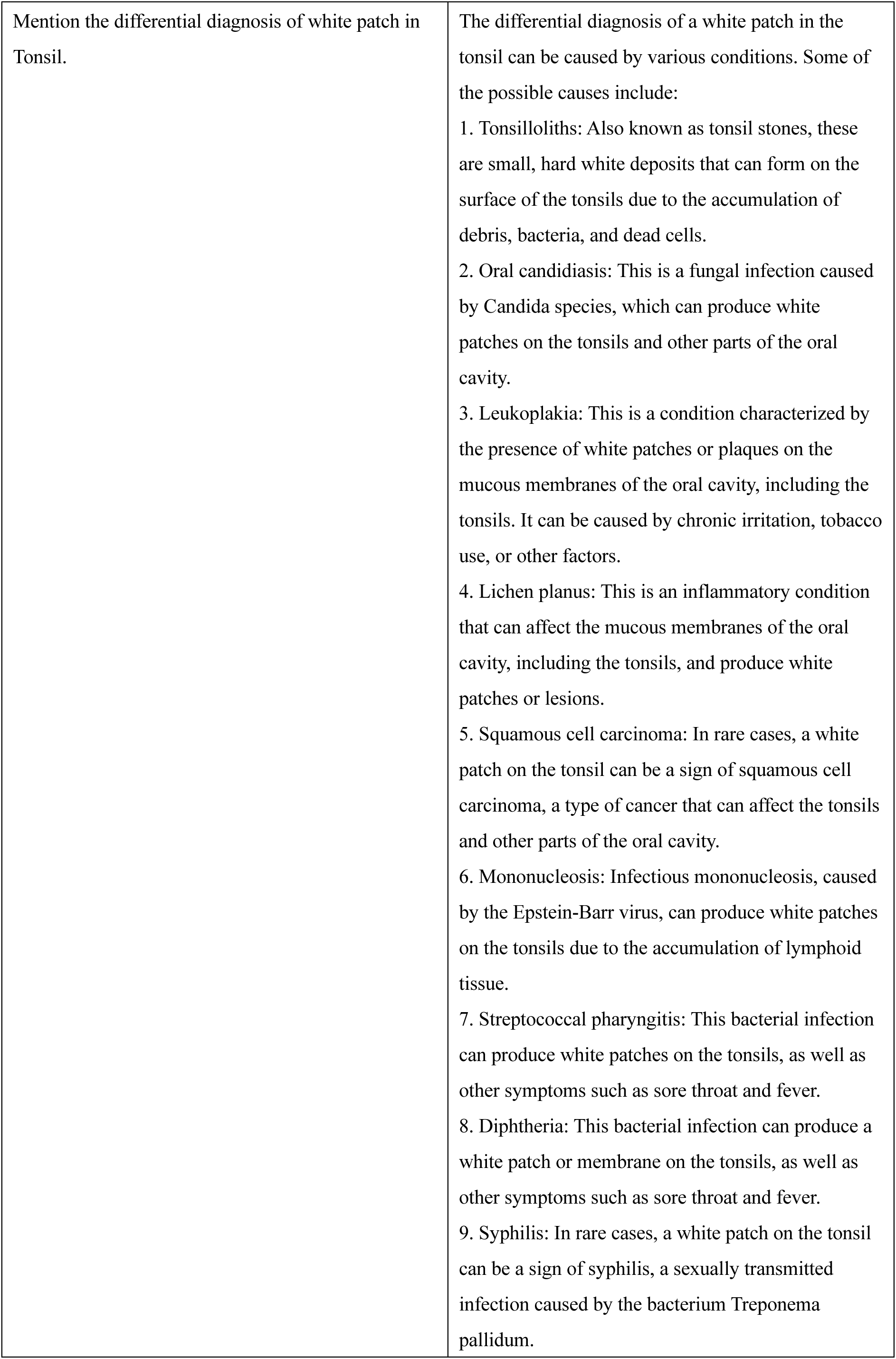
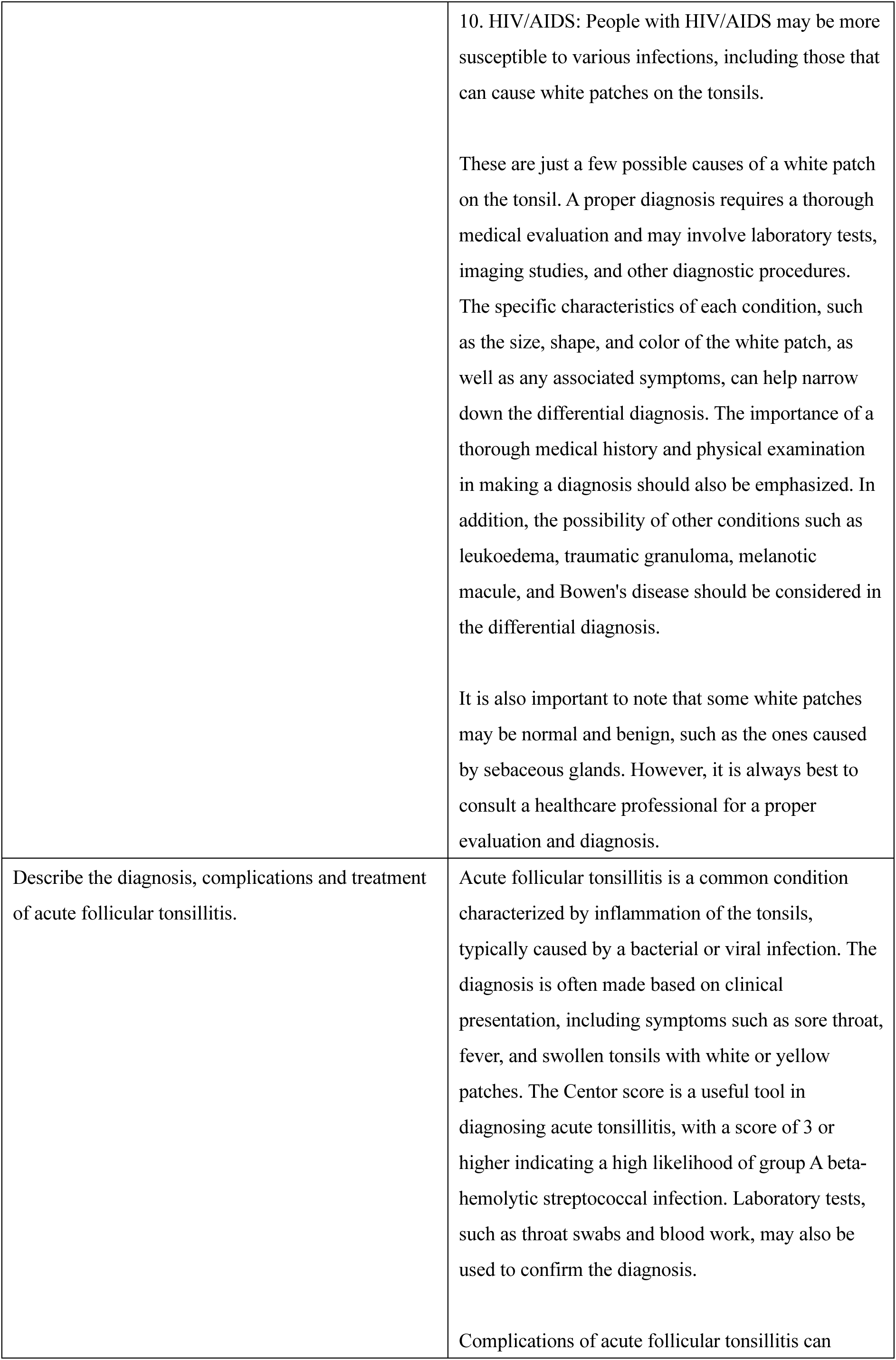
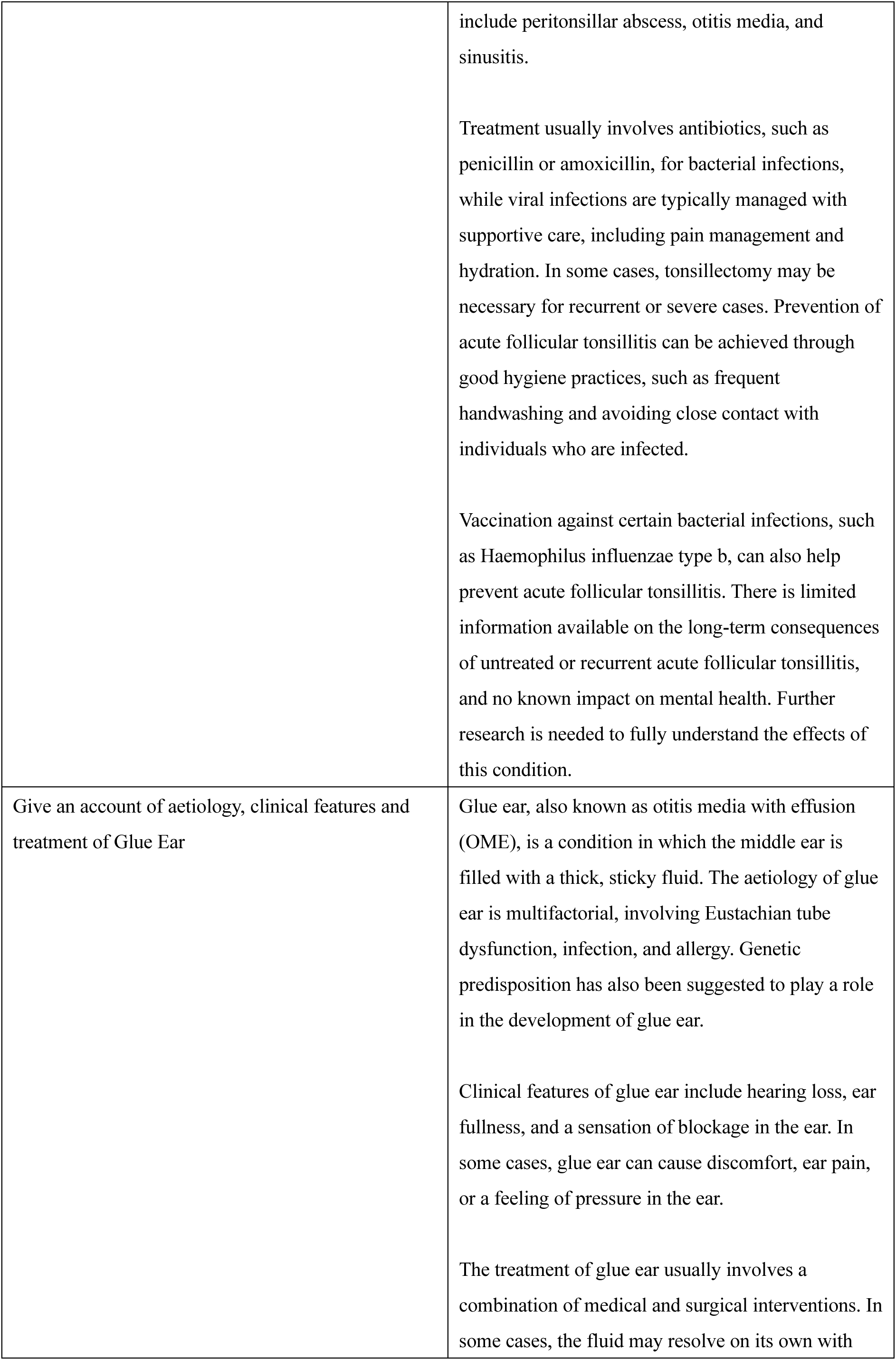
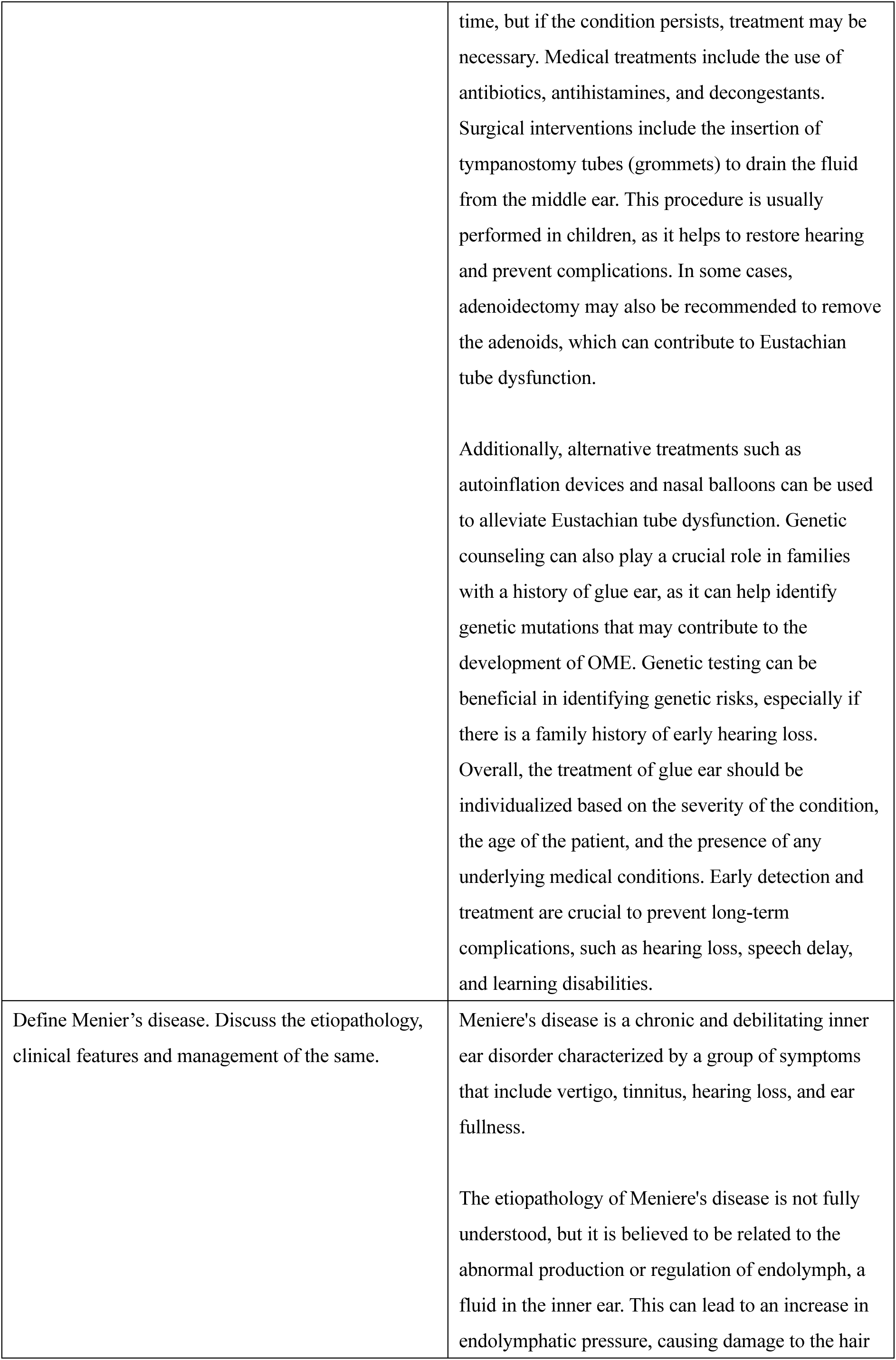
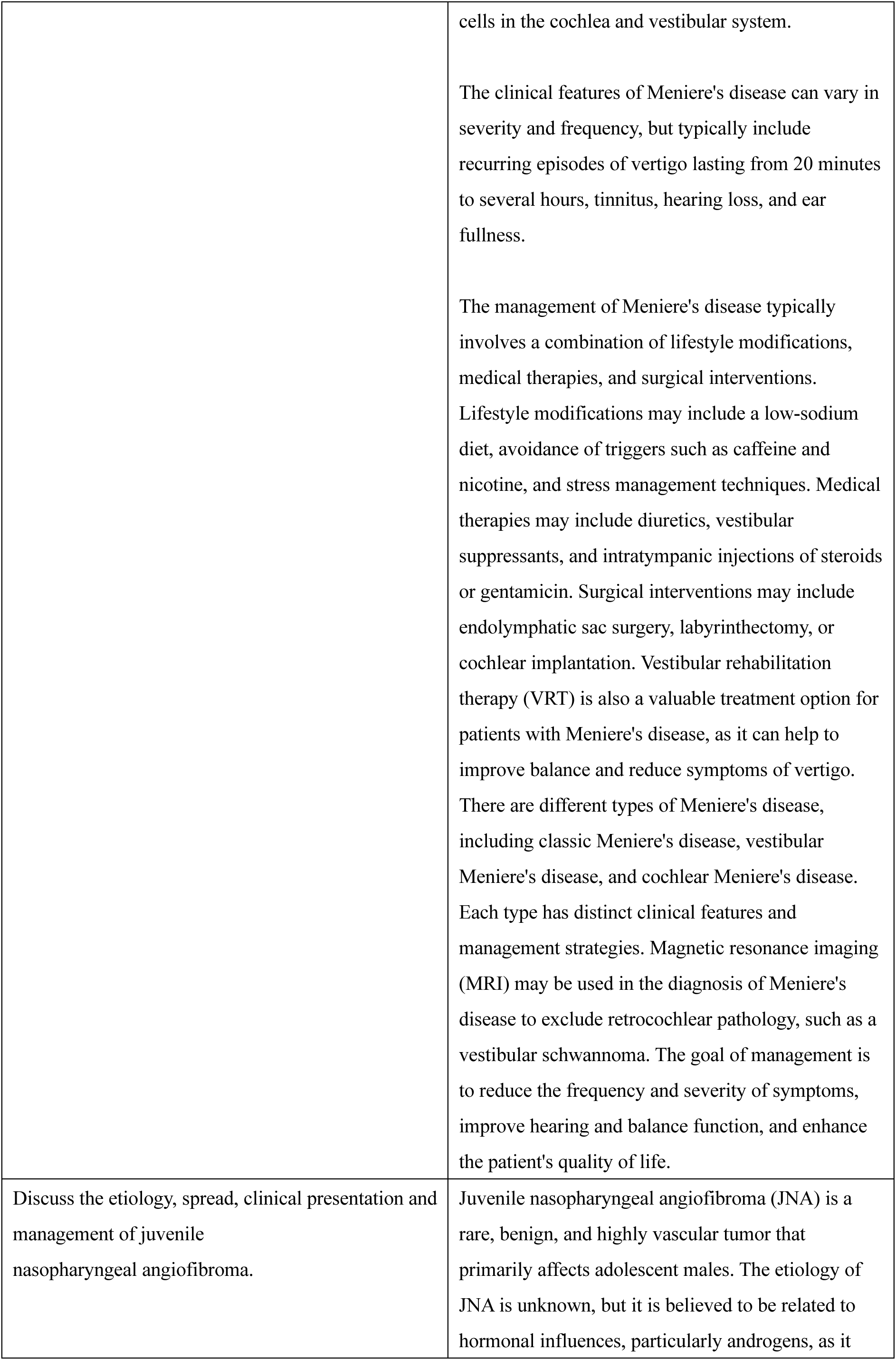
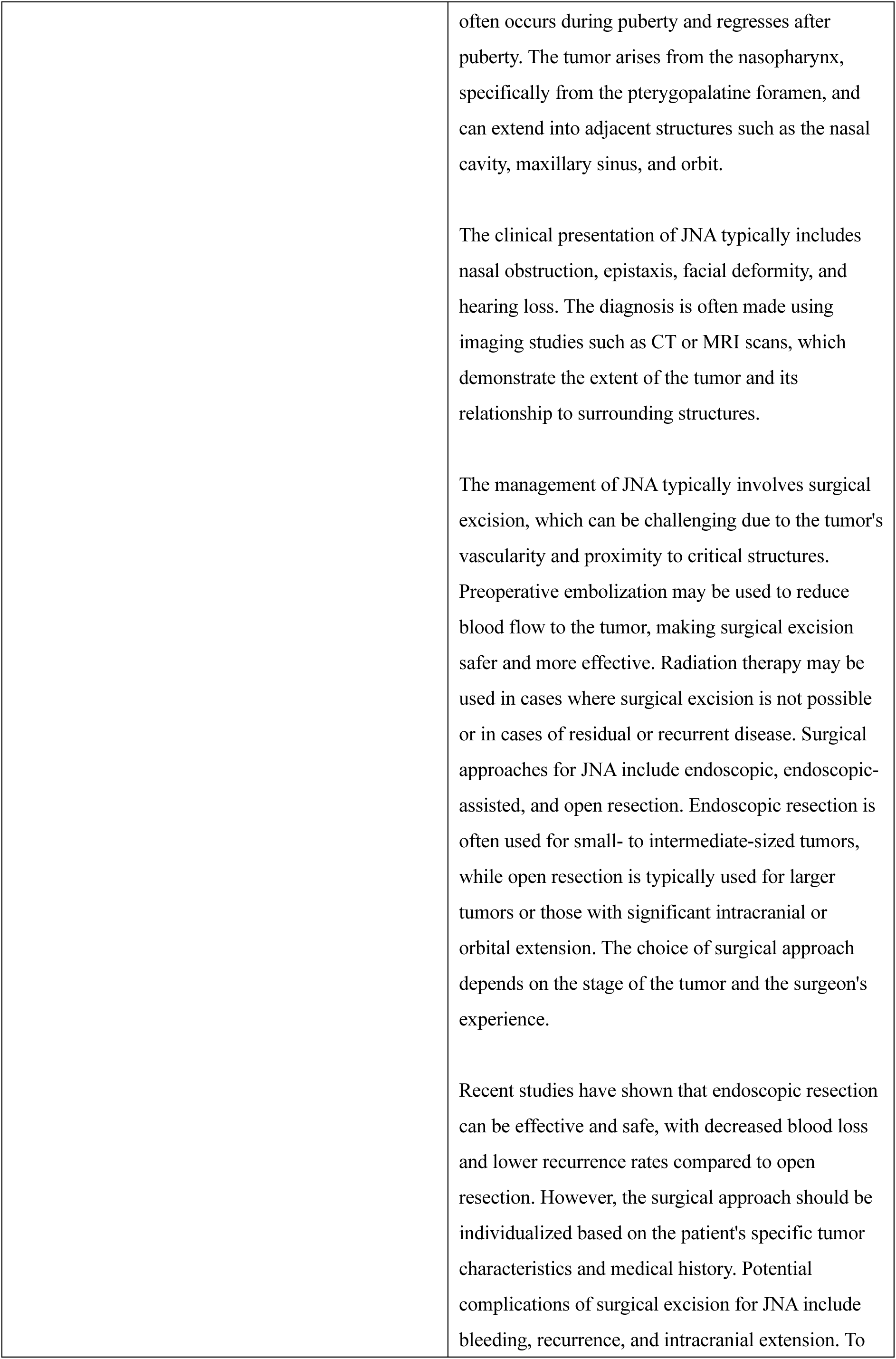
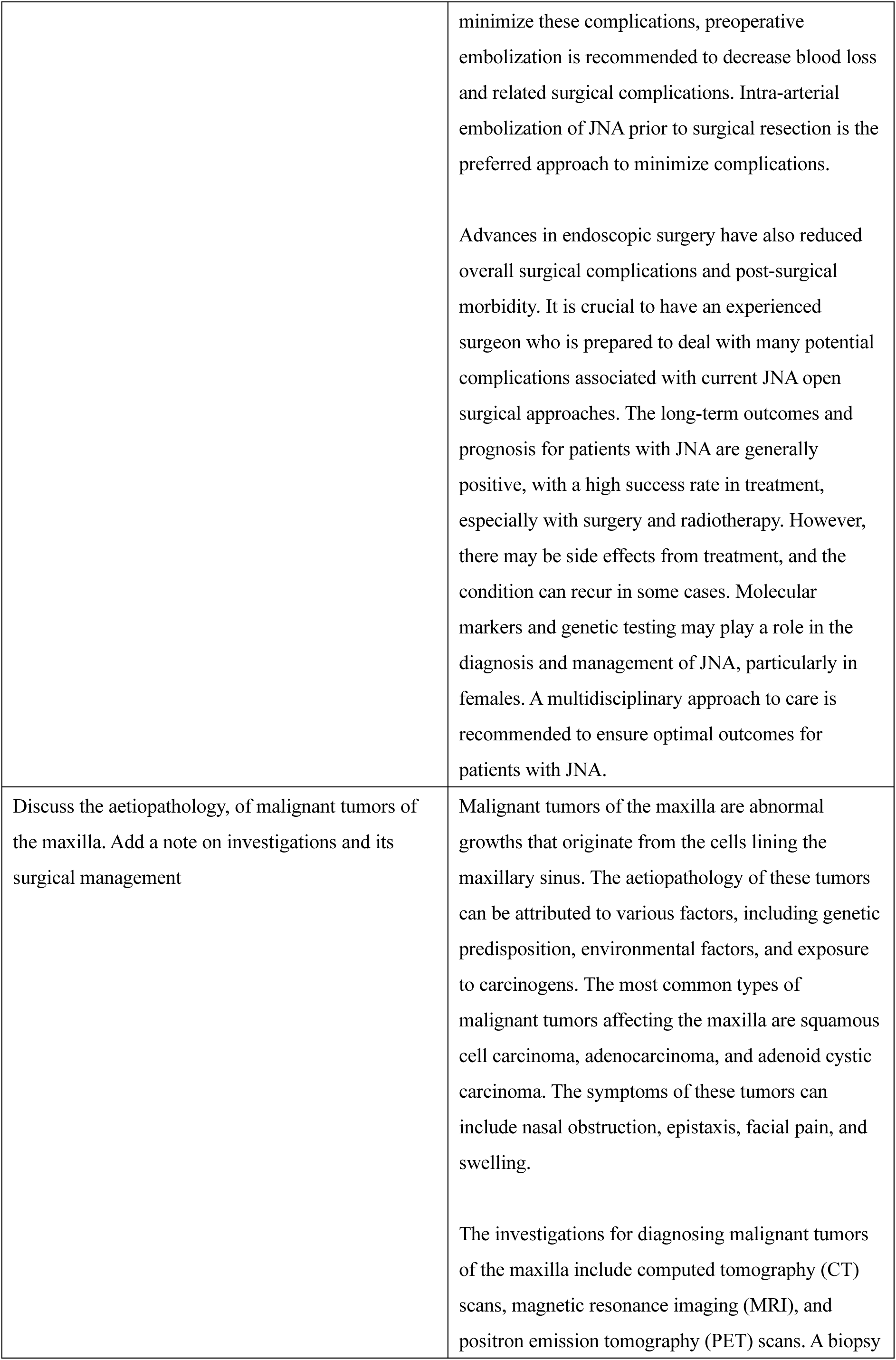
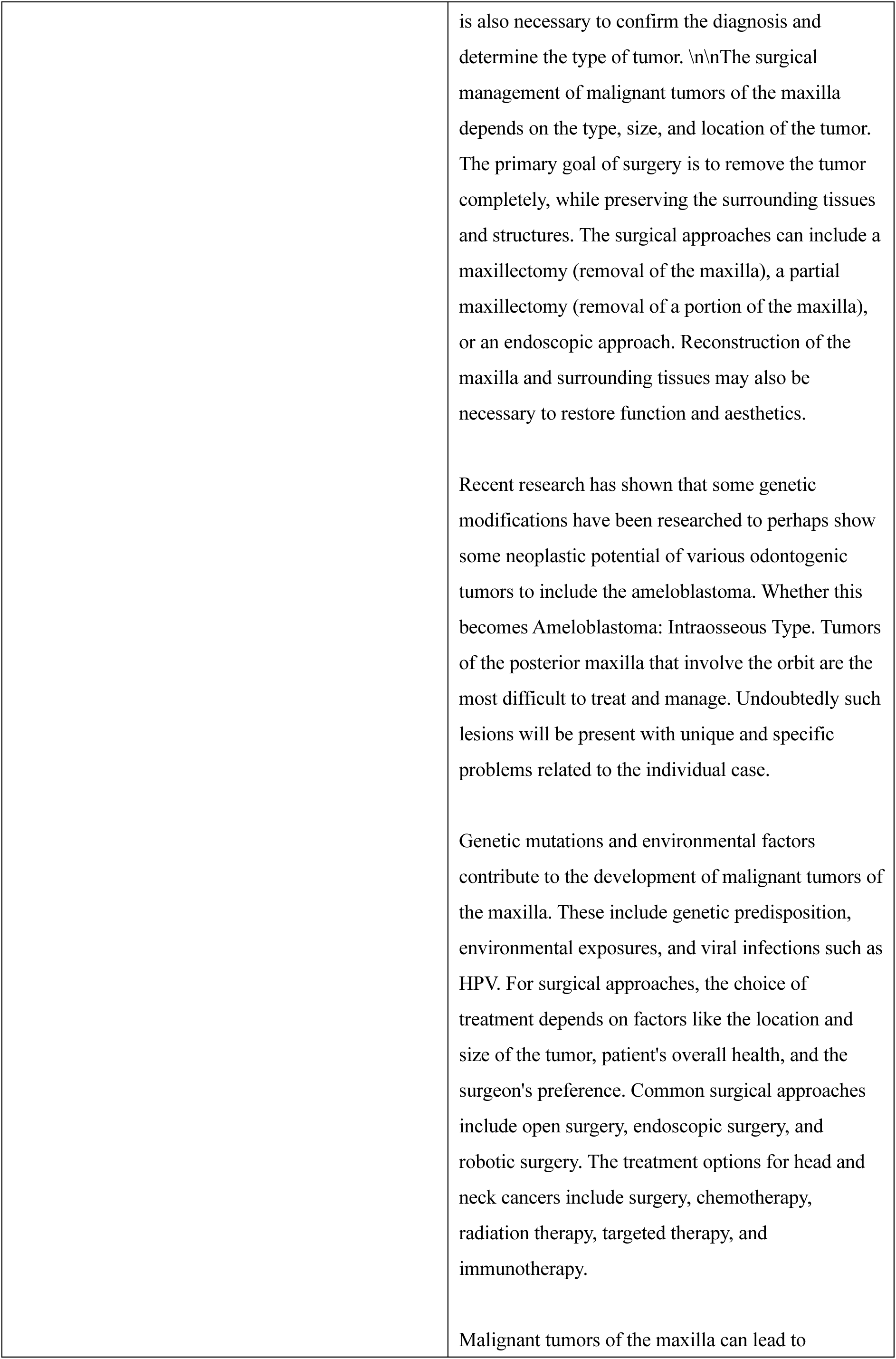
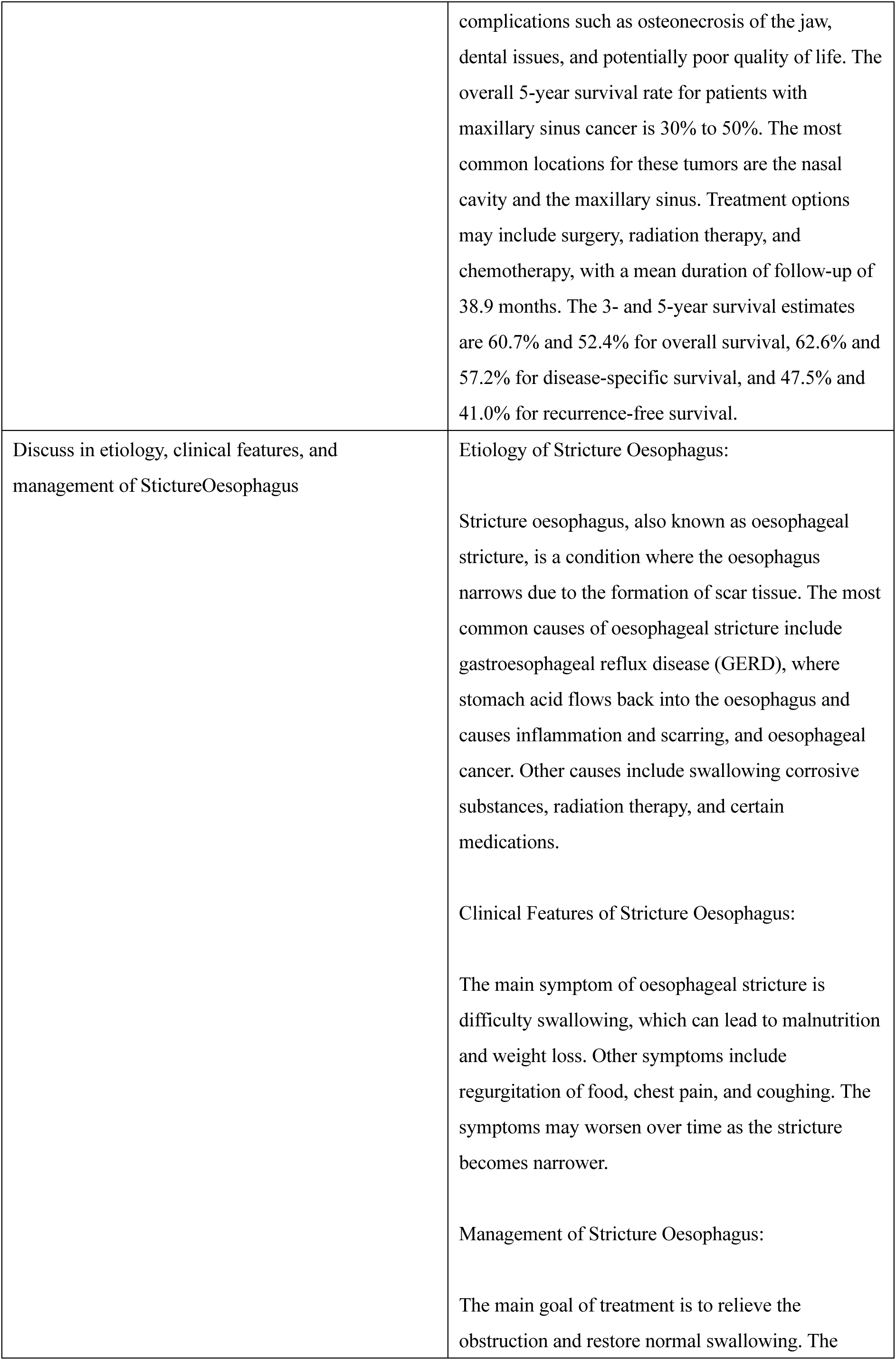
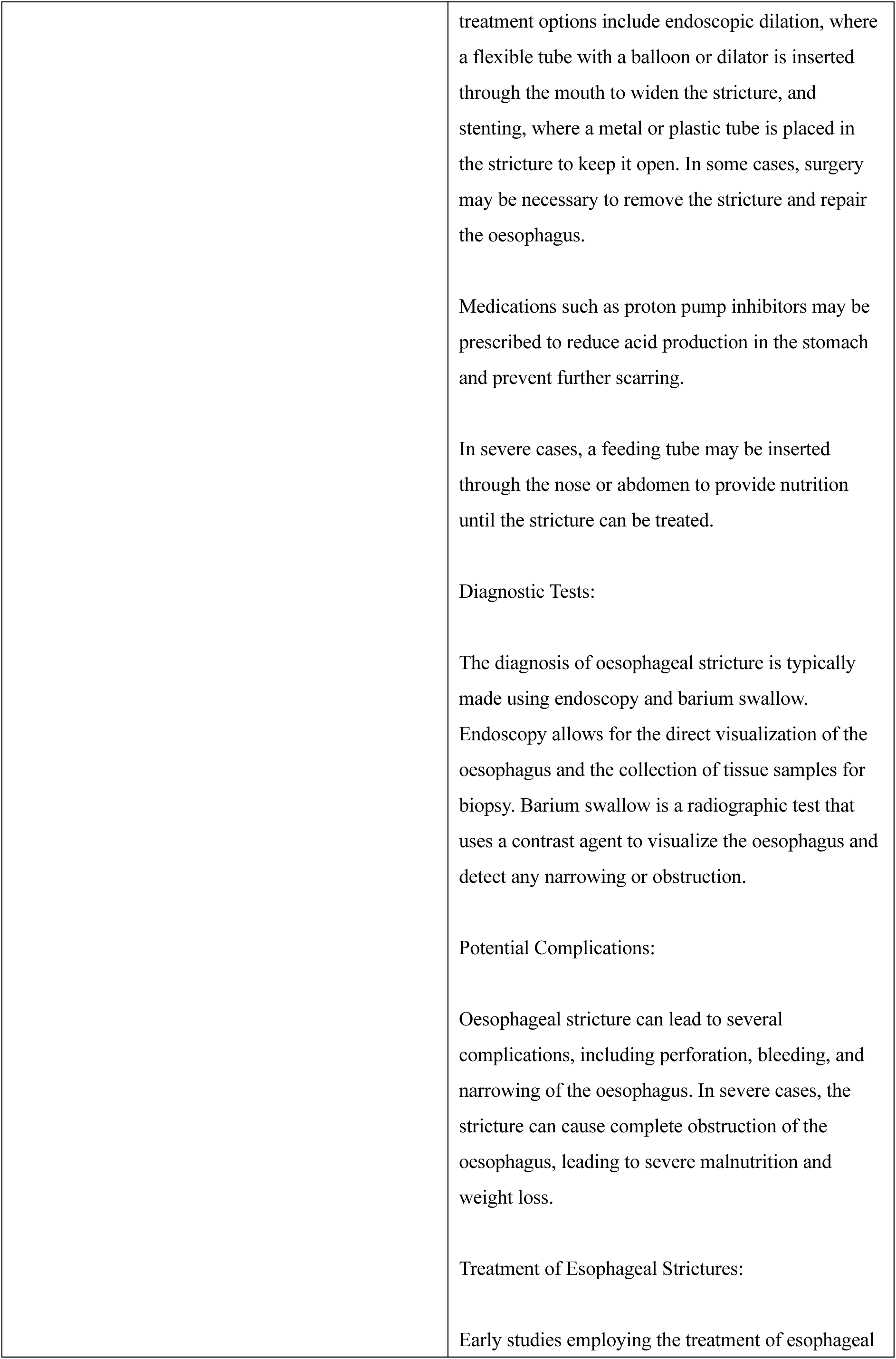
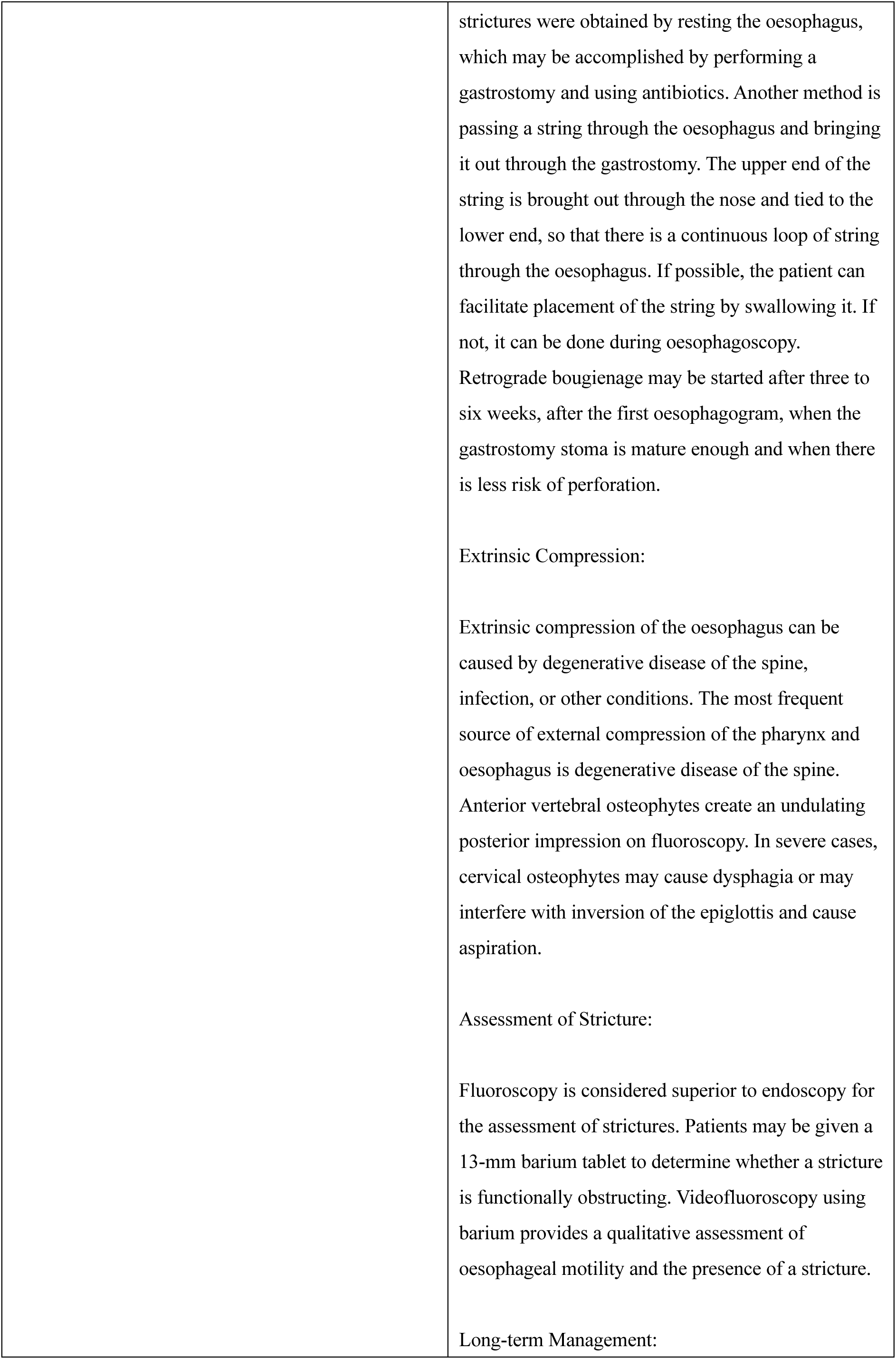
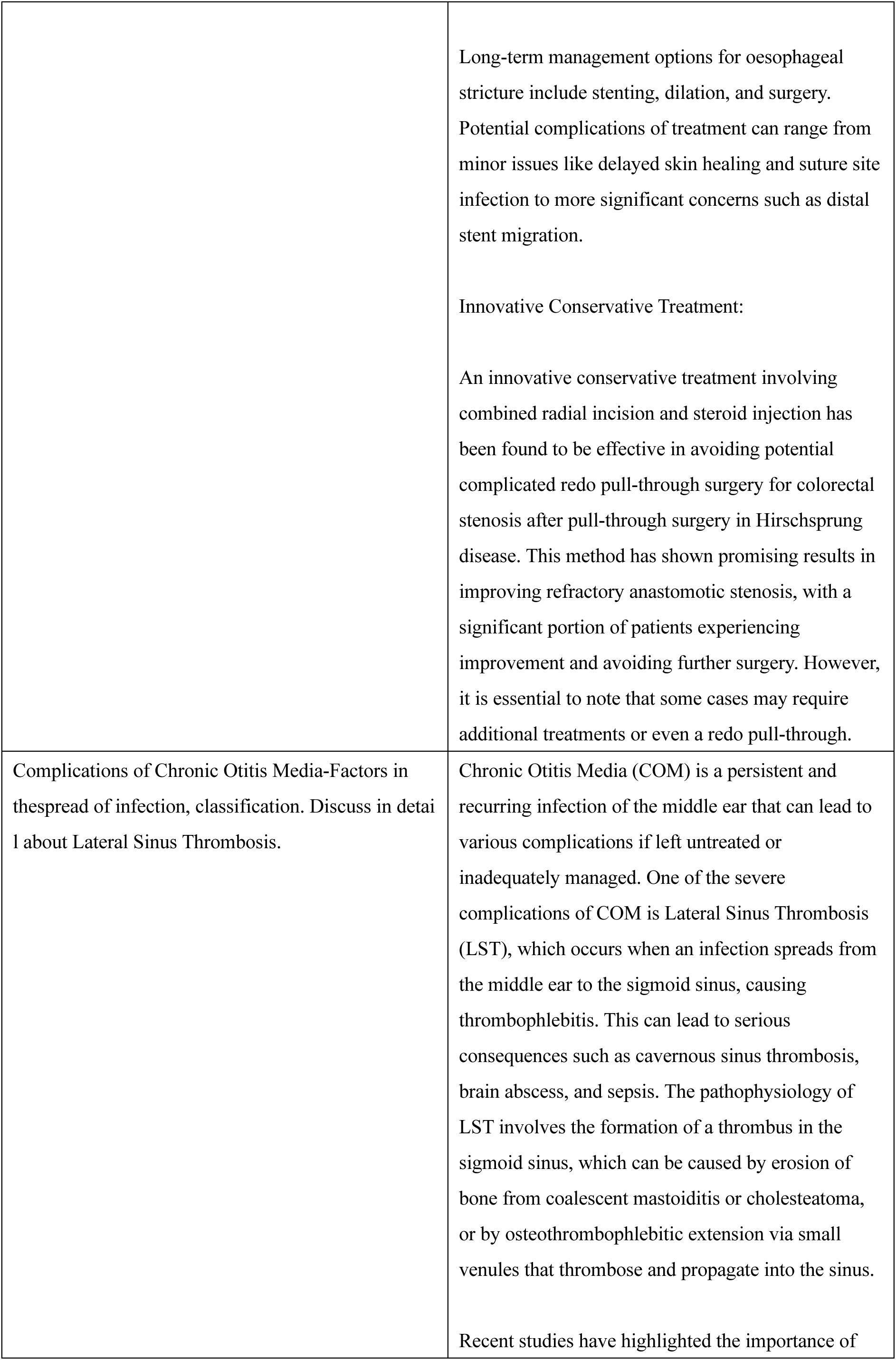
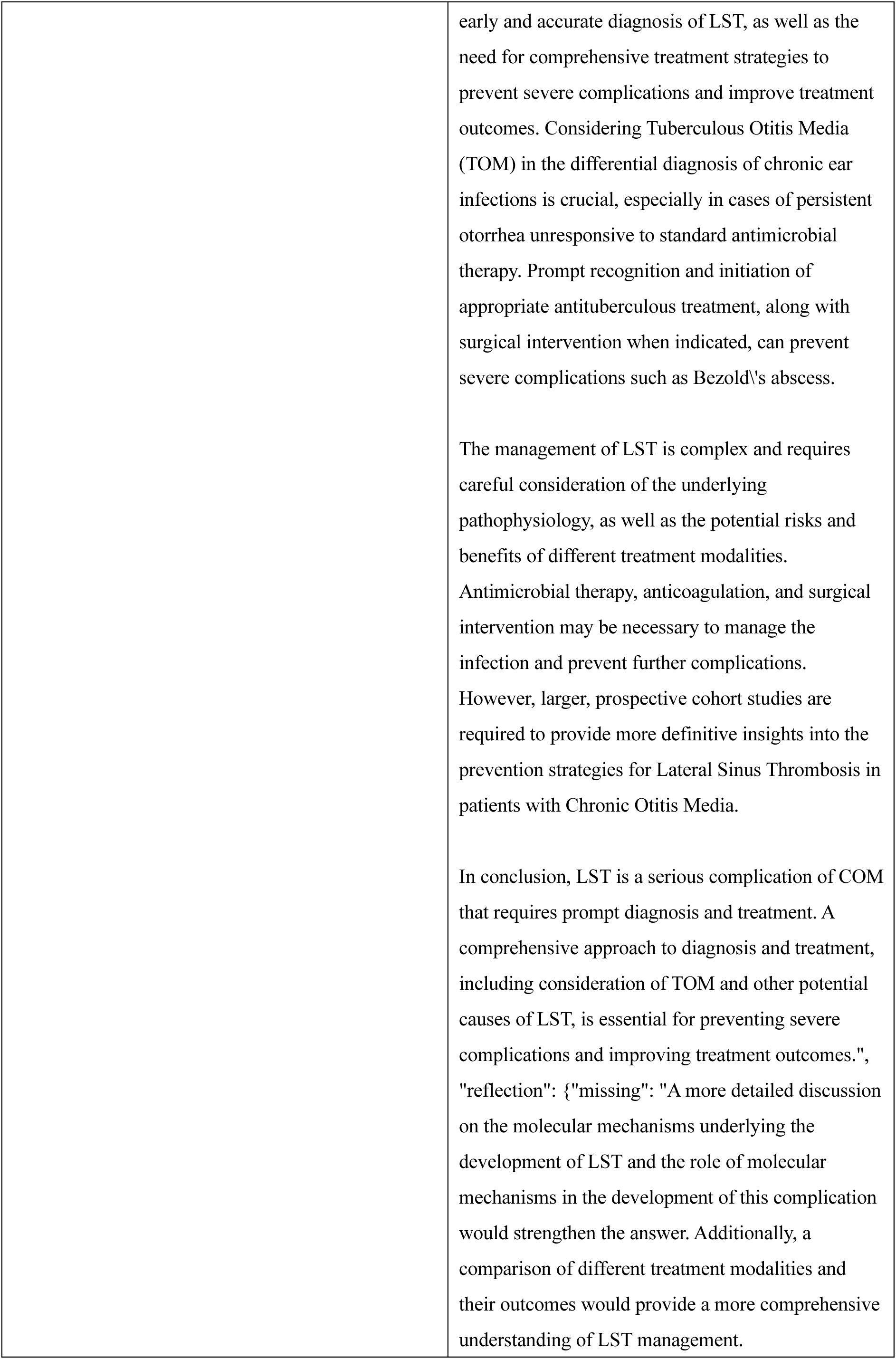
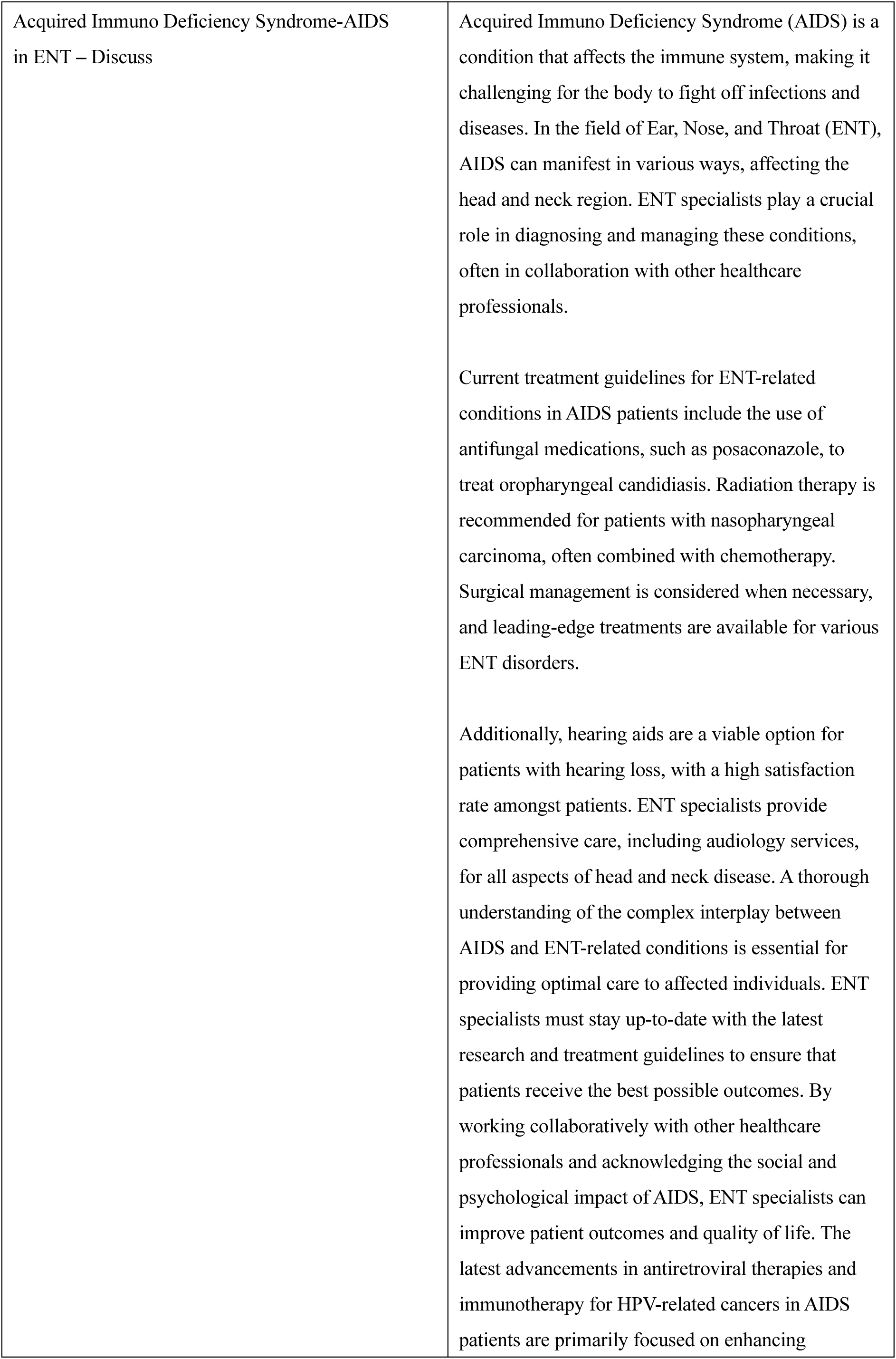
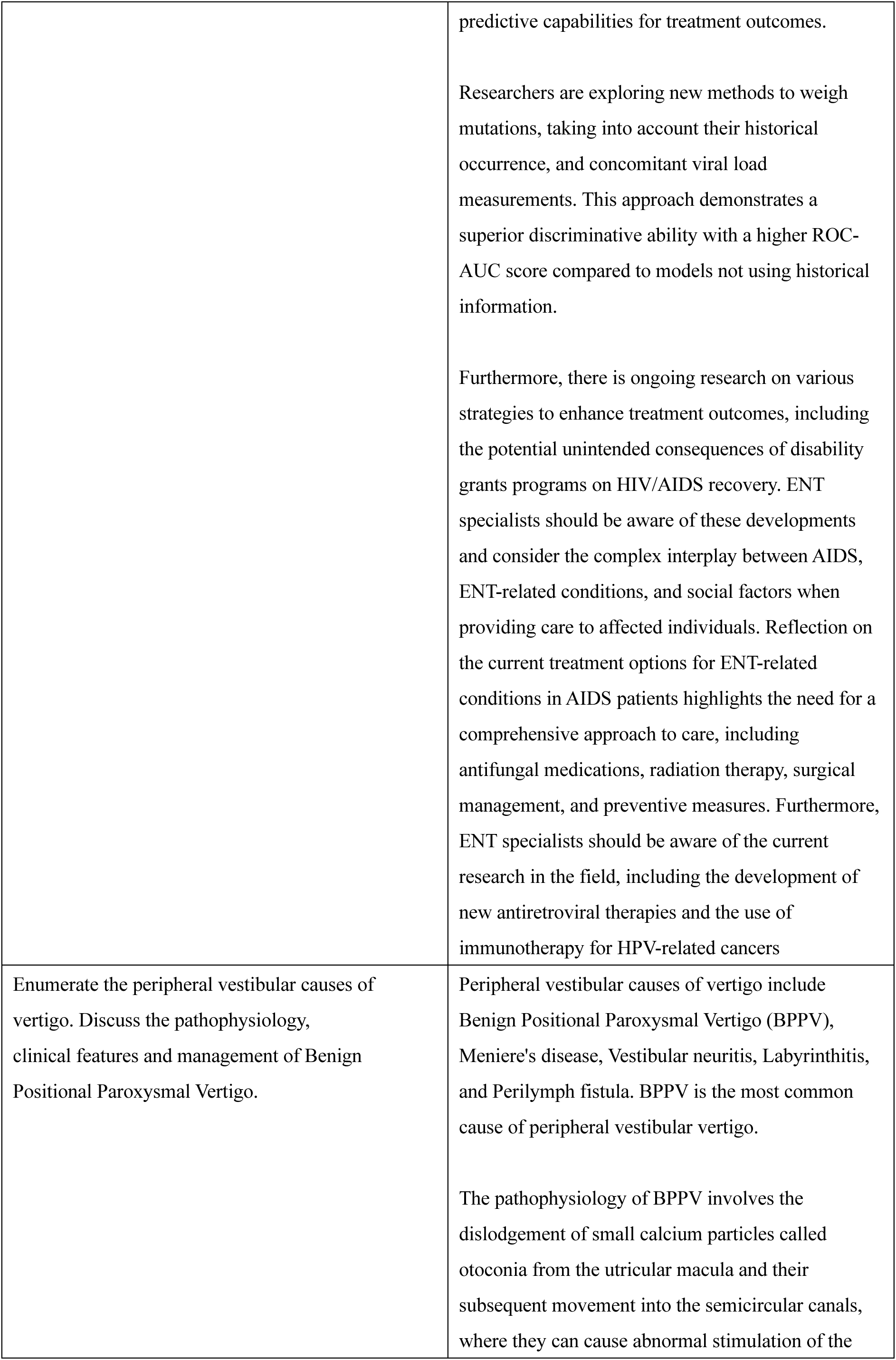
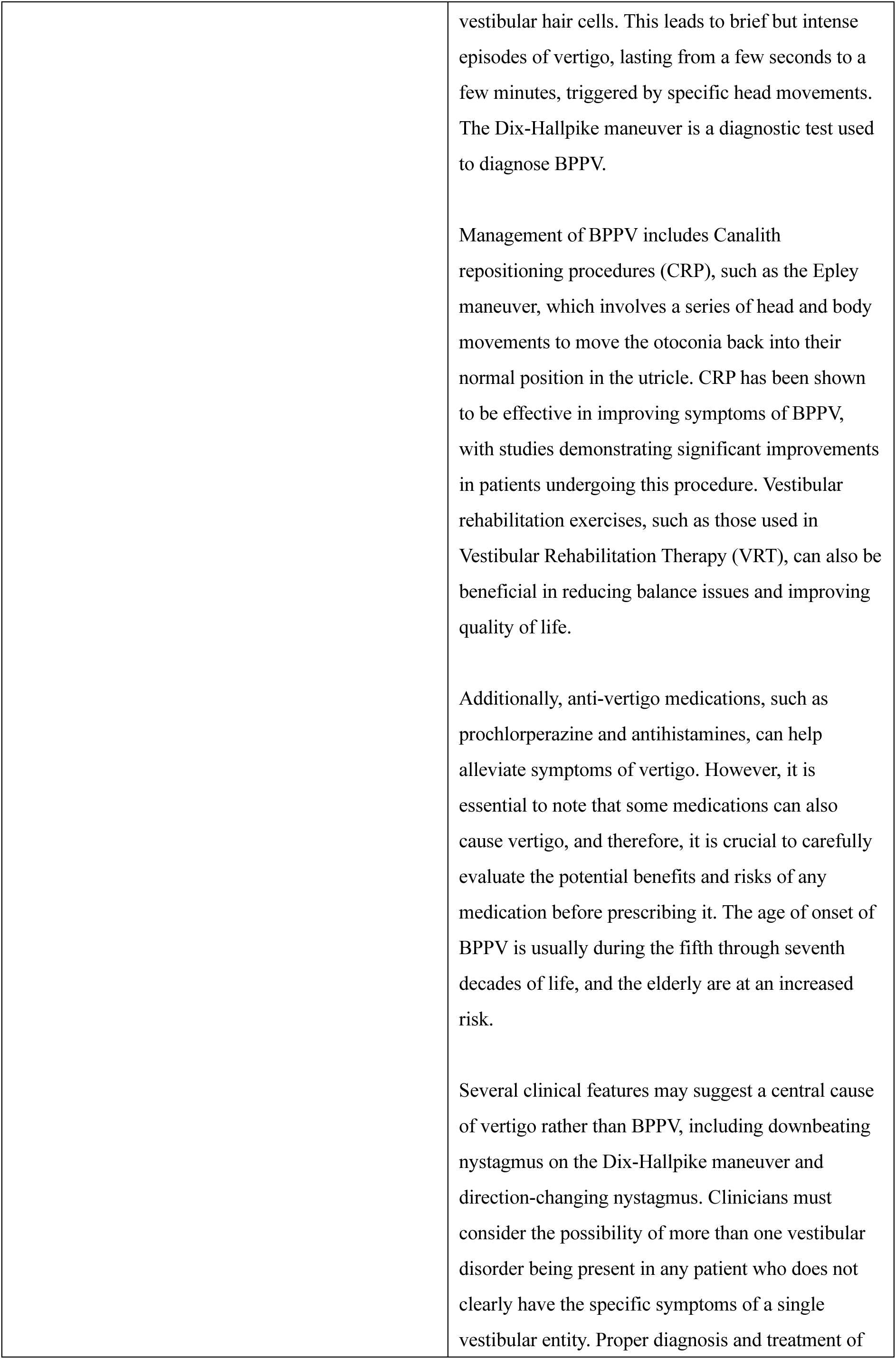
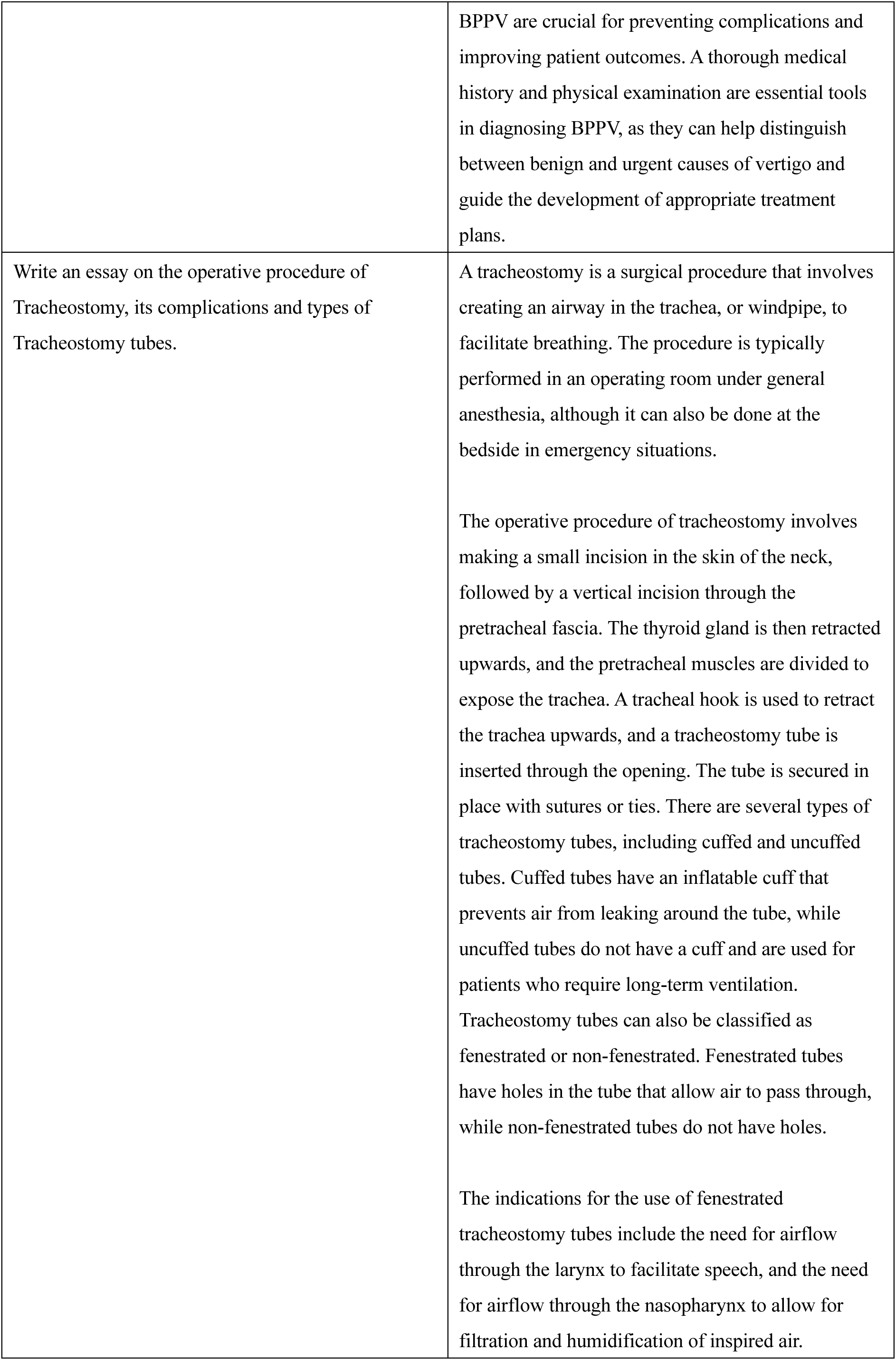
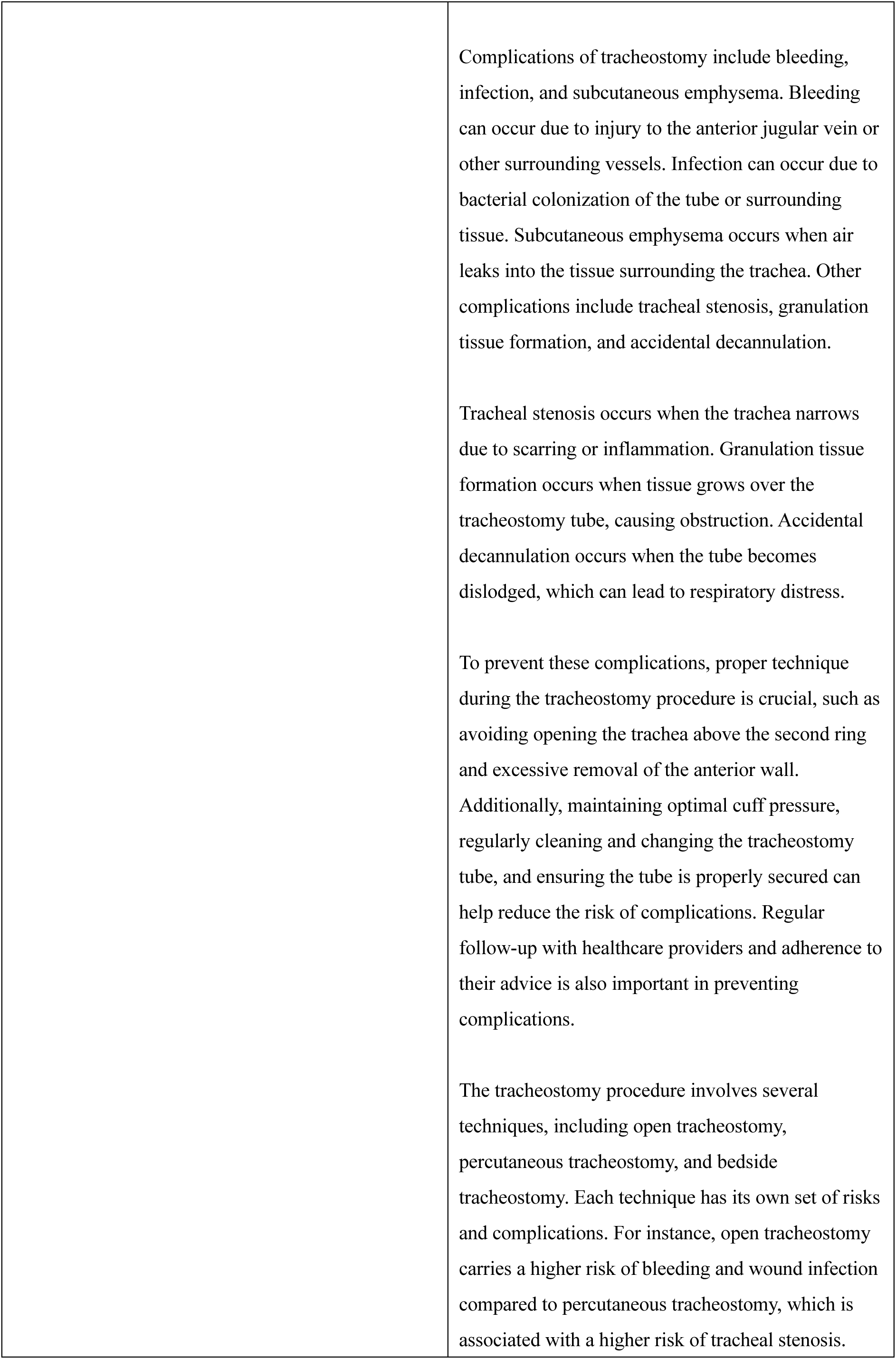
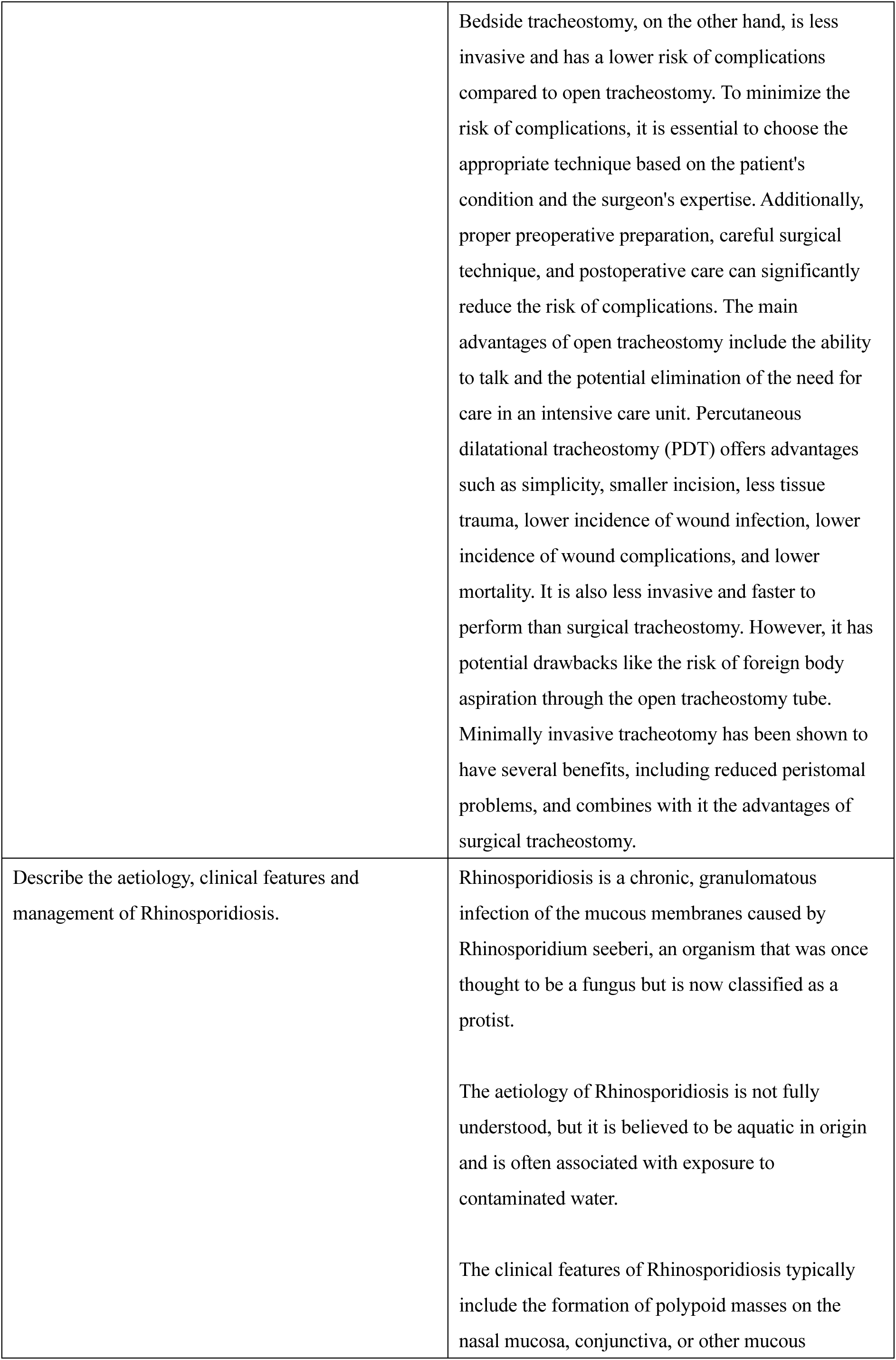
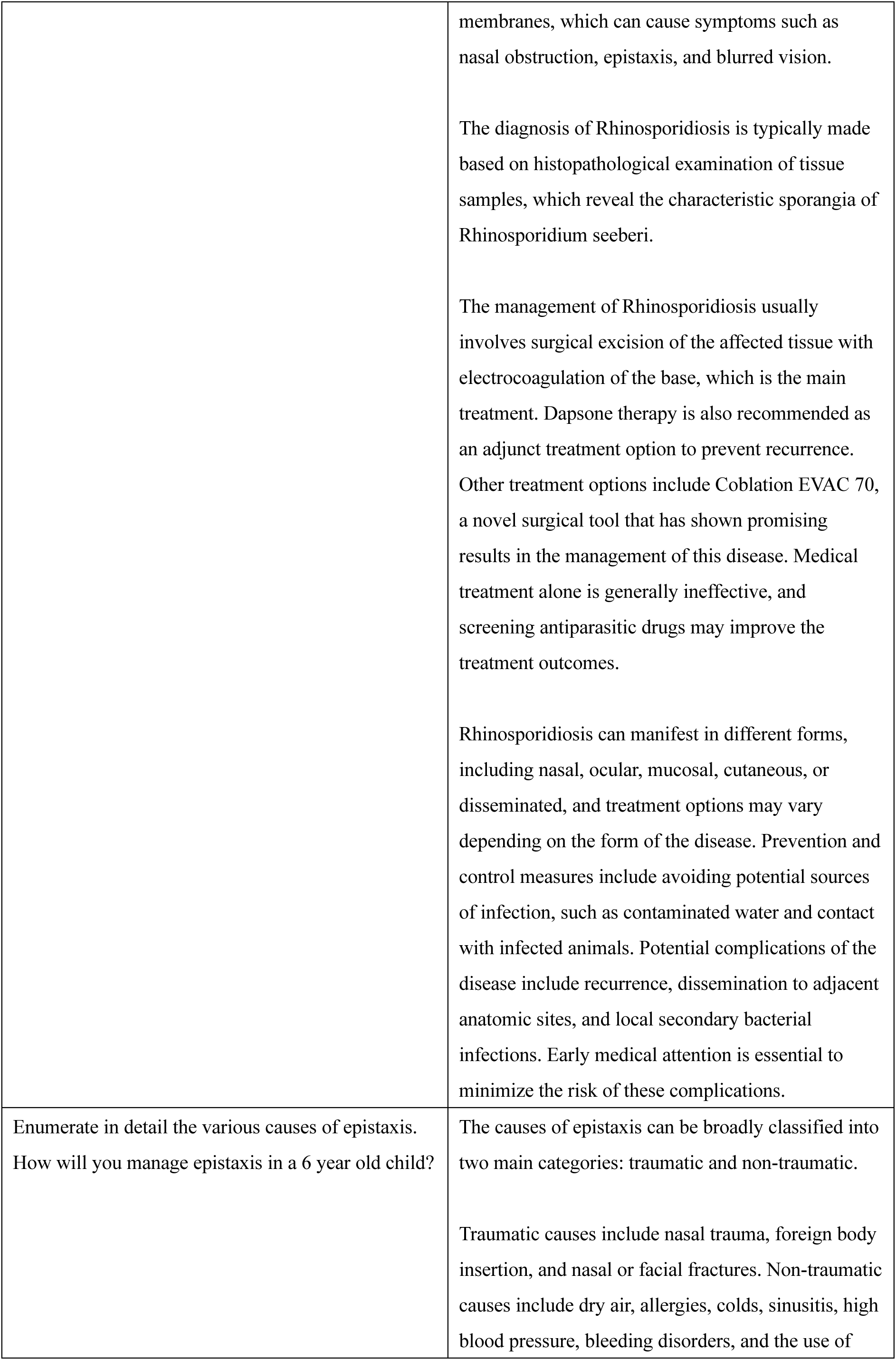
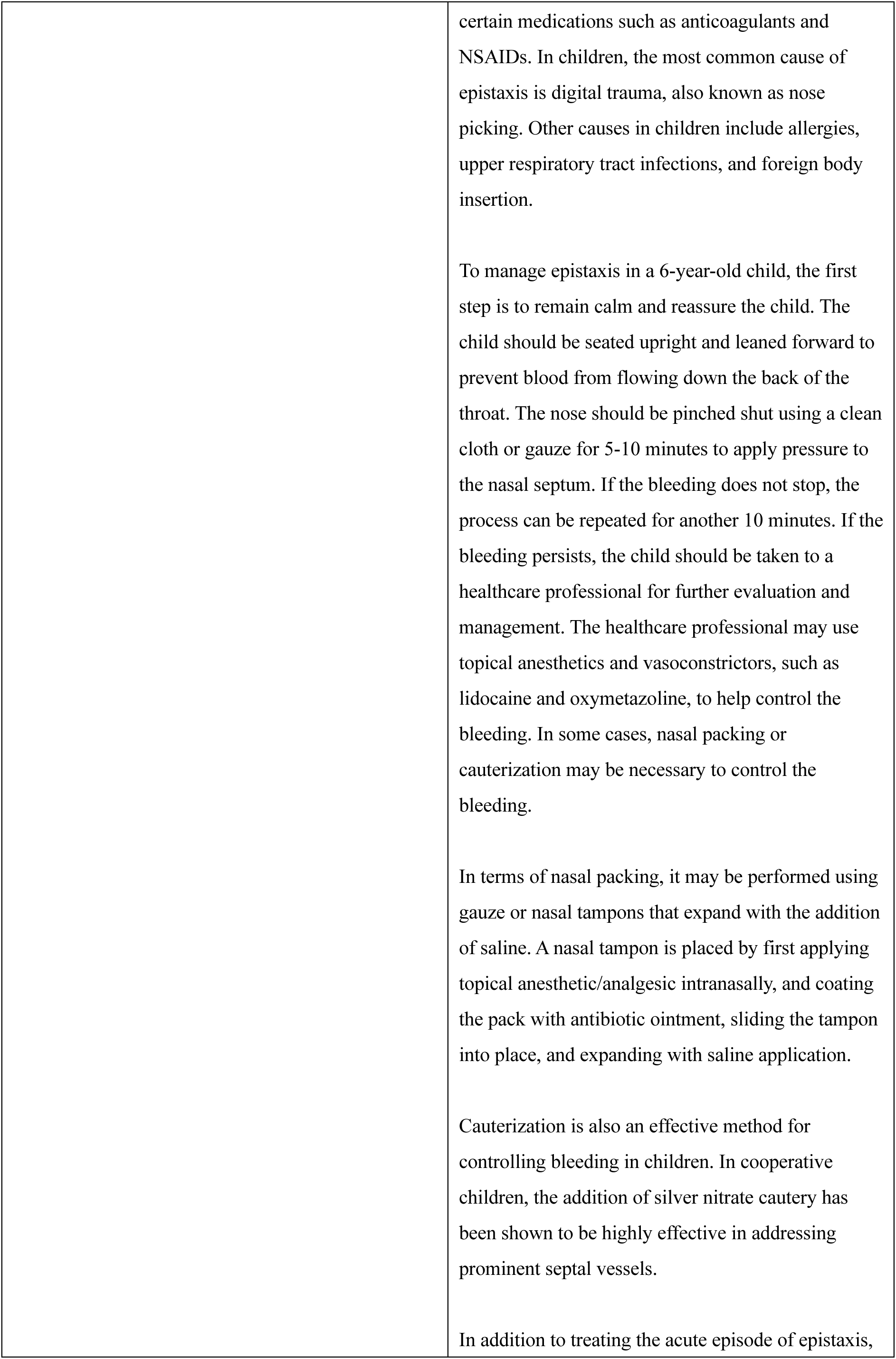
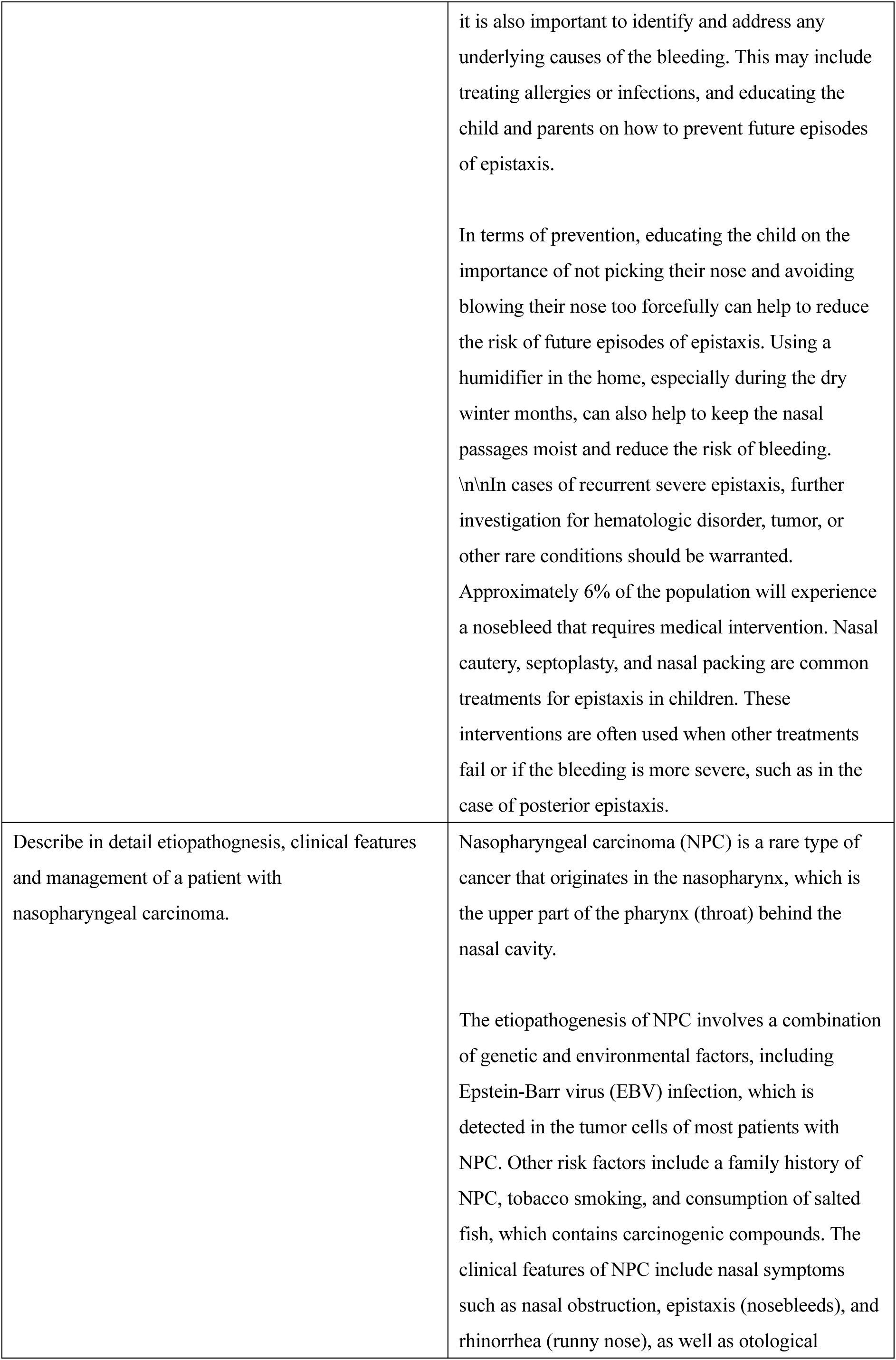
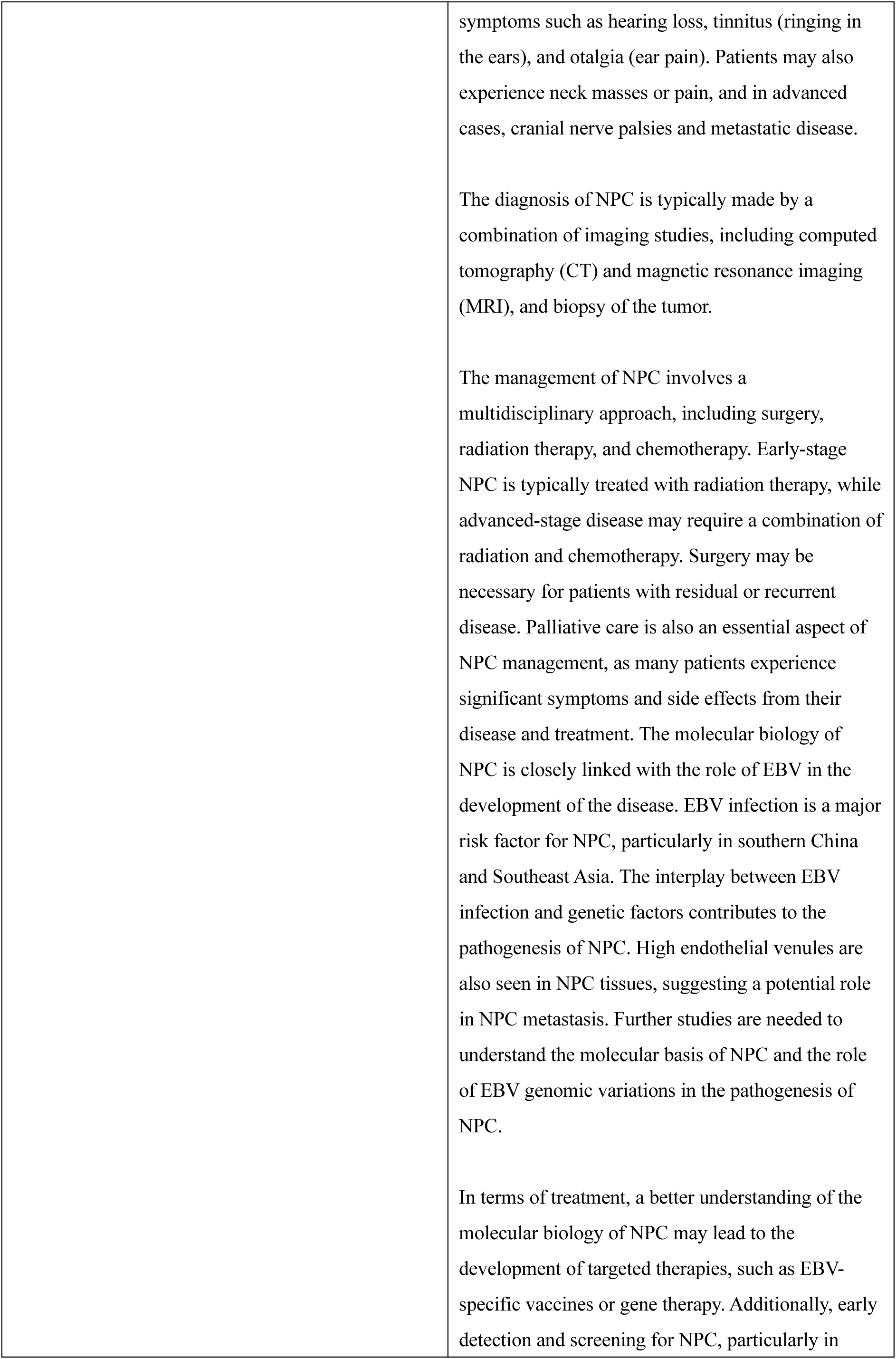
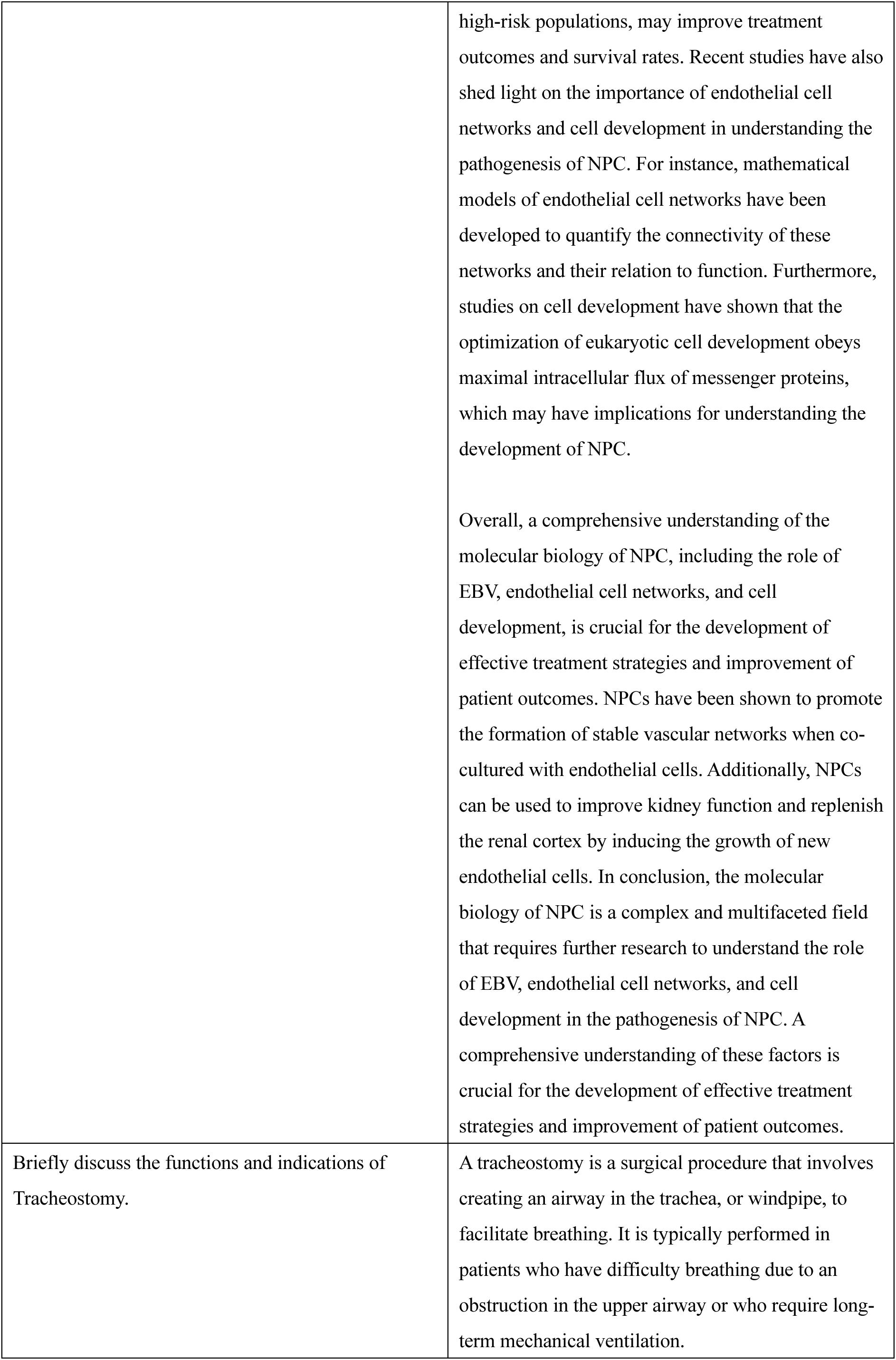
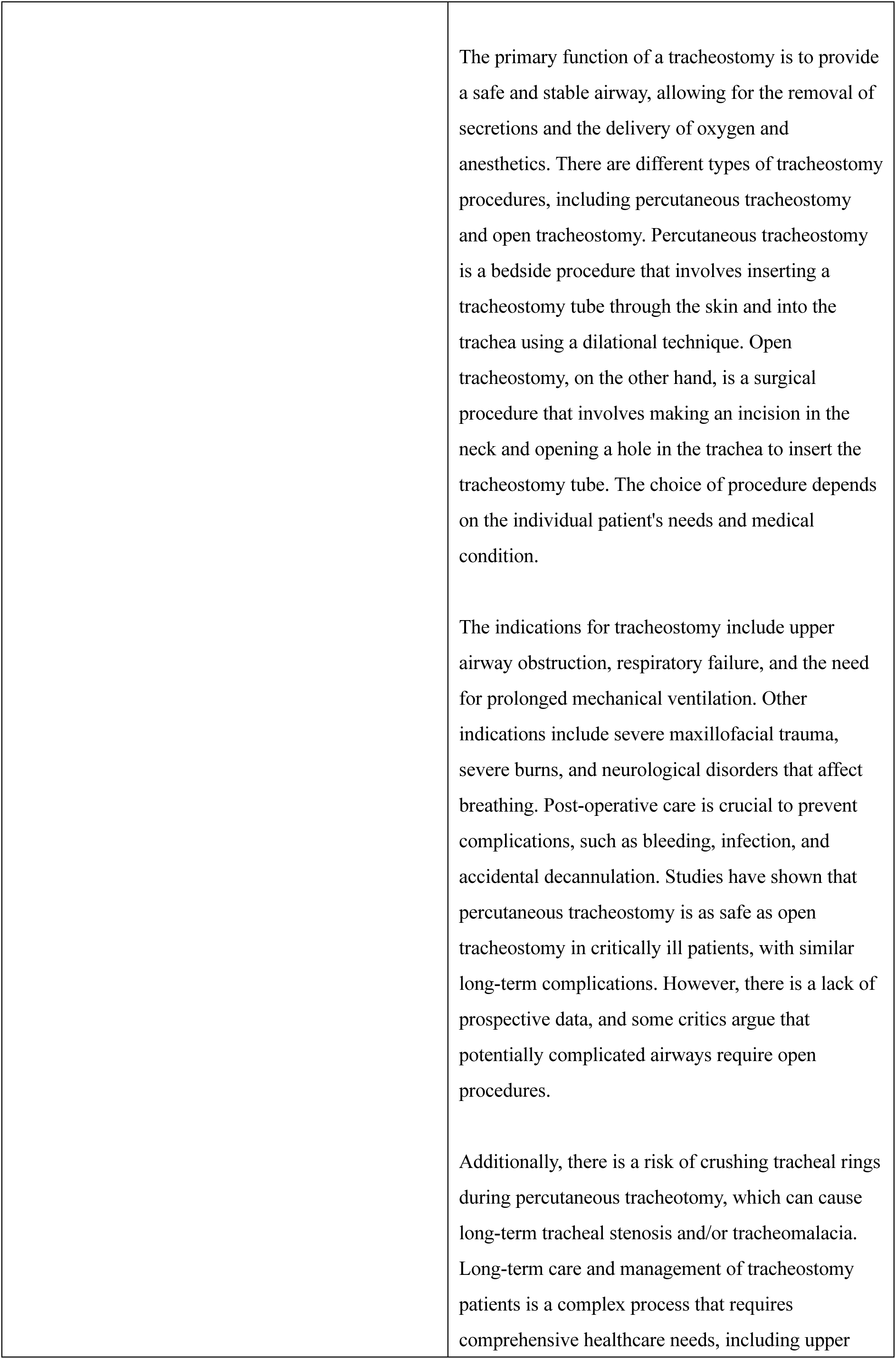
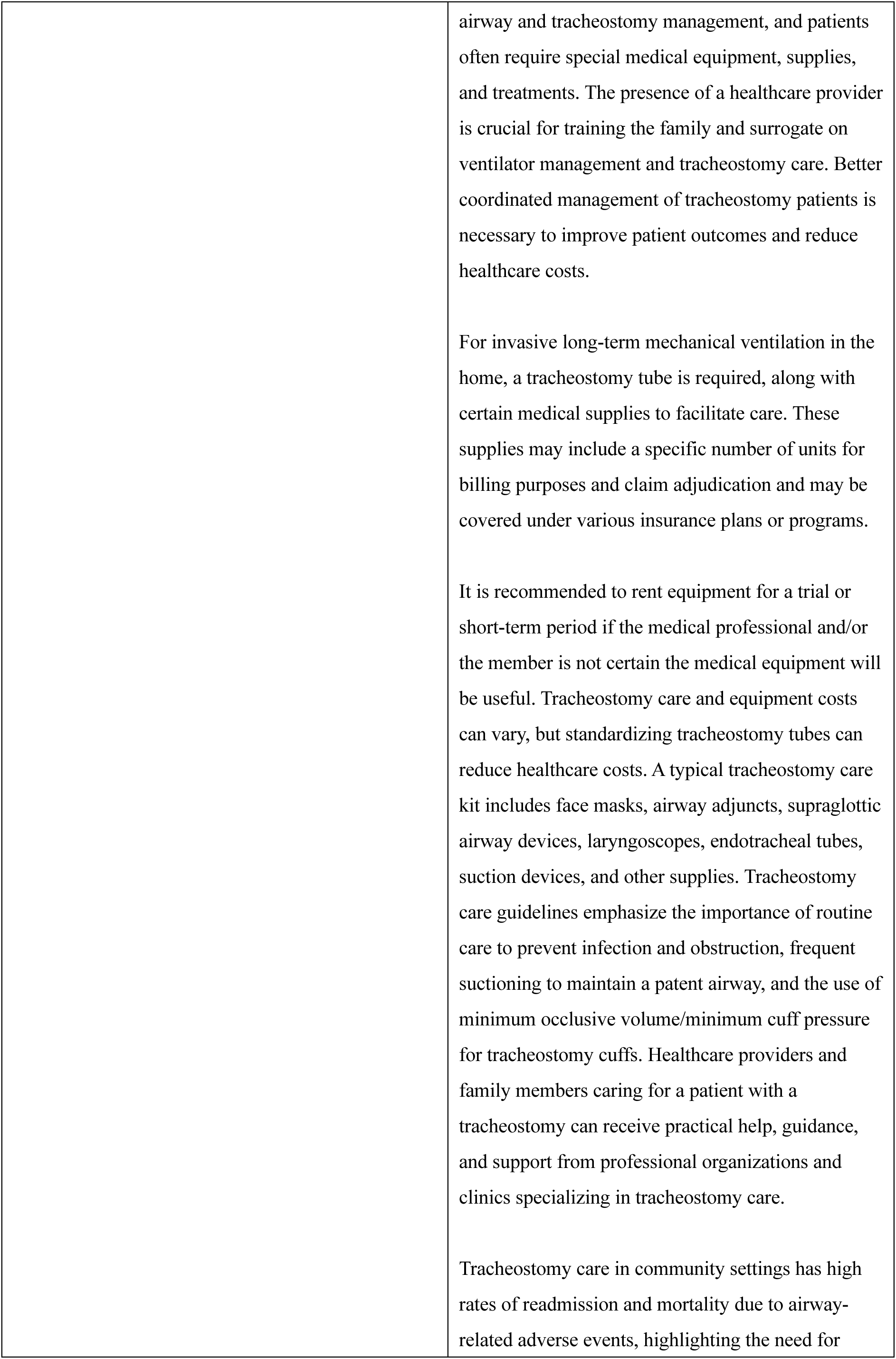
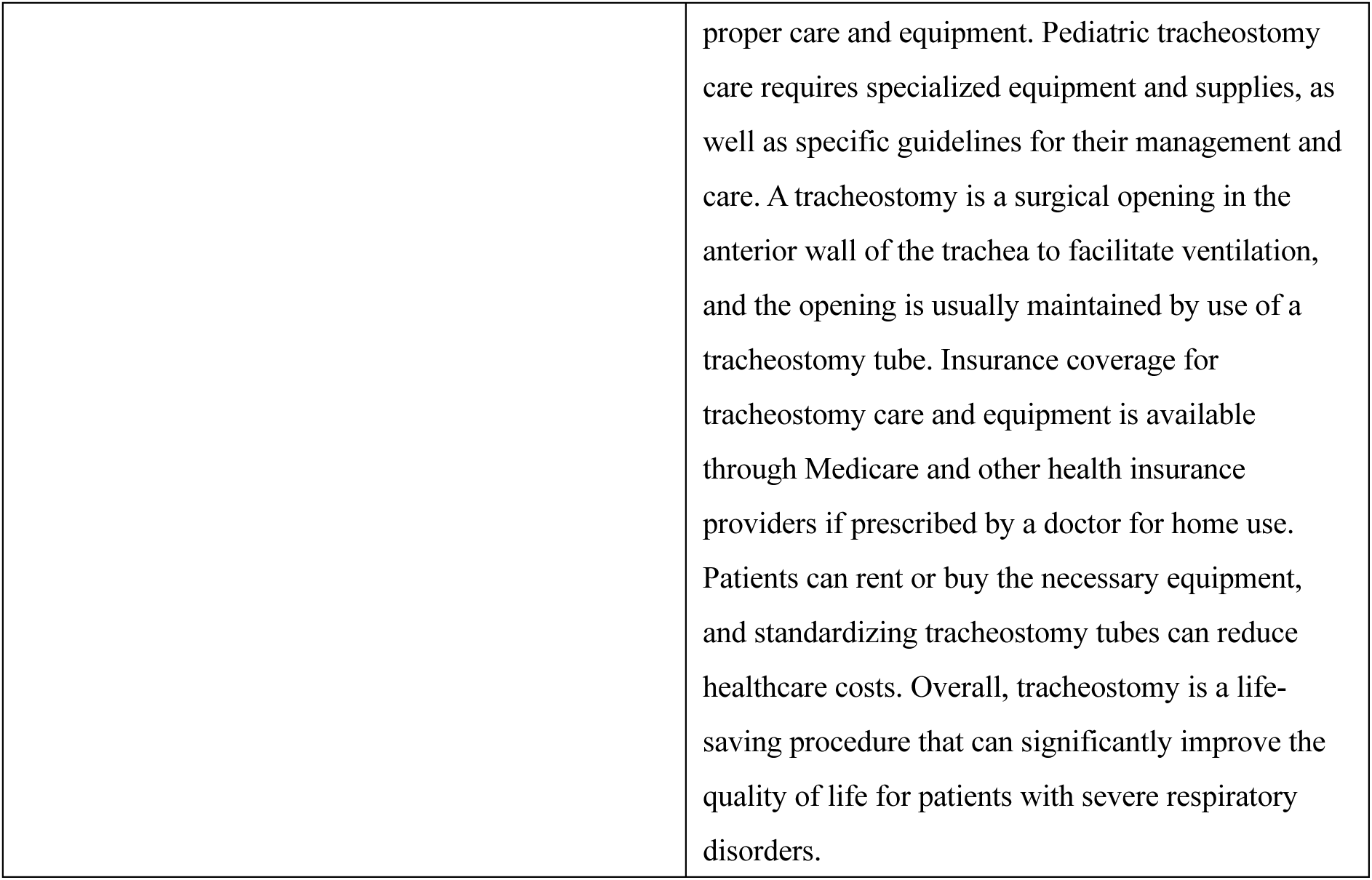
Essay questions and answers from ENTAgent.

